# Equitable deep learning for diabetic retinopathy detection using multi-dimensional retinal imaging with fair adaptive scaling: a retrospective study

**DOI:** 10.1101/2024.04.13.24305759

**Authors:** Min Shi, Muhammad Muneeb Afzal, Hao Huang, Congcong Wen, Yan Luo, Muhammad Osama Khan, Yu Tian, Leo Kim, Tobias Elze, Yi Fang, Mengyu Wang

## Abstract

**Background:** As deep learning becomes increasingly accessible for automated detection of diabetic retinopathy (DR), questions persist regarding its performance equity among diverse identity groups. We aimed to explore the fairness of current deep learning models and further create a more equitable model designed to minimize disparities in performance across groups.

**Methods:** This study used one proprietary and two publicly available datasets, containing two-dimensional (2D) wide-angle color fundus images, scanning laser ophthalmoscopy (SLO) fundus images, and three-dimensional (3D) Optical Coherence Tomography (OCT) B-Scans, to assess deep learning models for DR detection. We developed a Fair Adaptive Scaling (FAS) module that dynamically adjusts the significance of samples during model training for DR detection, aiming to lessen performance disparities across varied identity groups. FAS was incorporated into both 2D and 3D deep learning models to facilitate the binary classification of DR and non-DR cases. The area under the receiver operating characteristic curve (AUC) was adopted to measure the model performance. Additionally, we devised an equity-scaled AUC metric, which evaluates model fairness by balancing overall AUC against disparities among groups.

**Findings:** Using in-house color fundus images on the racial attribute, the overall AUC and ES-AUC of EfficientNet, after integrating with FAS, improved from 0.88 and 0.83 to 0.90 and 0.84 (p < 0.05), with AUCs for Asians and Whites improving by 0.04 and 0.03, respectively (p < 0.01). Regarding gender, both the overall AUC and ES-AUC of EfficientNet improved by 0.01 (p < 0.05) after integrating with FAS. While using in-house SLO fundus images based on race, the overall AUC and ES-AUC of EfficientNet improved from 0.80 to 0.83 (p < 0.01), with AUCs for Asians, Blacks, and Whites improving by 0.02, 0.01 and 0.04, respectively (p < 0.05). On gender, FAS improved the overall AUC and ES-AUC of EfficientNet by 0.02, with both genders showing an improvement of 0.02 (p < 0.01). Using the 3D deep learning model DenseNet121 on in-house OCT-B-Scans based on race, FAS improved the overall AUC and ES-AUC from 0.875 and 0.81 to 0.884 and 0.82 respectively, where the AUCs for Asians and Blacks improved by 0.03 and 0.02 (p < 0.01). On gender, FAS improved the overall AUC and ES-AUC of DenseNet121 by 0.04 and 0.03, while the AUCs for Females and Males improved by 0.05 and 0.04 (p < 0.01), respectively.

**Interpretation:** Existing deep learning models indeed exhibit variable performance across diverse identity groups in DR detection. The FAS proves beneficial in enhancing model equity and boosting DR detection accuracy, particularly for underrepresented groups.

## Introduction

Diabetic retinopathy (DR) is a common complication of diabetes that affects blood vessels in the retina,^1,2^ and is the leading cause of blindness in adults aged 20 to 74 years in the United States.^3–5^ As DR can develop at any point in the lifespan of a diabetic patient, regular eye examinations conducted by an ophthalmologist are crucial for the timely detection of DR, which enables prompt treatment vital in preserving vision. However, regular access to ophthalmic care is often hindered by a scarcity of eye care resources and the high costs associated with specialty care. It has been reported that racial and ethnic minorities such as Blacks and Hispanics are disproportionally affected, with DR prevalence 50% higher than non-Hispanic Whites.^6–9^ Additionally, Black and Hispanic patients with DR (Odds ratios: 1.78 and 1.68, respectively) are more likely to present with worse vision loss compared with non-Hispanic Whites.^10,11^ Although the DR disease burden is significantly greater in minorities, various studies have reported that the rate of eye examination for DR screening is significantly lower in these minority groups (49%) compared to non-Hispanic Whites (59%).^12^

In recent years, automated DR detection using deep learning^13–16^ through retinal imaging has emerged as an affordable solution, providing frequent and regular eye examinations for timely DR detection. This innovation aims to alleviate societal disease burdens and reduce health disparities among different demographic groups. Numerous studies have been conducted to develop deep learning models for automated DR detection,^13,14,16,17^ yet it remains unclear if these deep learning models perform equitably across different identity groups. Ensuring equitable performance is vital in any disease screening model to uphold the principles of social justice and fairness. The performance inequality observed in deep learning models may primarily stem from two factors: data inequality and data characteristic variability among different identity groups. For example, fewer Black and Asian DR patients are present in ophthalmic care, representing data inequality.^5^ Moreover, prior studies have shown that retinal anatomy varies with sex and racial information,^18,19^ which exemplifies data characteristic variability. Mitigating data inequality and addressing data characteristic variability is imperative to reduce performance disparities and achieve more equitable outcomes in deep learning. As of now, studies elucidating and harnessing the underlying performance disparities among demographic identity groups for DR screening with deep learning are limited.

In this study, we conducted a thorough assessment of state-of-the-art deep learning models for detecting DR using two-dimensional (2D) fundus images and three-dimensional (3D) optical coherence tomography (OCT) B-Scans. We examined disparities in model performance across various identity groups, including race, gender, ethnicity, marital status, and preferred language. Furthermore, we developed an equitable deep learning model that enhances equity in automated DR detection (**Figure 1**). The core idea of our approach is to introduce a Fair Adaptive Scaling (FAS) module that dynamically adjusts the significance of individual samples during training to achieve equitable DR detection performance among different identity groups. We tested the effectiveness of these models using a comprehensive proprietary dataset and two public datasets designed for DR detection, encompassing wide-angle color fundus, Scanning Laser Ophthalmoscopy (SLO) fundus, and OCT B-Scans. We used the area under the receiver operating characteristic curve (AUC) to compare the DR detection performance of various models. In addition, to balance the overall AUC with performance disparities among diverse identity groups, we introduced a novel metric termed equity-scaled AUC (ES-AUC), designed to measure and compare the fairness of different deep learning models.

**Figure 1.**
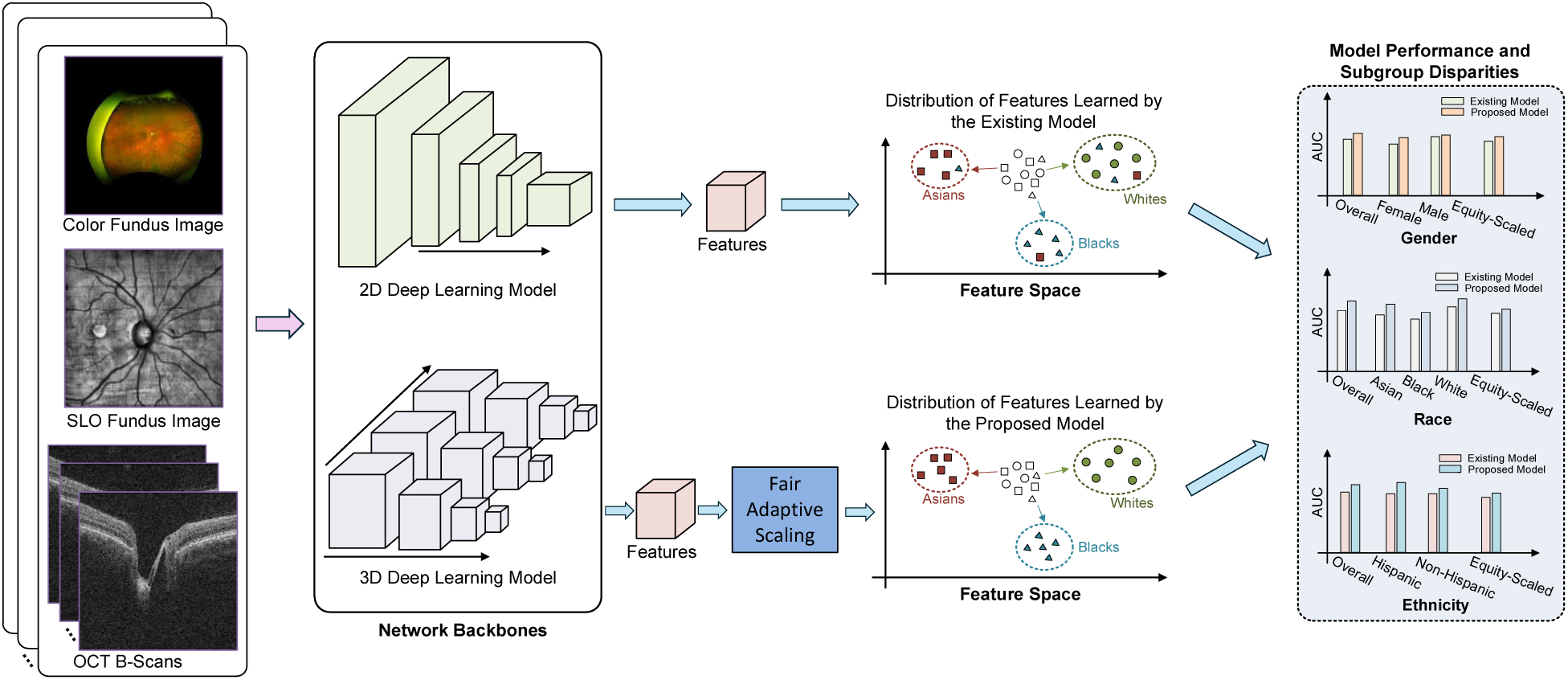
An illustration of the fairness issue in DR detection. Exiting deep learning models demonstrate significant group performance disparities measured by equality-scaled AUC. We proposed a fair adaptive scaling module to improve model performance disparities across different identity groups.

## Methods

### Ethics statement

The fundus and OCT data used for developing the equitable deep learning model were from Massachusetts Eye and Ear (MEE) between 2021 and 2023. The institutional review boards (IRB) of MEE approved the creation of the database in this retrospective study. This study complied with the guidelines outlined in the Declaration of Helsinki. In light of the study’s retrospective design, the requirement for informed consent was waived.

### Datasets

We adopted three different datasets to validate the existing and proposed equitable deep learning methods for DR detection. They are described as follows.

#### MEE Inhouse Data

This dataset spans three different data modalities, including wide-angle color fundus images, SLO fundus images, and 3D OCT B-Scans, where SLO fundus images and OCT B-scans are exactly paired from the same eye at the same visit. The wide-angle color fundus images were collected from 22,622 patients with an average age of 57.4 ± 19.4 years. The demographic distributions are as follows (**Supplemental Figure 1**): Gender: Female: 51.6%, Male: 48.4%; Race: Asian 6.5%, Black: 9.3%, White: 84.2%. Ethnicity: Non-Hispanic: 96.7%, 3.3% Hispanic; Preferred Language: English: 93.1%, Spanish: 1.3%, Others: 5.6%; Marital Status: Married or Partnered: 53.6%, Single: 29.7%, Divorced: 6.2%, Legally Separated: 0.7%, Widowed: 6.7%, Unknown: 3.1%. The SLO fundus images and OCT B-scans were collected from 49,164 patients with an average age of 63.9 ± 17.4 years. The demographic distributions are as follows (**Supplemental Figure 2**): Gender: Female: 58.3%, Male 41.7%; Race: Asian 7.9%, Black: 12.4%, White: 79.6%. Ethnicity: Non-Hispanic: 96.2%, Hispanic: 3.8%; Preferred Language: English: 91.4%, Spanish: 1.7%, Others: 6.9%; Marital Status: Married or Partnered: 55.0%, Single: 25.1%, Divorced: 7.0%, Legally Separated: 0.9%, Widowed: 8.2%, Unknown: 3.7%. Each subject was categorized into two classes, including non-vision-threatening DR and vision-threatening DR. Vision-threatening DR comprises severe non-proliferative diabetic retinopathy (NPDR) and proliferative diabetic retinopathy (PDR), while non-vision-threatening DR comprises normal, mild, and moderate NPDR. The diagnosis information of DR or non-DR was extracted from the International Classification of Diseases (ICD) codes in the patient’s electronic health records. For color fundus, 95.3% and 4.7% of patients were identified as non-vision-threatening DR and vision-threatening DR, respectively. For SLO fundus and OCT B-Scans, 97.7% and 2.3% of patients were identified as non-vision-threatening DR and vision-threatening DR, respectively.

#### Harvard-FairVision30k

This is a public dataset proposed to study the fairness issue in eye disease screening, including SLO fundus images and OCT B-Scans collected from 10,000 patients for DR detection.^20^ The average age was 64.5 ± 16.5 years. The demographic distributions are as follows (**Supplemental Figure 3**): Gender: Female: 55.5%, Male 44.5%; Race: Asian 7.6%, Black: 14.6%, White: 77.8%. Ethnicity: Non-Hispanic: 96.1%, 3.9% Hispanic; Preferred Language: English: 90.9%, Spanish: 2.0%, Others: 7.1%; Marital Status: Married or Partnered: 54.0%, Single: 25.2%, Divorced: 7.1%, Legally Separated: 1.0%, Widowed: 8.4%, Unknown: 4.3%. Of the patients, 90.9% were identified as non-vision-threatening DR and 9.1% as vision-threatening DR.

#### ODIR-5K

This is a public dataset proposed to study eye disease screening using color fundus images.^21^ After processing, 6,392 fundus images were collected from 3,358 patients, with an average age of 57.9 ± 11.7 years. Gender is available in this dataset, where Females and Males account for 46.4% and 53.6% of the subjects, respectively. Of the subjects, 66.8% were identified as non-DR and 33.2% as DR.

### Equitable deep learning model with fair adaptive scaling

We aimed to devise a fairness learning module to enhance existing deep learning models to achieve equitable DR detection performance across different identity groups. The model takes an image as input to predict the binary DR and non-DR category while considering the associated identity attributes (e.g., gender, race, and ethnicity) of the input image. The goal is to maximize the overall DR detection accuracy of all samples while minimizing the discrepancies across different identity groups. We proposed a Fair Adaptive Scaling (FAS) module (**Figure 2**), which can be integrated with existing models to improve model performance equity. FAS employs learnable group weights and past individual loss data to adjust the loss function during the current training batch. The idea is that samples that had higher group weights and individual loss values in the prior batch will be given more weight in the current batch’s loss function. Note that the group weight is a learnable parameter dynamically updated instead of an empirically selected fixed value. This approach combines both group and individual scaling not only to address fairness at the group level but also manage within-group sample variations. This is done to avoid issues that may arise if only group scaling is used, as it could overly weight or underweight most samples within a group due to isolated outliers, consequently deteriorating the model. FAS can be integrated with state-of-the-art 2D or 3D deep learning models for DR detection by taking the identity information into account. In this work, we adopted EfficientNet^22^ and ViT-B^23^ as backbones to validate the effectiveness of FAS, namely EfficientNet + FAS and ViT-B + FAS, as they performed the best in most comparisons. Specifically, for the 2D model, it takes a batch of color fundus or SLO fundus images together with identity attributes as inputs. The backbone model is used to extract features from the images, which are subsequently used for the binary (i.e. DR and non-DR) classification. During the supervised training, each sample’s contribution is dynamically scaled by the learnable group and individual weight values. Such a mechanism allows naturally underrepresented samples to adjust themselves to obtain equal importance during the model training.

**Figure 2.**
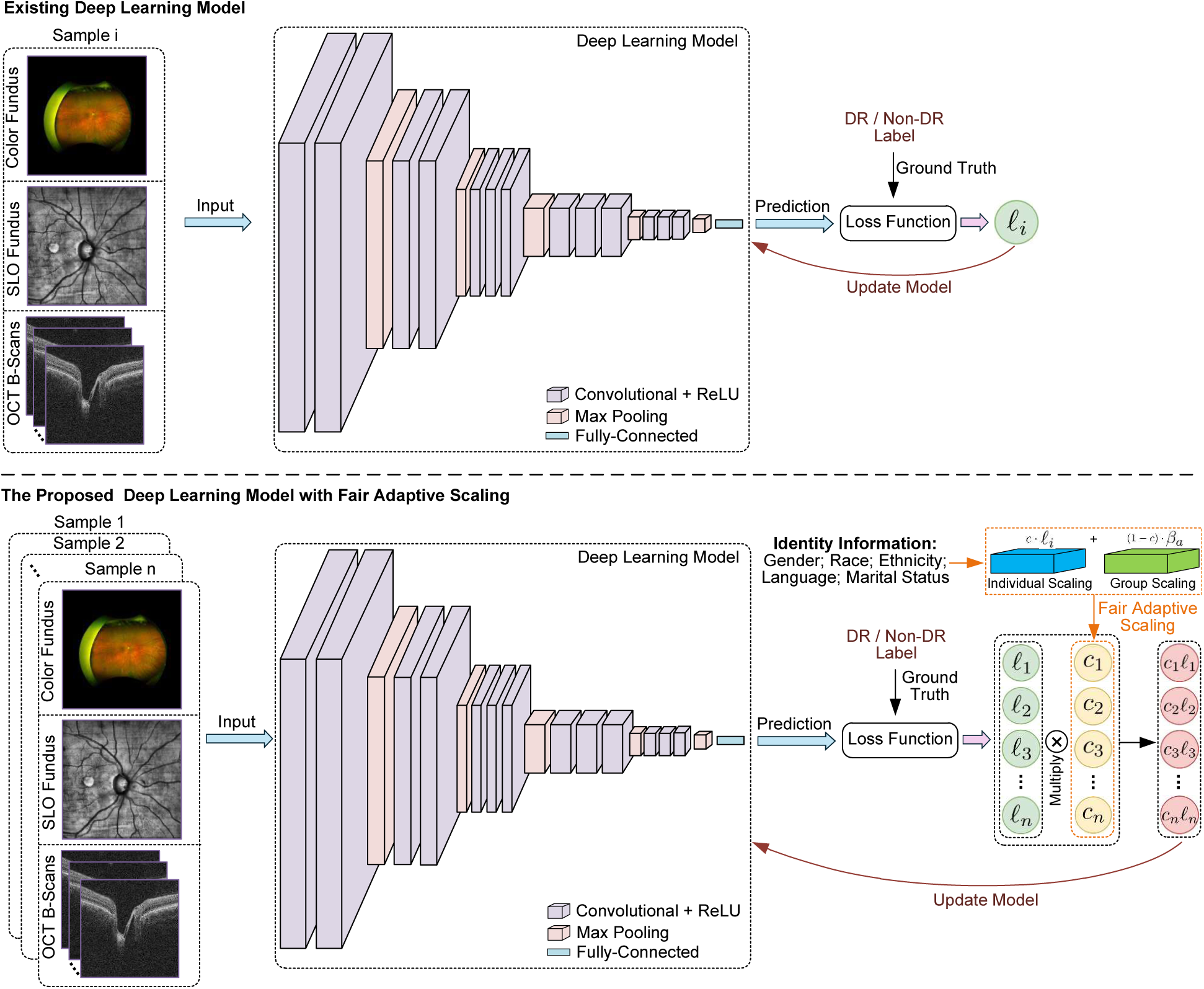
The comparison of existing deep learning model and the proposed model with fair adaptive scaling. Existing deep models learn features from the fundus image or OCT B-Scans for DR detection without considering the identity information. In contrast, the proposed model with fair adaptive scaling leverages the identity information to guide the model dynamically adjust the contributions of individual samples, thus achieving equitable DR detection across different identity groups.

To handle 3D OCT B-Scans, we adopted two types of deep learning backbones combined with the FAS. The first type of backbone is adapted from the 2D models EfficientNet^22^ and ViT-B^23^ by adding a mapping initial layer to transform 200-channel OCT images into corresponding 3-channel image, while the remaining learning architectures remain unchanged. The second type of backbone is the 3D versions of ResNet18 and DenseNet121, which feature 3D convolutions dedicated to 3D medical images.^24^

### Baseline models for comparison

We selected seven state-of-the-art baseline deep learning models to compare DR detection performance and performance equity across different identity groups. These models are VGG-16^25^, Swin-B^26^, ResNet^27^, ConvNeXt^28^, DenseNet^29^, EfficientNet^22^, and ViT-B^23^, all of which have been widely used for processing medical images. For these comparative methods, we used a consistent training pipeline, similar to that of the proposed models in this work. A validation dataset was used to tune the hyperparameters for individual models to achieve a competitive performance. In addition, we included respective variants of the above models by introducing an adversarial training loss or using a data oversampling process. Both techniques are deemed useful in mitigating performance disparities between subgroups in prior research.^30–32^ In particular, adversarial training forces the model not to learn identity-specific information from the images, thus avoiding the performance bias caused by identity information, while oversampling addresses data imbalance issue. We also investigated the transfer learning technique with EfficientNet and ViT-B models to further optimize the performance for each specific identity group.^33,34^ In this approach, a global model was first trained using all available training data. Subsequently, the model was fine-tuned for each individual identity group using the group-specific training data.

### Evaluation metrics and statistical analysis

Statistical analyses were performed in Python 3.8 (available at http://www.python.org) on a Linux system. The scikit-learn package was used to calculate the area under the receiver operating characteristic curve (AUC). To account for the potential tradeoff between overall AUC and group disparity, we proposed a new metric called equity-scaled AUC (ES-AUC) to compare model performance equity. The ES-AUC is defined as the overall AUC divided by one plus the sum of the absolute differences between the overall AUC and each group’s AUC, formulated as 𝑨𝑼𝑪_𝑬𝑺_ = 𝑨𝑼𝑪_𝒐𝒗𝒆𝒓𝒂𝒍𝒍_/(𝟏 + 𝚺l𝑨𝑼𝑪_𝒐𝒗𝒆𝒓𝒂𝒍𝒍_ −𝑨𝑼𝑪_𝒈𝒓𝒐𝒖𝒑_l). We used t-test and bootstrapping with replacement to compare the DR screening AUC and ES-AUC of different deep learning models, with or without FAS. The basic idea of bootstrapping is that inference about a population from sample data can be modeled by resampling the sample data and performing inference about a sample from resampled data. This method is a straightforward way to derive estimates of standard errors and confidence intervals. All statistical tests were two-sided, and p < 0.05 was considered to indicate a statistically significant result. We also calculated the overall sensitivity performance and corresponding ES-sensitivity at the thresholds of 0.9 and 0.95 specificities, which were reported in the Supplemental Material due to page limits.

## Results

### Results for color fundus images

For racial attribute, ViT-B achieved the best overall AUC of 0.90 and ES-AUC of 0.84 among seven state-of-the-art baseline deep learning models, followed by the EfficientNet with its overall AUC and ES-AUC being 0.88 and 0.83, respectively (**Figure 3a**). Data oversampling and adversarial training significantly improved the overall AUC performances of VGG, ResNet and ConvNeXt (p < 0.05), but were not shown useful for other models (**Supplemental Figure 4**). With transfer learning, EfficientNet significantly improved the AUC performances for Asians and Whites up to 0.04 (p < 0.01) and 0.01 (p < 0.05) respectively (**Figure 3b**). After integrating with FAS, the overall AUC and ES-AUC of EfficientNet improved from 0.88 and 0.83 to 0.90 and 0.84 (p < 0.05), where the AUCs for Asians and Whites improved by 0.04 and 0.03, respectively (p < 0.01, **Figure 3b**). Similarly, with FAS, the AUCs of ViT-B for Asians and Whites improved by 0.02 and 0.01 (p < 0.05), respectively. For the gender attribute, ViT-B and EfficientNet remained the best performing baseline models with both ES-AUC being 0.88 (**Figure 4a**). Adversarial training boosted the AUC of EfficientNet for Females by 0.02 (p < 0.01), while the oversampling and transfer learning did not bring significant AUC improvements for either EfficientNet and ViT-B (**Figures 4b and 4c**). With FAS, the overall AUC and ES-AUC of EfficientNet both improved by 0.01 (p < 0.05), while the AUC and ES-AUC for ViT-B increased by 0.02 and 0.03 (p < 0.01) respectively. For ethnic attribute, oversampling, transfer learning and adversarial training could not enhance EfficientNet and ViT-B (**Figures 5b and 5c**). In contrast, the overall AUC and AUC of EfficientNet with FAS for non-Hispanic group both improved by 0.01(p < 0.05, **Figure 5b**). The overall AUC, ES-AUC and AUC of ViT-B with FAS for Hispanic group all increased by 0.01 (p < 0.05, **Figure 5c**).

**Figure 3.**
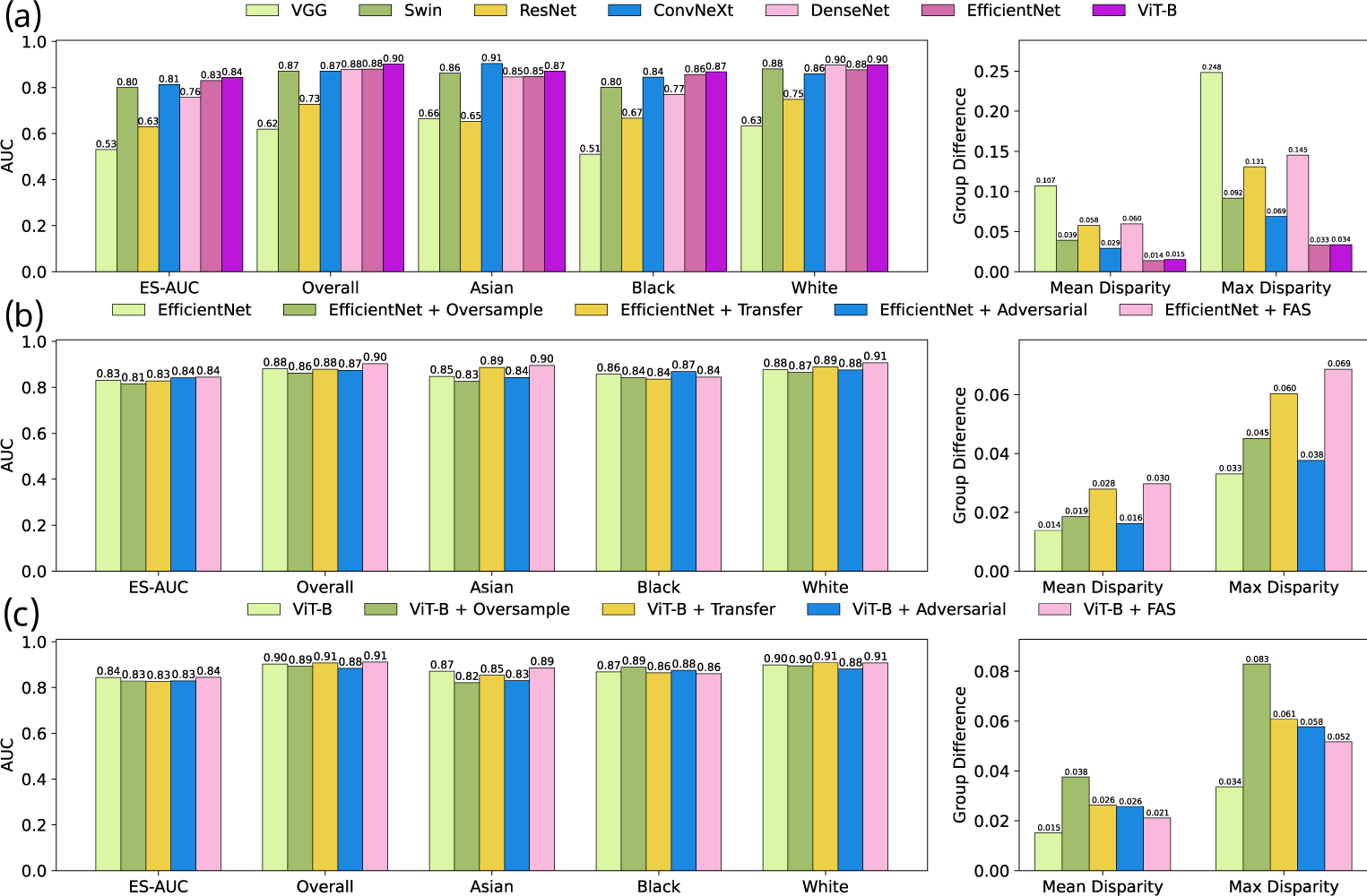
Results on Color Fundus Images on race attributes of the In-house Dataset. (a) The accuracy of various baseline models. (b) The accuracy of EfficientNet and its integration with oversampling, adversarial, transfer learning and our FAS techniques. (c) The accuracy of ViT-B and its integration with oversampling, adversarial, transfer learning and our FAS techniques.

**Figure 4.**
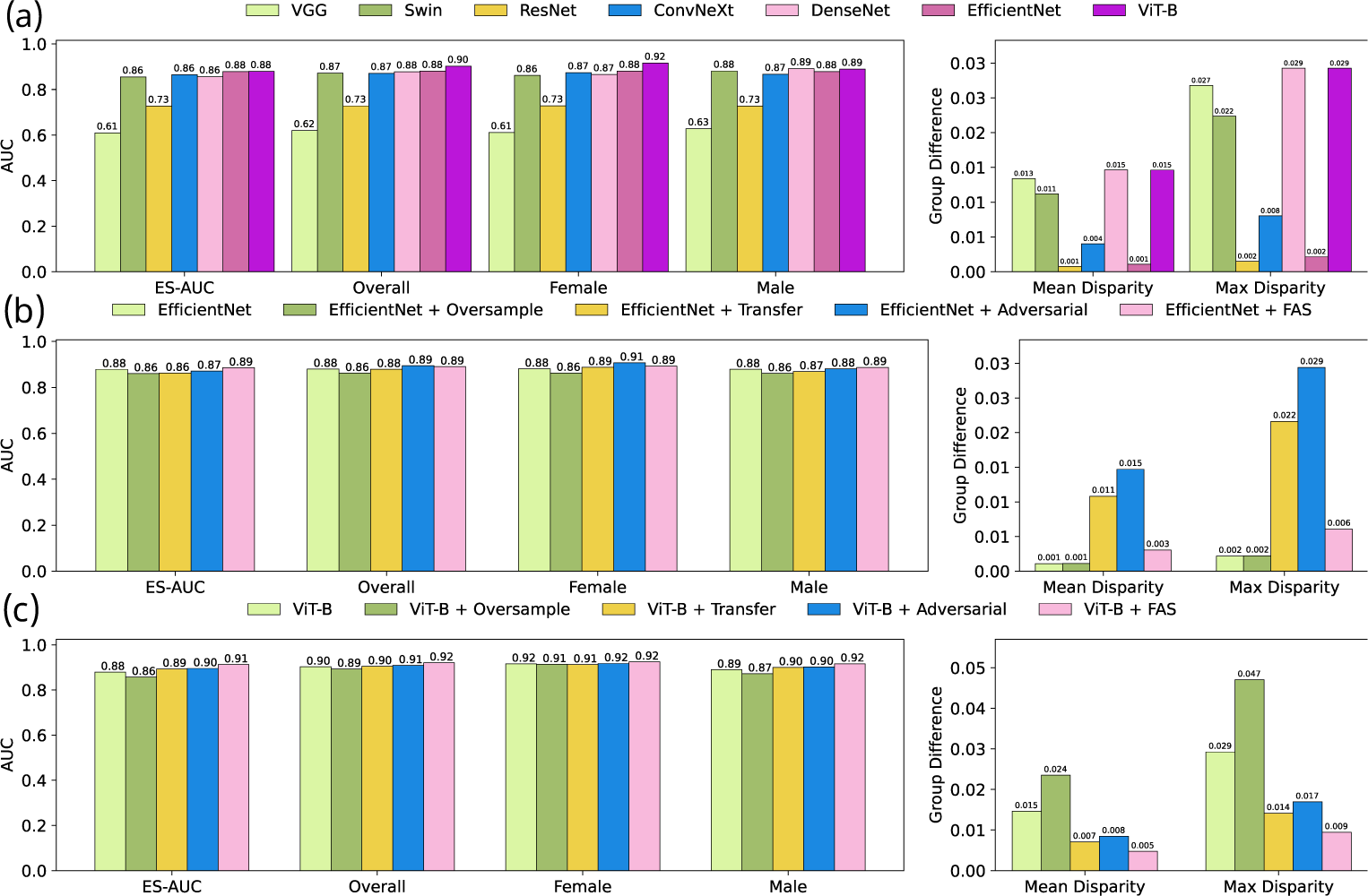
Results on Color Fundus Images on gender attributes of the In-house Dataset. (a) The accuracy of various baseline models. (b) The accuracy of EfficientNet and its integration with oversampling, adversarial, and our FAS techniques. (c) The accuracy of ViT-B and its integration with oversampling, adversarial, transfer learning and our FAS techniques.

**Figure 5.**
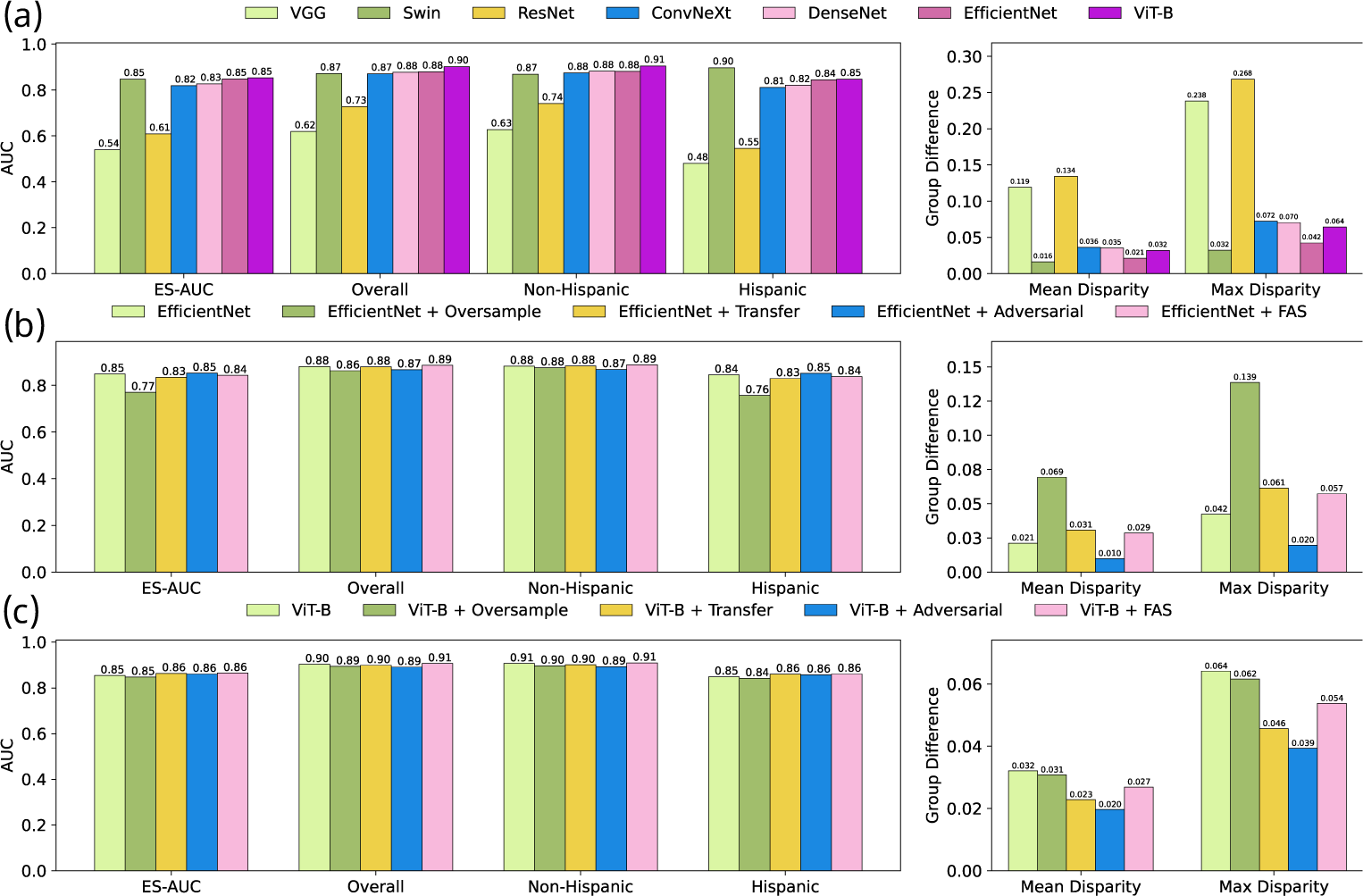
Results on Color Fundus Images on ethnicity attributes of the In-house Dataset. (a) The accuracy of baseline models. (b) The accuracy of EfficientNet and its integration with oversampling, adversarial, transfer learning and our FAS techniques. (c) The accuracy of ViT-B and its integration with oversampling, adversarial, transfer learning and our FAS techniques.

Similar results can be observed when using ODIR-5K dataset for the gender attribute. With FAS, the overall AUC and ES-AUC both improved by 0.01 (p < 0.05, **Supplemental Figure 8**). The overall AUC and ES-AUC for ViT-B improved by 0.03 and 0.02 respectively, after integrating the FAS (p < 0.01, **Supplemental Figure 8**). The AUCs for Females and Males improved from 0.75 and 0.76 to 0.78 and 0.78, respectively.

### Results for SLO fundus images

Using in-house MEE dataset on the racial attribute, ViT-B achieved the highest overall AUC of 0.82, while Swin-B achieved the highest ES-AUC of 0.77 (**Figure 6a**). In general, the strategies of oversampling, transfer learning and adversarial training could not improve the overall AUC and ES-AUC performances for both EfficientNet and ViT-B (**Figures 6b and 6c**). In contrast, with FAS, the overall AUC of EfficientNet significantly improved from 0.80 to 0.83 (p < 0.01), where the AUCs for Asians, Blacks and Whites improved by 0.02, 0.01 and 0.04, respectively (p < 0.05, **Figures 6b**). The overall AUC and ES-AUC of ViT-B with FAS increased from 0.82 and 0.71 to 0.84 and 0.75, respectively. In subgroups, the AUCs for Asians, Blacks and Whites significantly improved by 0.02, 0.03, and 0.02, respectively (p < 0.01, **Figure 6c**). On the gender attribute, conventional strategies such as oversampling, transfer learning, and adversarial training strategies failed to boost model performance and equity, while FAS significantly boosted EfficientNet and ViT-B (**Figures 7b and 7c**). Specifically, FAS improved EfficientNet’s overall AUC and ES-AUC by 0.02 (p < 0.01), where the same improvement of 0.02 was achieved for Females and Males (p < 0.01, **Figures 7b**). Similarly, with FAS, the overall AUC and ES-AUC of ViT-B improved by 0.02 and 0.01, where Females and Males had improvements of 0.03 and 0.02, respectively (p < 0.05, **Figure 7c**). On the ethnic attribute, after integrating FAS, the overall AUC and ES-AUC of EfficientNet improved by 0.02 and 0.04, respectively (p < 0.01, **Figure 8b**). The AUC for non-Hispanic group improved 0.02, but no improvement was observed for the Hispanic group (**Figure 8b**). With FAS, the overall AUC of ViT-B improved from 0.82 to 0.84, where the non-Hispanic group improved by 0.03 (p < 0.01, **Figure 8c**), although no improvement was observed for the Hispanic group.

**Figure 6.**
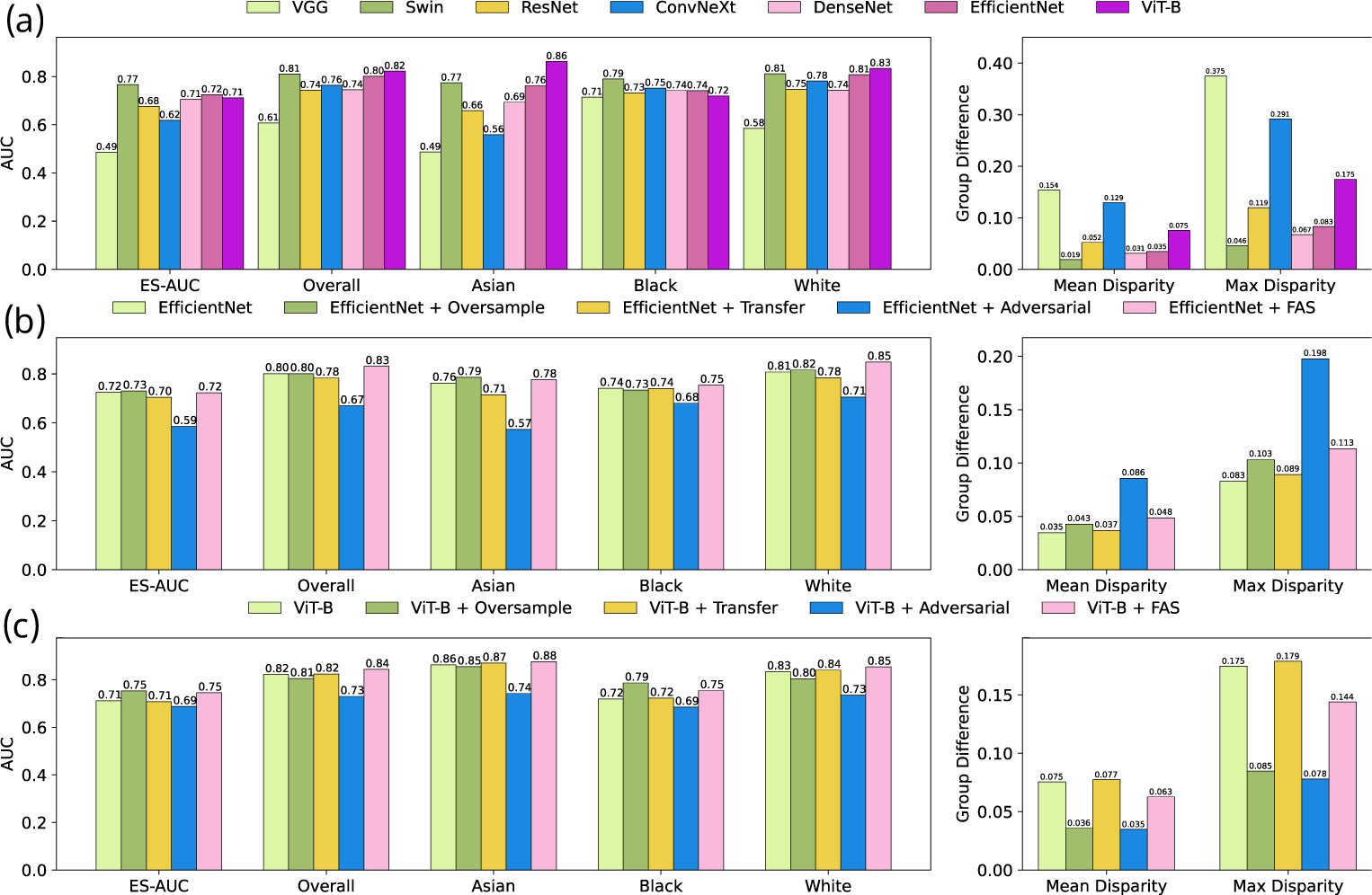
Results on SLO Fundus Images on race attributes of the In-house Dataset. (a) The accuracy of various baseline models. (b) The accuracy of EfficientNet and its integration with oversampling, adversarial, transfer learning and our FAS techniques. (c) The accuracy of ViT-B and its integration with oversampling, adversarial, transfer learning and our FAS techniques.

**Figure 7.**
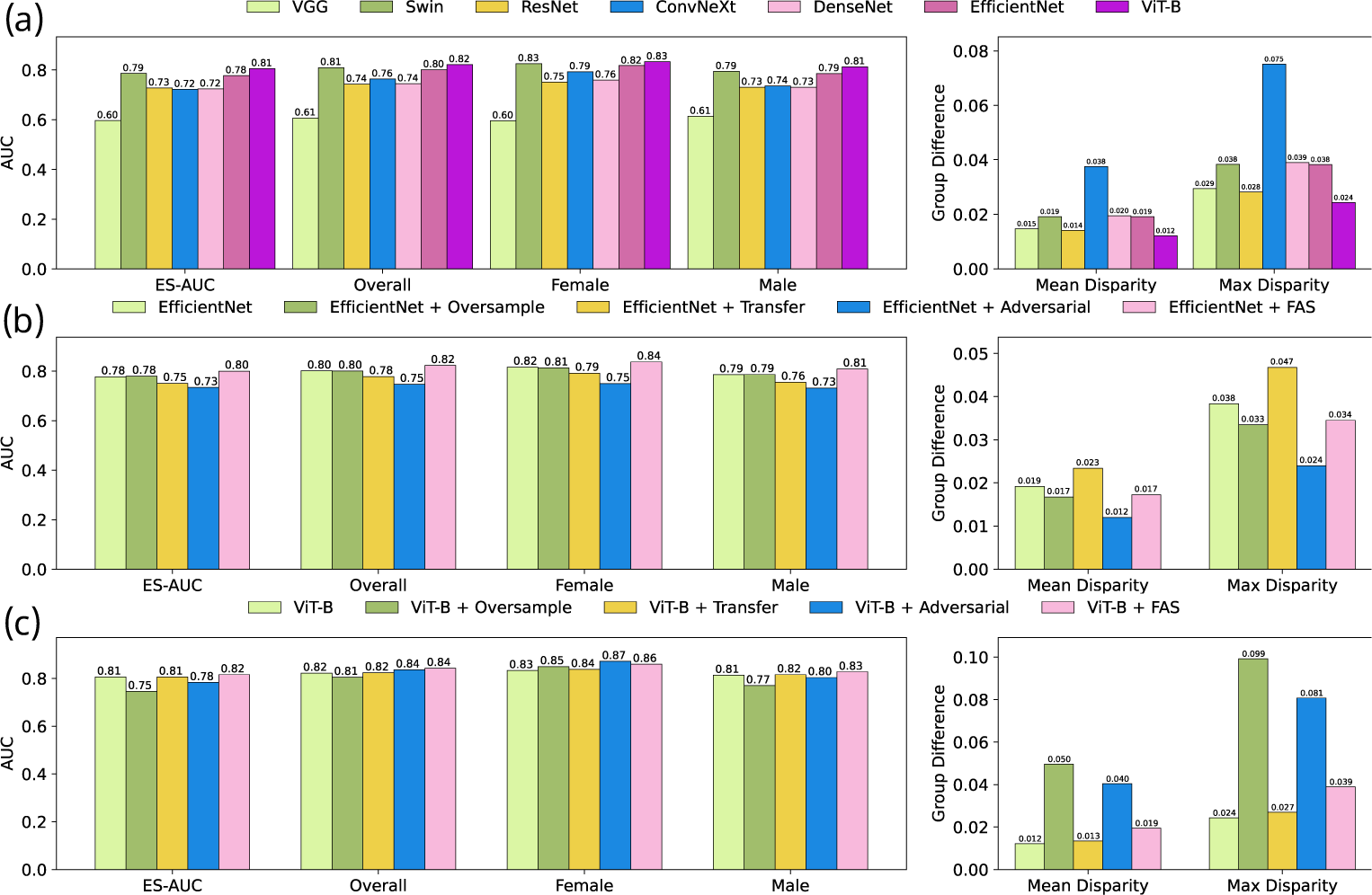
Results on SLO Fundus Images on gender attributes of the In-house Dataset. (a) The accuracy of various baseline models. (b) The accuracy of EfficientNet and its integration with oversampling, adversarial, transfer learning and our FAS techniques. (c) The accuracy of ViT-B and its integration with oversampling, adversarial, transfer learning and our FAS techniques.

**Figure 8.**
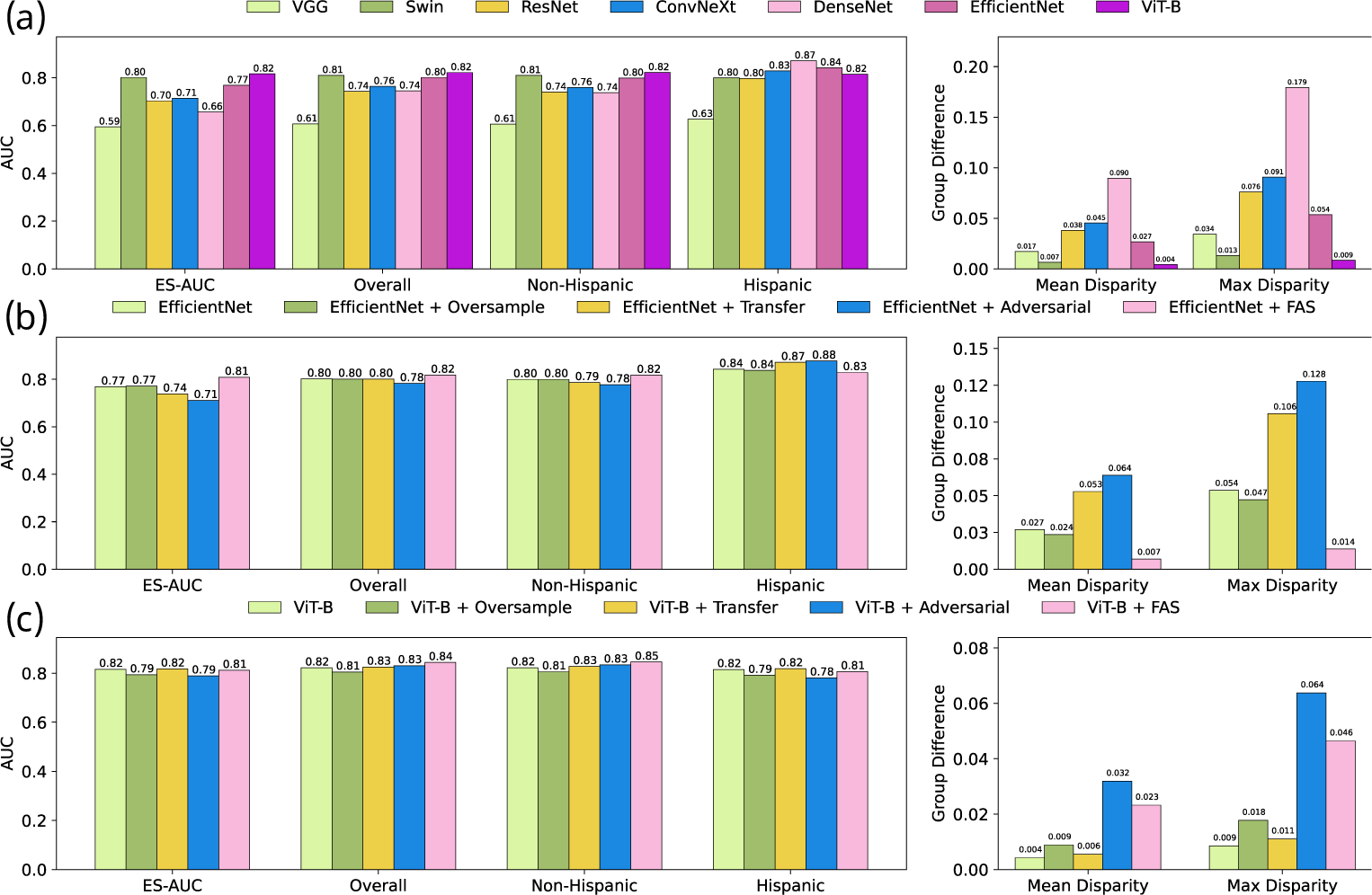
Results on SLO Fundus Images on ethnicity attributes of the In-house Dataset. (a) The accuracy of various baseline models. (b) The accuracy of EfficientNet and its integration with oversampling, adversarial, transfer learning and our FAS techniques. (c) The accuracy of ViT-B and its integration with oversampling, adversarial, transfer learning and our FAS techniques.

On the Harvard-FairVision30k dataset, FAS was also shown to be effective in boosting the overall AUC performance and reducing group performance disparities. For example, on the racial attribute, the AUC and ES-AUC of EfficientNet with FAS improved from 0.79 and 0.67 to 0.81 and 0.74, respectively. Notably, significant AUC improvements (0.04 and 0.07) were achieved for Asians and Blacks, respectively (p < 0.01, **Supplemental Figure 13**). Similarly, the performance disparities were significantly improved for ViT-B after integrating the FAS, with the ES-AUC and AUCs for Asians and Blacks all improved by 0.02 (p < 0.01).

### Results for OCT B-Scans

DenseNet121 and ResNet18, based on 3D convolutions with or without integrating the FAS, were evaluated on Race, Gender, and Ethnicity. Compared with DenseNet121 on the racial attribute using in-house OCT B-Scans, DenseNet121 + FAS improved the overall AUC and ES-AUC from 0.875 and 0.81 to 0.890 and 0.83, respectively (p < 0.01, **Table 1**), where the AUCs for Asians and Blacks improved by 0.032 and 0.02. Similarly, for Resnet18 with FAS, the overall AUC and ES-AUC both improved by 0.012 (p < 0.05, **Table 1**), with a more prominent AUC improvement for Asians (0.026) compared with Blacks (0.011) and Whites (0.011). On the gender attribute, FAS improved the overall AUC and ES-AUC of DenseNet121 by 0.044 and 0.027, where the AUCs for Females and Males improved by 0.054 and 0.035, respectively (p < 0.01, **Table 1**). After integrating FAS with Resnet18 on Gender, the overall AUC and ES-AUC significantly increased from 0.872 and 0.856 to 0.903 and 0.882, respectively. On the ethnic attribute, the overall AUC for DenseNet121 integrating the FAS improved by 0.019, although the ES-AUC showed no improvement (**Table 1**). The overall AUC and ES-AUC for ResNet18 + FAS improved over ResNet18 by 0.022 and 0.062, respectively, with the non-Hispanic group improving from 0.87 to 0.904, and Hispanic groups remaining nearly unchanged (**Table 1**).

**Table 1.** Experimental results on in-house OCT B-Scans using 3D deep learning models.

| Race | ES-AUC | Overall AUC | Asian | Black | White | Mean Disparity | Max Disparity |
| --- | --- | --- | --- | --- | --- | --- | --- |
| DenseNet121 | 0.810 | 0.875 | 0.814 | 0.893 | 0.875 | 0.047 | 0.090 |
| Resnet18 | 0.818 | 0.872 | 0.903 | 0.897 | 0.863 | 0.025 | 0.046 |
| DenseNet121 + FAS | 0.830 | 0.890 | 0.846 | 0.913 | 0.886 | 0.038 | 0.075 |
| Resnet18 + FAS | 0.820 | 0.884 | 0.929 | 0.908 | 0.874 | 0.031 | 0.062 |
| Gender | ES-AUC | Overall AUC | Female | Male | Mean Disparity | Max Disparity |  |
| DenseNet121 | 0.843 | 0.875 | 0.890 | 0.853 | 0.030 | 0.042 |  |
| Resnet18 | 0.856 | 0.872 | 0.879 | 0.860 | 0.015 | 0.022 |  |
| DenseNet121 + FAS | 0.870 | 0.919 | 0.944 | 0.888 | 0.043 | 0.061 |  |
| Resnet18 + FAS | 0.882 | 0.903 | 0.910 | 0.887 | 0.018 | 0.025 |  |
| Ethnicity | ES-AUC | Overall AUC | Non-Hispanic | Hispanic | Mean Disparity | Max Disparity |  |
| DenseNet121 | 0.846 | 0.875 | 0.876 | 0.843 | 0.027 | 0.038 |  |
| Resnet18 | 0.835 | 0.872 | 0.870 | 0.913 | 0.035 | 0.049 |  |
| DenseNet121 + FAS | 0.823 | 0.894 | 0.899 | 0.812 | 0.069 | 0.097 |  |
| Resnet18 + FAS | 0.897 | 0.904 | 0.904 | 0.912 | 0.006 | 0.009 |  |

On the Harvard-FairVision30K dataset, we can observe consistent improvement after integrating FAS with DenseNet121 and ResNet18 (**Table 2**). On the racial attribute, FAS improved the overall AUC of DenseNet121 by 0.01 and ES-AUC by 0.055, with significant AUC improvements of 0.051 and 0.024 for Asians and Blacks, respectively (p < 0.01, **Table 2**). For DenseNet18 + FAS, the overall AUC and ES-AUC improved by 0.023 and 0.047, respectively. The AUCs for Blacks significantly improved from 0.825 to 0.879. On the gender attribute, the improvements in model performance and disparity were marginal for DenseNet121 after integrating with FAS. In contrast, with FAS, DenseNet18’s overall AUC and ES-AUC significantly improved by 0.017 and 0.02 (p < 0.01, **Table 2**). For the ethnic attribute, the overall AUC DenseNet18 after integrating the FAS improved from 0.876 to 0.895, while the improvement for DenseNet18 + FAS was not significant.

**Table 2.** Experimental results on Harvard-FairVision30k OCT B-Scans using 3D deep learning models.

| Race | ES-AUC | Overall AUC | Asian | Black | White | Mean Disparity | Max Disparity |
| --- | --- | --- | --- | --- | --- | --- | --- |
| DenseNet121 | 0.812 | 0.914 | 0.849 | 0.869 | 0.929 | 0.046 | 0.087 |
| Resnet18 | 0.805 | 0.876 | 0.855 | 0.825 | 0.893 | 0.039 | 0.078 |
| DenseNet121 + FAS | 0.867 | 0.924 | 0.900 | 0.893 | 0.934 | 0.024 | 0.044 |
| Resnet18 + FAS | 0.852 | 0.889 | 0.858 | 0.879 | 0.892 | 0.019 | 0.038 |
| Gender | ES-AUC | Overall AUC | Female | Male | Mean Disparity | Max Disparity |  |
| DenseNet121 | 0.904 | 0.914 | 0.916 | 0.905 | 0.009 | 0.012 |  |
| Resnet18 | 0.862 | 0.876 | 0.864 | 0.880 | 0.013 | 0.018 |  |
| DenseNet121 + FAS | 0.909 | 0.919 | 0.921 | 0.911 | 0.008 | 0.011 |  |
| Resnet18 + FAS | 0.882 | 0.893 | 0.883 | 0.896 | 0.010 | 0.015 |  |
| Ethnicity | ES-AUC | Overall AUC | Non-Hispanic | Hispanic | Mean Disparity | Max Disparity |  |
| DenseNet121 | 0.910 | 0.914 | 0.914 | 0.910 | 0.003 | 0.004 |  |
| Resnet18 | 0.864 | 0.876 | 0.876 | 0.862 | 0.011 | 0.016 |  |
| DenseNet121 + FAS | 0.897 | 0.919 | 0.917 | 0.941 | 0.018 | 0.026 |  |
| Resnet18 + FAS | 0.857 | 0.895 | 0.892 | 0.937 | 0.036 | 0.050 |  |

## Discussion

As deep learning models are widely used for automatic disease screening, it is essential to investigate existing models and pursue equitable model performance across different identity groups. In this work, we thoroughly examined seven state-of-the-art deep learning models for DR detection and analyzed their performance disparities across diverse identity groups. Three different datasets encompassing 2D color fundus, SLO fundus, and 3D OCT B-Scans were included in the evaluation. The experimental results demonstrated that existing deep models generally suffer from the performance disparities, which are particularly unfair for underrepresented groups. For example, for the racial attribute on the in-house SLO fundus images, ViT-B achieved the lowest AUC score of 0.72 for Blacks, compared to 0.83 for Whites (**Figure 6a**). We proposed a Fair Adaptive Scaling (FAS) module to enhance existing model performance and mitigate model performance disparities across different identity groups. We demonstrated the effectiveness of FAS through its integration with EfficientNet and ViT-B and compared it with three conventional strategies for reducing group performance disparities, including data oversampling, transfer learning, and adversarial training. FAS proved helpful in boosting model performance in DR detection as well as reducing subgroup performance disparities. For example, for the racial attribute on the in-house SLO fundus images, ViT-B combined with FAS improved the AUCs for Blacks and Whites from 0.72 to 0.83 to 0.75 and 0.85, respectively (F**igure 6c**). As a result, the corresponding ES-AUC improved from 0.71 to 0.75, indicating that the ViT-B model has become fairer for subgroup DR detection after integrating with FAS.

The major idea of FAS is to dynamically learn the contribution of each individual sample for DR detection, conditioned on the associated identity attributes. This is achieved by employing learnable group weights (i.e. group scaling) and past individual loss data (i.e. individual scaling) to adjust the loss function during the current training batch. Essentially, samples that had higher group weights and individual loss values in the prior batch will be given more weight in the current batch’s loss function. This approach of combining both group and individual scaling is taken to not only address fairness at a group level but also manage within-group sample variations. FAS can affect the way the model learns features from the input image sample in order to achieve improved model performance and reduce group disparities (**Figure 9**). The distribution of features learned by the existing deep learning model was highly indistinguishable across different identity groups and centralized (**Figures 9a and 9b**). In contrast, the distribution of features from the deep learning model with FAS had clearer boundaries and was more spread out in the feature space. Such a reformed feature distribution, incurred by FAS, may have contributed to the improvement of overall model performance and reduced group disparities in DR detection. Compared with conventional strategies of data oversampling, transfer learning and adversarial training for mitigating group disparities, FAS was able to demonstrate superior effectiveness and robustness for different identity attributes for all three datasets used in the evaluation.

**Figure 9.**
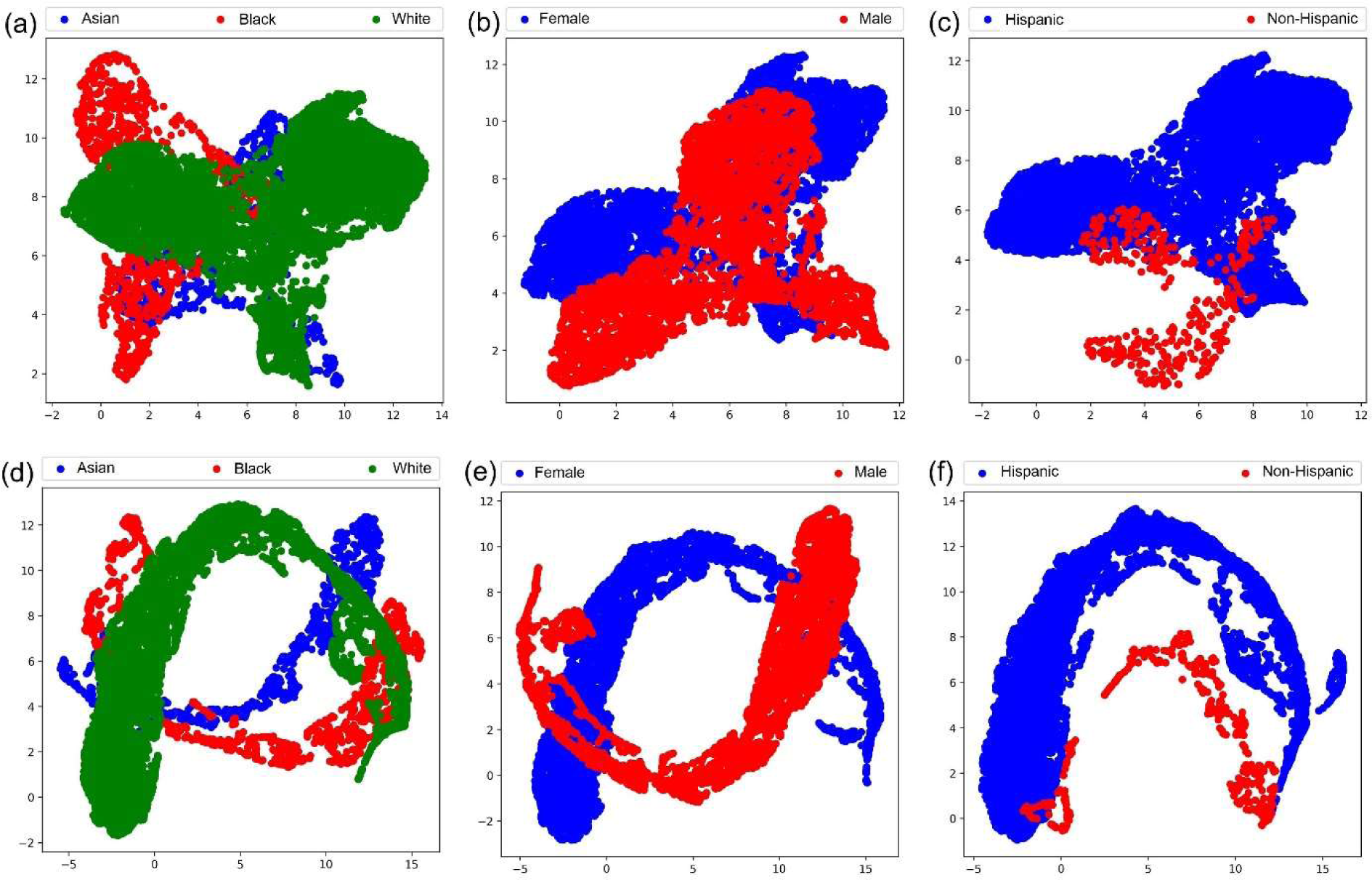
The distribution of features learned from in-house SLO fundus images by the existing baseline EfficientNet model and the EfficientNet + FAS model. (a) EfficientNet on Race. (b) EfficientNet on Gender. (c) EfficientNet on Ethnicity. (d) EfficientNet + FAS on Race. (e) EfficientNet + FAS on Gender. (f) EfficientNet + FAS on Ethnicity.

Our study had several limitations. **First**, the proposed deep learning models with FAS did not consistently improve the overall AUC performance and group disparities quantified by ES-AUC, max disparity, and mean disparity for all five sensitive attributes including Race, Gender, Ethnicity, Language, and Marital Status. For example, for in-house color fundus images, the overall AUC and ES-AUC of EfficientNet (**Figures 3b and 4b**) and ViT-B (**Figures 3c and 4c**) showed significant improvements on Race and Gender after integrating with FAS but did not show improvements on Language (**Supplemental Figures 7d and 7f**). A possible reason is that retinal images from different subgroups, identified by certain sensitive attributes (e.g., Language), present considerable structural variance, meaning that one subgroup may contain more hard classification cases than another. While the model is trained to pursue global classification accuracy, this could compromise the accuracy for certain subgroups. Therefore, sophisticated strategies need to be designed to explicitly balance the accuracy across different identity groups. **Second**, the experimental results demonstrated that ES-AUC, mean and max disparity metrics were inconsistent for comparing model equity. In this work, the ES-AUC is treated as a more comprehensive equity measurement than mean and max disparities, given that mean and max disparities do not fully considered the variance of subgroup performances. Additionally, other fairness metrics such as demographic parity, equalized odds, and equal opportunity can also be adopted. **Third**, we have thoroughly validated how fair the model would be regarding data sample size for different sensitive attributes. The data sample sizes involved in this study were relatively large, which could bias the model performance and equity. However, we tested the influence of sample sizes using the in-house color fundus images on Race, Gender, and Ethnicity. The experiments demonstrated that the issue of model inequity existed at different scales of data samples, and the proposed deep learning model with FAS helped to mitigate model performance disparities across different identity groups (**Supplemental Figures 24-26**). Lastly, we have not fully explored the efficacy of FAS when paired with other supervised deep learning models like the Swin network and unsupervised deep learning models like the masked autoencoder, even though FIN has the versatility to be paired with various learning frameworks.

In conclusion, we proposed a FAS module to promote model performance equity for DR detection. FAS is an independent module that can be integrated into many existing deep learning models to improve model fairness across different identity groups. Extensive experiments using three different datasets for DR detection and comparisons with conventional fairness learning strategies demonstrated the effectiveness of FAS in boosting both overall model performance and group performance disparities, especially for underrepresented groups.

## Data Availability

The Harvard-FairVision30k dataset is available through the public link https://ophai.hms.harvard.edu/datasets/harvard-fairvision30k and was used with approvals. The ODIR-5K dataset is publicly available at https://www.kaggle.com/datasets/andrewmvd/ocular-disease-recognition-odir5k. The in-house data were provided by the Massachusetts Eye and Ear (MEE). The institutional review boards (IRB) of MEE approved the creation of the database in this retrospective study.

## Notes

### Competing Interest Statement

The authors have declared no competing interest.

### Funding Statement

This work was supported by NIH R00 EY028631, NIH R21 EY035298, Research To Prevent Blindness International Research Collaborators Award, and Alcon Young Investigator Grant.

### Summary of Updates

The structure and format of the paper have been improved.

