## Supplemental Material for "Equitable deep learning for diabetic retinopathy detection using multi-dimensional retinal imaging with fair adaptive scaling: a retrospective study"

### Supplementary Material

The supplementary material for the manuscript entitled “Equitable deep learning for diabetic retinopathy detection using multi-dimensional retinal imaging with fair adaptive scaling: a retrospective study”

#### Data characteristics: Inhouse Color Fundus

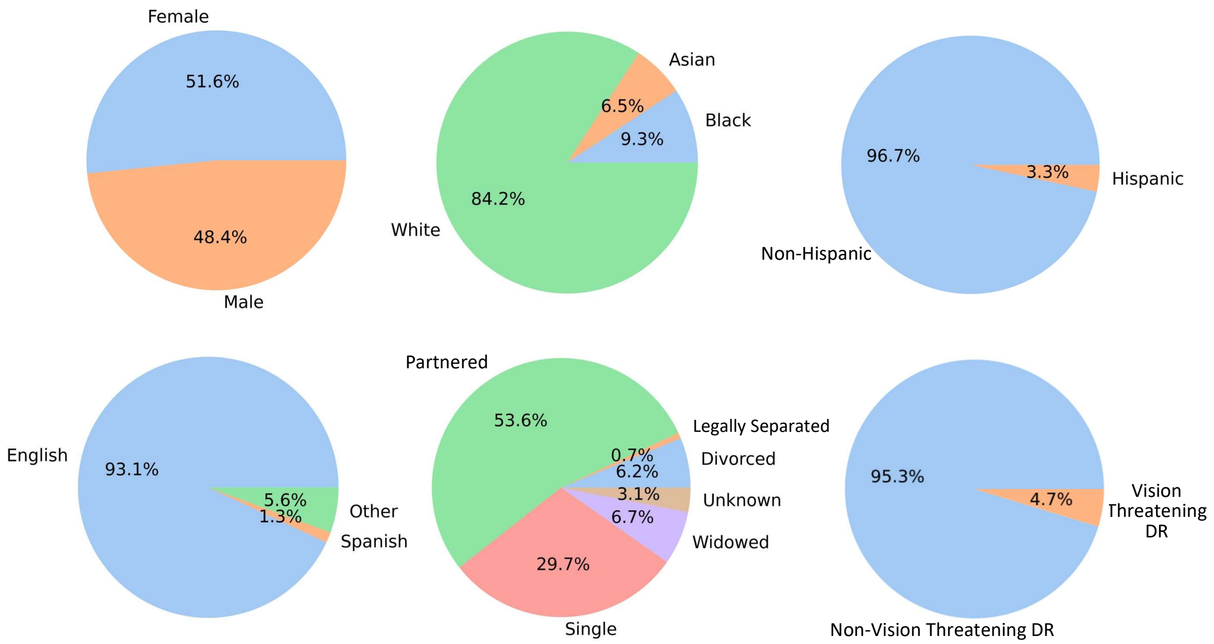


**Figure 1. The demographic distributions of the in-house color fundus dataset for gender, race, ethnicity, preferred language, marital status, and legally separated status.**

#### Data characteristics: Inhouse SLO Fundus + OCT B-scans

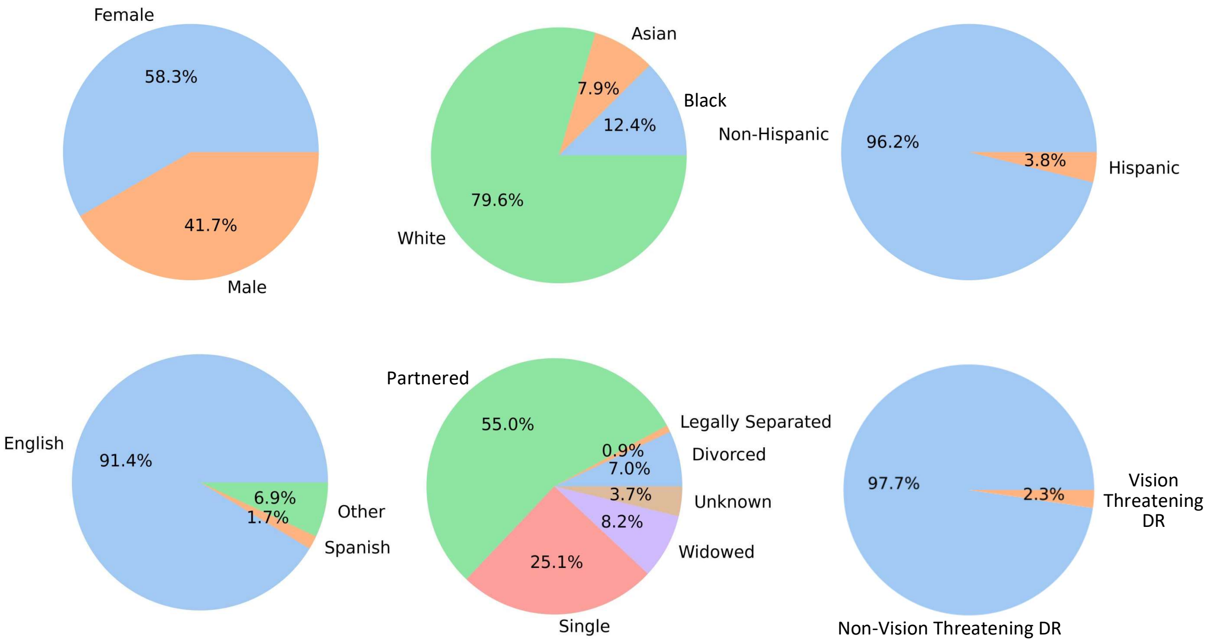


**Figure 2. The demographic distributions of the In-house SLO fundus images and OCT B-scans dataset for gender, race, ethnicity, preferred language, marital status, and legally separated status.**

#### Data characteristics: Haravrd-FairVision30K

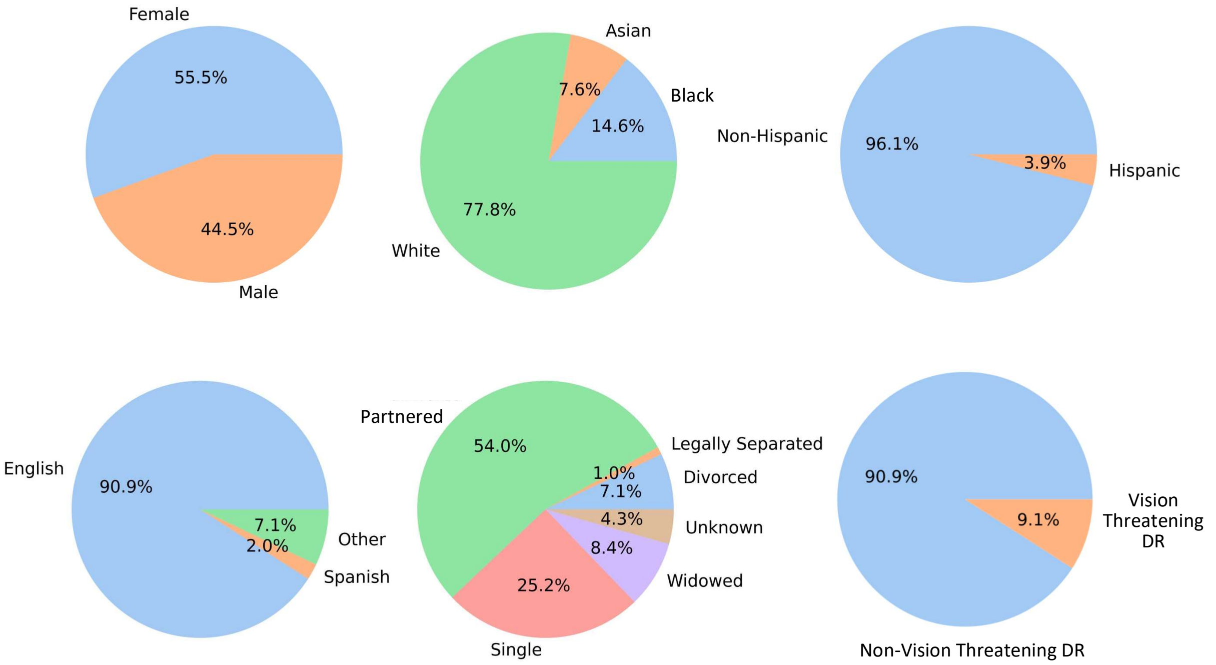

**Figure 3. The demographic distributions of the Harvard-FairVision30k dataset for gender, race, ethnicity, preferred language, marital status, and legally separated status.**

#### Inhouse Color Fundus: Race

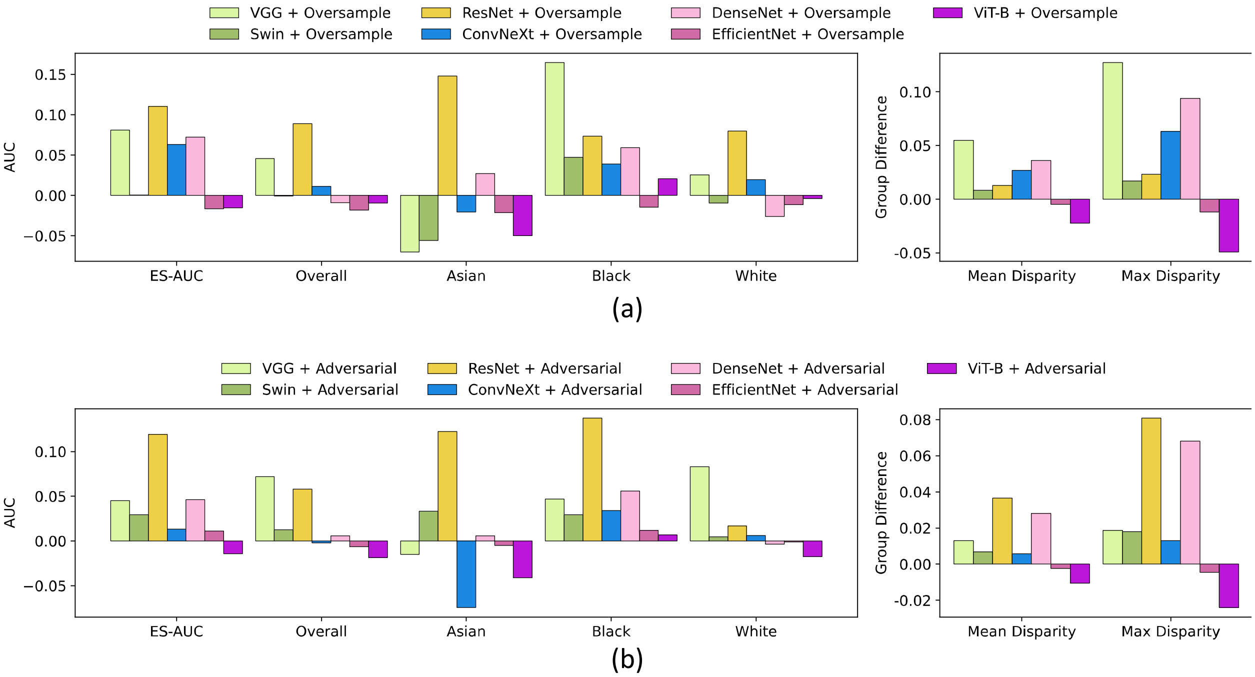


**Figure 4. Results on color fundus images on race attributes of the in-house dataset.** (a) Differences in accuracy for various baseline models after applying oversampling techniques. (b) Differences in accuracy for various baseline models after applying adversarial techniques.

#### Inhouse Color Fundus: Gender

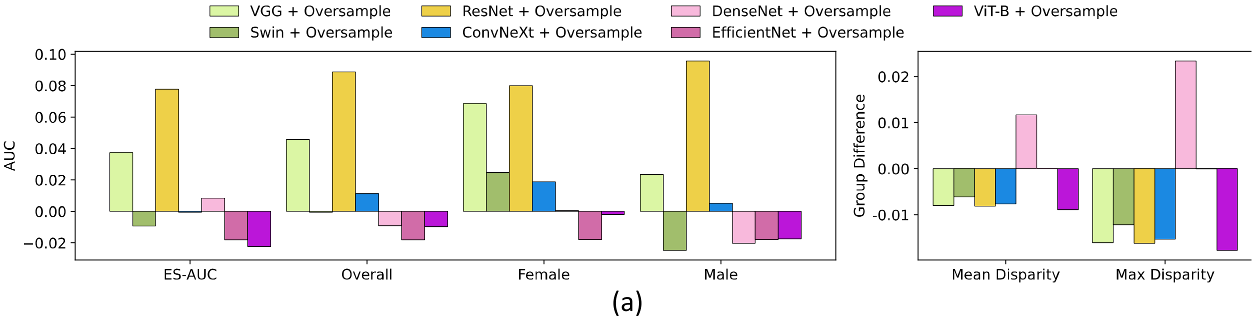

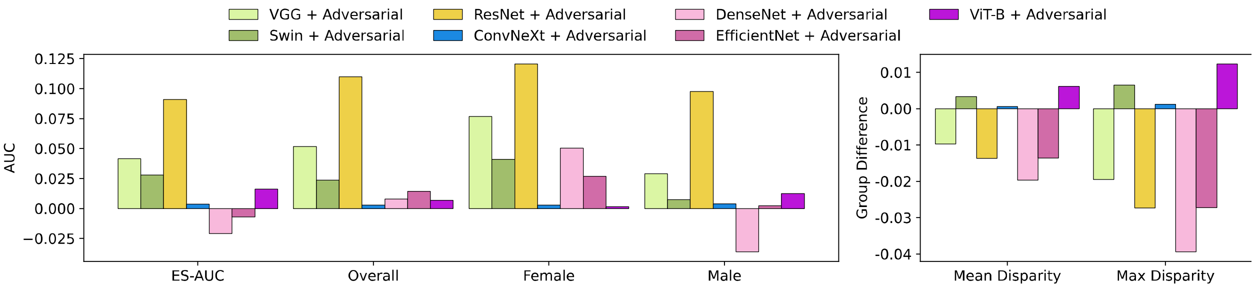


(b)

**Figure 5. Results on color fundus images on gender attributes of the in-house dataset.** (a) Differences in accuracy for various baseline models after applying oversampling techniques. (b) Differences in accuracy for various baseline models after applying adversarial techniques.

#### Inhouse Color Fundus: Ethnicity

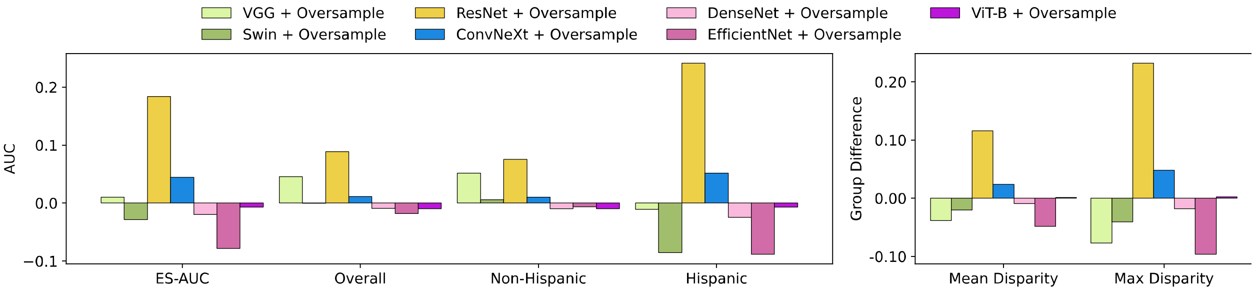


(a)

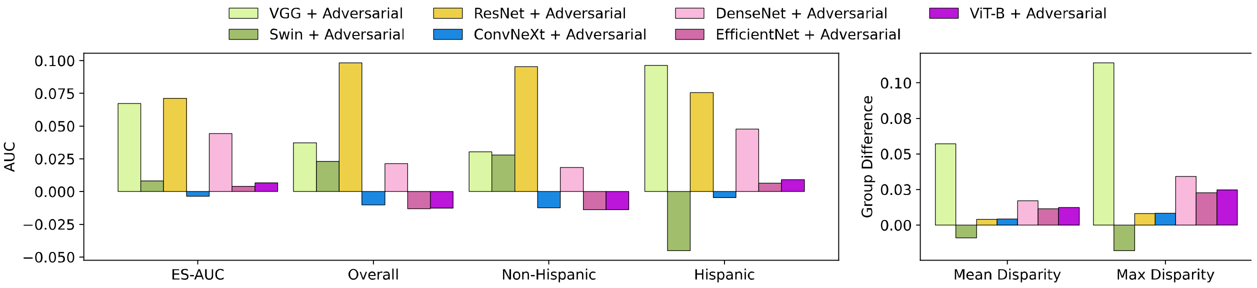


(b)

**Figure 6. Results on color fundus images on ethnicity attributes of the in-house dataset.** (a) Differences in accuracy for various baseline models after applying oversampling techniques. (b) Differences in accuracy for various baseline models after applying adversarial techniques.

#### Inhouse Color Fundus: Language

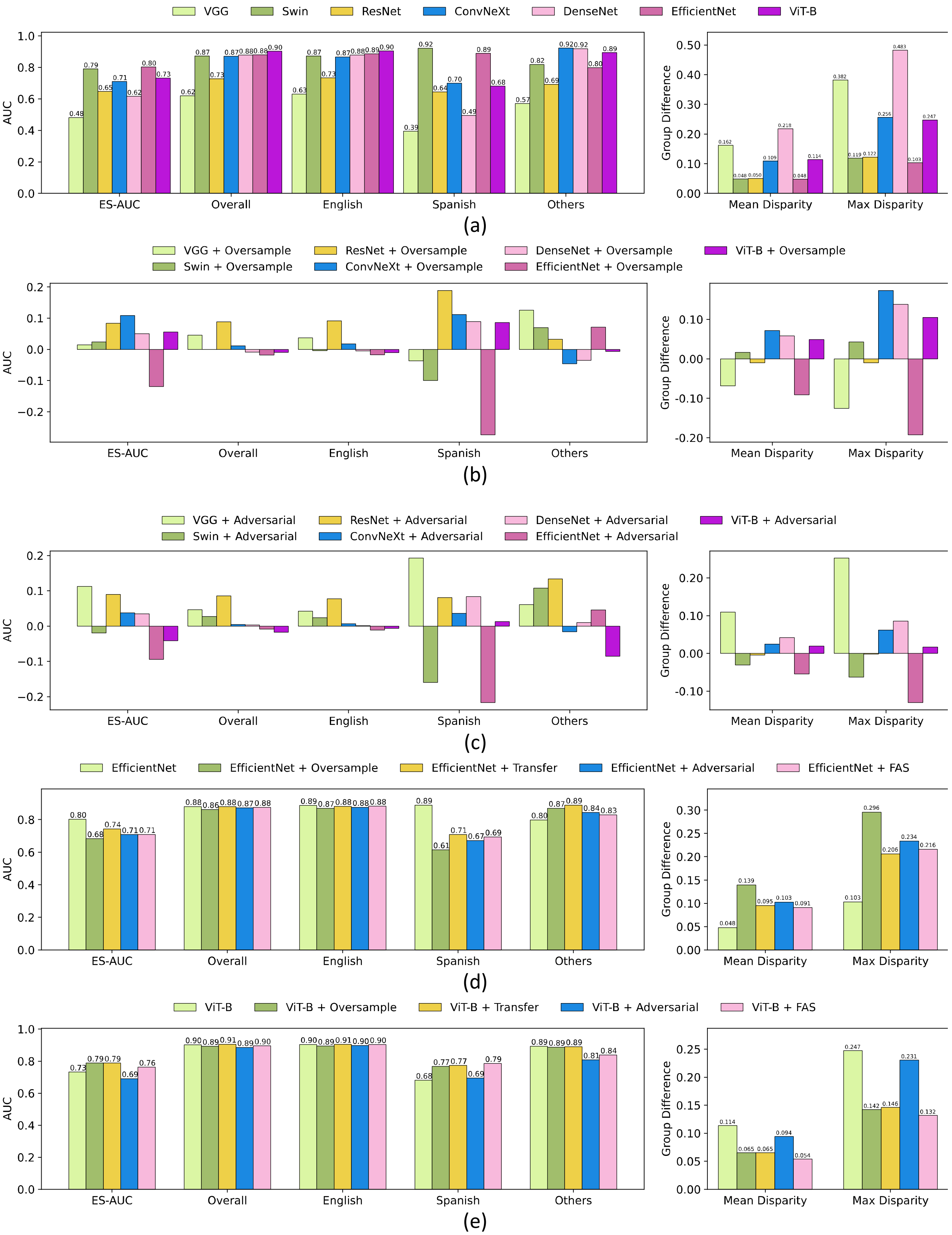
VGG Swin ResNet ConvNeXt DenseNet EfficientNet ViT-B
AUC
Group Difference
ES-AUC Overall English Spanish Others
Mean Disparity Max Disparity
0.48 0.79 0.65 0.71 0.62 0.80 0.73
0.62 0.87 0.73 0.87 0.88 0.88 0.90
0.63 0.87 0.73 0.87 0.88 0.89 0.90
0.39 0.92 0.64 0.70 0.49 0.89 0.68
0.57 0.82 0.69 0.92 0.92 0.80 0.89
0.162 0.048 0.050 0.109 0.218 0.048 0.114
0.382 0.119 0.122 0.256 0.483 0.103 0.247
(a)
VGG + Oversample Swin + Oversample ResNet + Oversample ConvNeXt + Oversample DenseNet + Oversample EfficientNet + Oversample ViT-B + Oversample
(b)
VGG + Adversarial Swin + Adversarial ResNet + Adversarial ConvNeXt + Adversarial DenseNet + Adversarial EfficientNet + Adversarial ViT-B + Adversarial
(c)
EfficientNet EfficientNet + Oversample EfficientNet + Transfer EfficientNet + Adversarial EfficientNet + FAS
0.80 0.68 0.74 0.71 0.71
0.88 0.86 0.88 0.87 0.88
0.89 0.87 0.88 0.88 0.88
0.89 0.61 0.71 0.67 0.69
0.80 0.87 0.89 0.84 0.83
0.048 0.139 0.095 0.103 0.091
0.103 0.296 0.206 0.234 0.216
(d)
ViT-B ViT-B + Oversample ViT-B + Transfer ViT-B + Adversarial ViT-B + FAS
0.73 0.79 0.79 0.69 0.76
0.90 0.89 0.91 0.89 0.90
0.90 0.89 0.91 0.90 0.90
0.68 0.77 0.77 0.69 0.79
0.89 0.89 0.89 0.81 0.84
0.114 0.065 0.065 0.094 0.054
0.247 0.142 0.146 0.231 0.132
(e)

**Figure 7. Results on color fundus images on language attributes of the in-house dataset.** (a) The accuracy of various baseline models. (b) Differences in accuracy for various baseline models after applying oversampling techniques. (c) Differences in accuracy for various baseline models after applying adversarial techniques. (d) The accuracy of EfficientNet and its integration with oversampling, adversarial, transfer learning and our FAS techniques. (e) The accuracy of ViT-B and its integration with oversampling, adversarial, transfer learning and our FAS techniques.

**Inhouse Color Fundus: Marital Status**

| | ES-AUC | Overall AUC | Married & Partnered | Single | Divorced | Widowed | Legally Separated | Mean Disparity | Max Disparity |
|---|---|---|---|---|---|---|---|---|---|
| VGG | 0.521 | 0.620 | 0.631 | 0.615 | 0.591 | 0.516 | 0.661 | 0.079 | 0.234 |
| Swin | 0.769 | 0.907 | 0.904 | 0.906 | 0.941 | 0.859 | 1.000 | 0.052 | 0.156 |
| ResNet | 0.667 | 0.728 | 0.732 | 0.721 | 0.723 | 0.730 | 0.800 | 0.041 | 0.109 |
| ConvNeXt | 0.667 | 0.871 | 0.890 | 0.880 | 0.771 | 0.822 | 1.000 | 0.088 | 0.263 |
| DenseNet | 0.694 | 0.878 | 0.877 | 0.892 | 0.920 | 0.792 | 1.000 | 0.077 | 0.237 |
| EfficientNet | 0.724 | 0.897 | 0.898 | 0.924 | 0.866 | 0.828 | 0.786 | 0.055 | 0.155 |
| ViT-B | 0.756 | 0.903 | 0.906 | 0.924 | 0.872 | 0.791 | 0.929 | 0.056 | 0.152 |

**Table 1. The accuracy of various baseline models on marital status attributes for in-house color fundus images.**

| | ES-AUC | Overall AUC | Married & Partnered | Single | Divorced | Widowed | Legally Separated | Mean Disparity | Max Disparity |
|---|---|---|---|---|---|---|---|---|---|
| VGG* | -0.008 | 0.046 | 0.059 | 0.017 | 0.164 | 0.079 | -0.075 | -0.016 | -0.019 |
| Swin* | -0.018 | -0.016 | -0.024 | 0.006 | -0.036 | -0.034 | -0.037 | 0.001 | 0.000 |
| ResNet* | 0.002 | 0.089 | 0.089 | 0.108 | 0.097 | -0.035 | 0.092 | -0.038 | -0.134 |
| ConvNeXt* | 0.044 | 0.011 | 0.006 | 0.022 | 0.037 | 0.021 | -0.024 | 0.023 | 0.072 |
| DenseNet* | 0.101 | -0.009 | -0.014 | -0.037 | -0.008 | 0.100 | -0.136 | 0.052 | 0.171 |
| EfficientNet* | -0.005 | -0.035 | -0.036 | -0.030 | -0.029 | -0.073 | 0.042 | 0.002 | -0.006 |
| ViT-B* | -0.023 | -0.01 | -0.022 | -0.001 | 0.001 | 0.049 | 0.071 | -0.005 | -0.027 |

**Table 2. Differences in accuracy of various baseline models after applying oversampling techniques on marital status attributes using in-house color fundus images.** * denotes a model with oversampling techniques.

| | ES-AUC | Overall AUC | Married & Partnered | Single | Divorced | Widowed | Legally Separated | Mean Disparity | Max Disparity |
|---|---|---|---|---|---|---|---|---|---|
| VGG# | -0.022 | 0.063 | 0.033 | 0.085 | 0.153 | 0.048 | 0.172 | -0.051 | -0.160 |
| Swin# | -0.006 | -0.001 | -0.013 | 0.024 | -0.040 | -0.002 | 0.000 | -0.001 | -0.003 |
| ResNet# | 0.059 | 0.128 | 0.117 | 0.143 | 0.171 | 0.065 | 0.122 | -0.009 | -0.040 |
| ConvNeXt# | -0.117 | -0.117 | -0.101 | -0.139 | -0.089 | -0.201 | -0.126 | -0.027 | -0.072 |
| DenseNet# | 0.012 | -0.014 | -0.024 | -0.008 | -0.012 | -0.016 | -0.075 | 0.016 | 0.065 |
| EfficientNet# | 0.033 | -0.025 | -0.023 | -0.039 | 0.009 | -0.040 | 0.137 | 0.004 | 0.000 |

| ViT-B# | -0.034 | -0.004 | 0.004 | -0.030 | 0.042 | -0.039 | 0.037 | -0.023 | -0.085 |
|---|---|---|---|---|---|---|---|---|---|

**Table 3. Differences in accuracy of various baseline models after applying adversarial techniques on marital status attributes using in-house color fundus images.** # denotes a model with adversarial techniques.

| | ES-AUC | Overall AUC | Married & Partnered | Single | Divorced | Widowed | Legally Separated | Mean Disparity | Max Disparity |
|---|---|---|---|---|---|---|---|---|---|
| EfficientNet | 0.724 | 0.897 | 0.898 | 0.924 | 0.866 | 0.828 | 0.786 | 0.055 | 0.155 |
| EfficientNet* | 0.719 | 0.862 | 0.862 | 0.894 | 0.836 | 0.756 | 0.828 | 0.053 | 0.160 |
| EfficientNet¶ | 0.700 | 0.879 | 0.883 | 0.898 | 0.917 | 0.746 | 0.941 | 0.078 | 0.222 |
| EfficientNet# | 0.757 | 0.872 | 0.874 | 0.885 | 0.876 | 0.788 | 0.923 | 0.051 | 0.155 |
| EfficientNet† | 0.728 | 0.902 | 0.898 | 0.922 | 0.920 | 0.803 | 1.000 | 0.070 | 0.219 |

**Table 4. The performances of EfficientNet and its variants on marital status attributes using in-house color fundus images.** * denotes EfficientNet with oversampling, ¶ denotes EfficientNet with transfer learning, # denotes EfficientNet with adversarial, and † denotes EfficientNet with our FAS techniques.

| | ES-AUC | Overall AUC | Married & Partnered | Single | Divorced | Widowed | Legally Separated | Mean Disparity | Max Disparity |
|---|---|---|---|---|---|---|---|---|---|
| ViT-B | 0.756 | 0.903 | 0.906 | 0.924 | 0.872 | 0.791 | 0.929 | 0.056 | 0.152 |
| ViT-B* | 0.733 | 0.893 | 0.884 | 0.923 | 0.873 | 0.840 | 1.000 | 0.061 | 0.179 |
| ViT-B¶ | 0.729 | 0.908 | 0.909 | 0.927 | 0.910 | 0.764 | 0.988 | 0.081 | 0.247 |
| ViT-B# | 0.722 | 0.898 | 0.910 | 0.895 | 0.913 | 0.752 | 0.966 | 0.080 | 0.237 |
| ViT-B† | 0.760 | 0.916 | 0.936 | 0.921 | 0.873 | 0.808 | 0.944 | 0.055 | 0.149 |

**Table 5. The performances of ViT-B and its variants on marital status attributes using in-house color fundus images.** * denotes ViT-B with oversampling, ¶ denotes EfficientNet with transfer learning, # denotes EfficientNet with adversarial, and † denotes ViT-B with our FAS techniques.

##### Inhouse Color Fundus: Gender + Race

| | ES-AUC | Overall AUC | Female Asian | Female Black | Female White | Male Asian | Male Black | Male White | Mean Disparity | Max Disparity |
|---|---|---|---|---|---|---|---|---|---|---|
| VGG | 0.422 | 0.620 | 0.763 | 0.549 | 0.458 | 0.553 | 0.621 | 0.645 | 0.792 | 2.546 |
| Swin | 0.732 | 0.907 | 0.945 | 0.902 | 0.897 | 0.783 | 0.938 | 0.875 | 0.132 | 0.399 |
| ResNet | 0.547 | 0.728 | 0.588 | 0.700 | 0.705 | 0.629 | 0.742 | 0.755 | 0.261 | 0.731 |
| ConvNeXt | 0.713 | 0.871 | 0.922 | 0.882 | 0.904 | 0.766 | 0.85 | 0.87 | 0.135 | 0.420 |
| DenseNet | 0.611 | 0.878 | 0.746 | 0.922 | 0.784 | 0.751 | 0.886 | 0.911 | 0.197 | 0.465 |
| EfficientNet | 0.692 | 0.897 | 0.980 | 0.894 | 0.873 | 0.737 | 0.893 | 0.918 | 0.185 | 0.614 |
| ViT-B | 0.768 | 0.903 | 0.888 | 0.846 | 0.908 | 0.827 | 0.91 | 0.887 | 0.077 | 0.207 |

**Table 6. The accuracy of various baseline models on gender and race attributes for in-house color fundus images.**

| | ES-AUC | Overall AUC | Female Asian | Female Black | Female White | Male Asian | Male Black | Male White | Mean Disparity | Max Disparity |
|---|---|---|---|---|---|---|---|---|---|---|
| VGG* | 0.127 | 0.046 | -0.221 | 0.11 | 0.235 | 0.102 | 0.059 | -0.008 | 0.494 | 1.631 |
| Swin* | -0.024 | -0.016 | -0.128 | 0.021 | 0.008 | 0.002 | -0.027 | 0.006 | 0.000 | 0.043 |
| ResNet* | 0.078 | 0.089 | 0.168 | 0.177 | 0.097 | 0.049 | 0.070 | 0.090 | 0.058 | 0.101 |
| ConvNeXt* | 0.062 | 0.011 | -0.060 | 0.022 | 0.017 | 0.073 | 0.037 | 0.002 | 0.065 | 0.206 |
| DenseNet* | 0.152 | -0.009 | 0.147 | -0.078 | 0.017 | 0.104 | -0.010 | -0.044 | 0.118 | 0.214 |
| EfficientNet* | 0.067 | -0.035 | -0.204 | -0.030 | -0.020 | 0.095 | -0.024 | -0.057 | 0.097 | 0.357 |
| ViT-B* | -0.067 | -0.010 | -0.001 | -0.088 | 0.036 | -0.001 | -0.007 | 0.001 | -0.076 | -0.269 |

**Table 7. Differences in accuracy of various baseline models after applying oversampling techniques on gender and race attributes using in-house color fundus images.** * denotes a model with oversampling techniques.

| | ES-AUC | Overall AUC | Female Asian | Female Black | Female White | Male Asian | Male Black | Male White | Mean Disparity | Max Disparity |
|---|---|---|---|---|---|---|---|---|---|---|
| VGG# | 0.063 | 0.079 | -0.116 | 0.113 | 0.070 | 0.031 | 0.134 | 0.041 | 0.427 | 1.402 |
| Swin# | 0.025 | -0.001 | -0.016 | 0.075 | 0.020 | 0.051 | -0.048 | 0.033 | 0.026 | 0.045 |
| ResNet# | 0.001 | -0.066 | 0.024 | -0.004 | -0.041 | -0.039 | -0.050 | -0.112 | 0.019 | 0.073 |
| ConvNeXt# | 0.109 | 0.016 | -0.021 | -0.021 | -0.014 | 0.085 | 0.037 | 0.017 | 0.090 | 0.290 |
| DenseNet# | 0.104 | -0.003 | -0.020 | -0.084 | 0.090 | 0.094 | -0.015 | -0.030 | 0.056 | 0.052 |
| EfficientNet# | -0.004 | -0.007 | -0.124 | -0.154 | 0.004 | 0.066 | -0.001 | -0.022 | 0.042 | 0.216 |
| ViT-B# | -0.010 | 0.021 | -0.047 | 0.022 | 0.011 | 0.052 | 0.028 | 0.018 | 0.000 | -0.025 |

**Table 8. Differences in accuracy of various baseline models after applying adversarial techniques on gender and race attributes using in-house color fundus images.** # denotes a model with adversarial techniques.

| | ES-AUC | Overall AUC | Female Asian | Female Black | Female White | Male Asian | Male Black | Male White | Mean Disparity | Max Disparity |
|---|---|---|---|---|---|---|---|---|---|---|
| EfficientNet | 0.692 | 0.897 | 0.980 | 0.894 | 0.873 | 0.737 | 0.893 | 0.918 | 0.185 | 0.614 |
| EfficientNet* | 0.759 | 0.862 | 0.776 | 0.863 | 0.852 | 0.832 | 0.870 | 0.861 | 0.088 | 0.257 |
| EfficientNet# | 0.689 | 0.890 | 0.856 | 0.740 | 0.876 | 0.803 | 0.892 | 0.895 | 0.143 | 0.398 |
| EfficientNet† | 0.674 | 0.893 | 0.974 | 0.898 | 0.831 | 0.734 | 0.906 | 0.898 | 0.191 | 0.611 |

**Table 9. The performances of EfficientNet and its variants on gender and race attributes using in-house color fundus images.** * denotes EfficientNet with oversampling, # denotes EfficientNet with adversarial, and † denotes EfficientNet with our FAS techniques.

| | ES-AUC | Overall AUC | Female Asian | Female Black | Female White | Male Asian | Male Black | Male White | Mean Disparity | Max Disparity |
|---|---|---|---|---|---|---|---|---|---|---|
| ViT-B | 0.768 | 0.903 | 0.888 | 0.846 | 0.908 | 0.827 | 0.910 | 0.887 | 0.077 | 0.207 |
| ViT-B* | 0.700 | 0.893 | 0.887 | 0.758 | 0.945 | 0.826 | 0.903 | 0.888 | 0.153 | 0.476 |
| ViT-B# | 0.757 | 0.923 | 0.841 | 0.869 | 0.919 | 0.879 | 0.939 | 0.905 | 0.077 | 0.231 |

| | | | | | | | | | |
|---|---|---|---|---|---|---|---|---|---|
| ViT-B† | 0.789 | 0.910 | 0.918 | 0.918 | 0.895 | 0.830 | 0.931 | 0.889 | 0.081 | 0.247 |

**Table 10. The performances of ViT-B and its variants on gender and race status attributes using in-house color fundus images.** * denotes ViT-B with oversampling, # denotes EfficientNet with adversarial, and † denotes ViT-B with our FAS techniques.

**Inhouse Color Fundus: Gender + Ethnicity**

| | ES-AUC | Overall AUC | Female Non-Hispanic | Female Hispanic | Female Non-Hispanic | Male Hispanic | Mean Disparity | Max Disparity |
|---|---|---|---|---|---|---|---|---|
| VGG | 0.485 | 0.620 | 0.614 | 0.642 | 0.543 | 0.448 | 0.628 | 1.629 |
| Swin | 0.764 | 0.907 | 0.937 | 0.880 | 0.937 | 0.807 | 0.132 | 0.321 |
| ResNet | 0.537 | 0.728 | 0.733 | 0.750 | 0.647 | 0.482 | 0.467 | 1.179 |
| ConvNeXt | 0.768 | 0.871 | 0.877 | 0.872 | 0.810 | 0.805 | 0.091 | 0.194 |
| DenseNet | 0.717 | 0.878 | 0.864 | 0.905 | 0.925 | 0.742 | 0.188 | 0.485 |
| EfficientNet | 0.701 | 0.897 | 0.913 | 0.889 | 0.667 | 0.922 | 0.265 | 0.643 |
| ViT-B | 0.783 | 0.903 | 0.922 | 0.890 | 0.830 | 0.852 | 0.087 | 0.226 |

**Table 11. The accuracy of various baseline models on gender and ethnicity attributes for in-house color fundus images.**

| | ES-AUC | Overall AUC | Female Non-Hispanic | Female Hispanic | Female Non-Hispanic | Male Hispanic | Mean Disparity | Max Disparity |
|---|---|---|---|---|---|---|---|---|
| VGG* | -0.011 | 0.046 | 0.076 | 0.027 | -0.033 | -0.001 | 0.004 | 0.161 |
| Swin* | -0.026 | -0.016 | -0.025 | 0.004 | -0.113 | -0.030 | -0.003 | -0.024 |
| ResNet* | 0.189 | 0.089 | 0.081 | 0.069 | 0.074 | 0.357 | 0.322 | 0.806 |
| ConvNeXt* | 0.064 | 0.011 | 0.019 | 0.002 | 0.070 | 0.040 | 0.043 | 0.062 |
| DenseNet* | 0.035 | -0.009 | 0.007 | -0.029 | -0.148 | 0.073 | 0.077 | 0.216 |
| EfficientNet* | -0.004 | -0.035 | -0.034 | -0.017 | 0.084 | -0.158 | 0.101 | 0.289 |
| ViT-B* | -0.026 | -0.010 | 0.002 | -0.020 | -0.050 | 0.055 | -0.055 | -0.138 |

**Table 12. Differences in accuracy of various baseline models after applying oversampling techniques on gender and ethnicity attributes using in-house color fundus images.** * denotes a model with oversampling techniques.

| | ES-AUC | Overall AUC | Female Non-Hispanic | Female Hispanic | Female Non-Hispanic | Male Hispanic | Mean Disparity | Max Disparity |
|---|---|---|---|---|---|---|---|---|
| VGG# | 0.099 | 0.087 | 0.108 | 0.059 | 0.136 | 0.098 | 0.294 | 0.774 |
| Swin# | -0.016 | -0.008 | -0.028 | 0.025 | -0.120 | -0.012 | 0.004 | 0.036 |
| ResNet# | 0.080 | 0.098 | 0.114 | 0.078 | 0.064 | 0.144 | 0.191 | 0.502 |
| ConvNeXt# | -0.284 | -0.214 | -0.196 | -0.223 | -0.447 | -0.117 | -0.766 | -1.870 |
| DenseNet# | 0.035 | 0.015 | 0.038 | -0.009 | -0.061 | 0.004 | 0.028 | 0.088 |
| EfficientNet# | 0.033 | -0.015 | -0.010 | -0.017 | 0.131 | -0.126 | 0.142 | 0.360 |
| ViT-B# | 0.051 | 0.003 | -0.003 | 0.005 | 0.041 | 0.024 | 0.041 | 0.111 |

**Table 13. Differences in accuracy of various baseline models after applying adversarial techniques on gender and ethnicity attributes using in-house color fundus images.** # denotes a model with adversarial techniques.

| | ES-AUC | Overall AUC | Female Non-Hispanic | Female Hispanic | Female Non-Hispanic | Male Hispanic | Mean Disparity | Max Disparity |
|---|---|---|---|---|---|---|---|---|
| EfficientNet | 0.701 | 0.897 | 0.913 | 0.889 | 0.667 | 0.922 | 0.265 | 0.643 |
| EfficientNet* | 0.697 | 0.862 | 0.879 | 0.872 | 0.751 | 0.763 | 0.164 | 0.353 |
| EfficientNet# | 0.734 | 0.881 | 0.903 | 0.872 | 0.798 | 0.796 | 0.123 | 0.282 |
| EfficientNet† | 0.688 | 0.903 | 0.937 | 0.889 | 0.779 | 0.762 | 0.182 | 0.434 |

**Table 14. The performances of EfficientNet and its variants on gender and ethnicity attributes using in-house color fundus images.** * denotes EfficientNet with oversampling, # denotes EfficientNet with adversarial, and † denotes EfficientNet with our FAS techniques.

| | ES-AUC | Overall AUC | Female Non-Hispanic | Female Hispanic | Female Non-Hispanic | Male Hispanic | Mean Disparity | Max Disparity |
|---|---|---|---|---|---|---|---|---|
| ViT-B | 0.783 | 0.903 | 0.922 | 0.890 | 0.830 | 0.852 | 0.087 | 0.226 |
| ViT-B* | 0.756 | 0.893 | 0.923 | 0.870 | 0.780 | 0.908 | 0.141 | 0.364 |
| ViT-B# | 0.834 | 0.906 | 0.919 | 0.896 | 0.872 | 0.876 | 0.046 | 0.115 |
| ViT-B† | 0.845 | 0.906 | 0.921 | 0.884 | 0.904 | 0.938 | 0.050 | 0.134 |

**Table 15. The performances of ViT-B and its variants on gender and ethnicity attributes using in-house color fundus images.** * denotes ViT-B with oversampling, # denotes EfficientNet with adversarial, and † denotes ViT-B with our FAS techniques.

##### ODIR Color Fundus: Gender

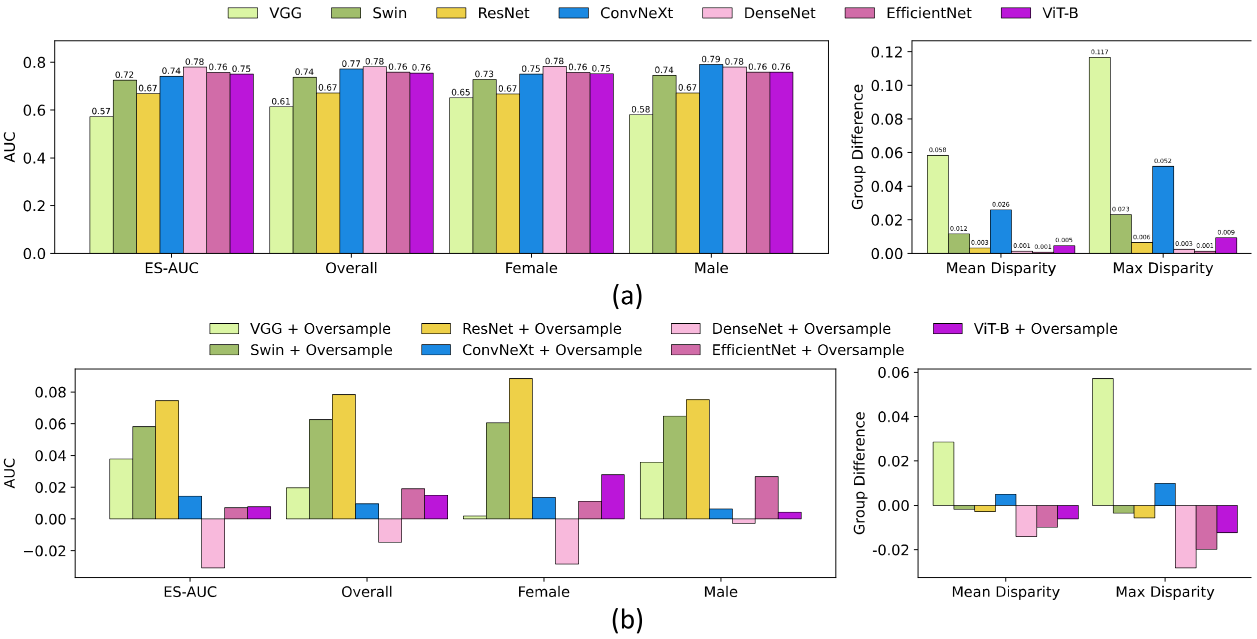

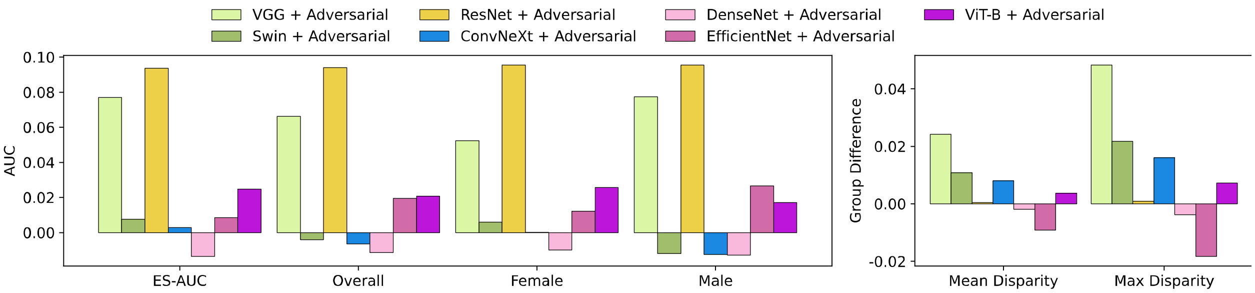


(c)

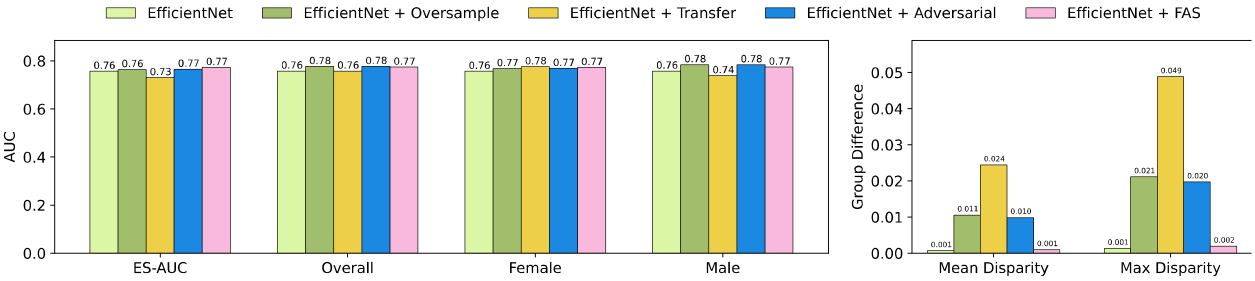


(d)

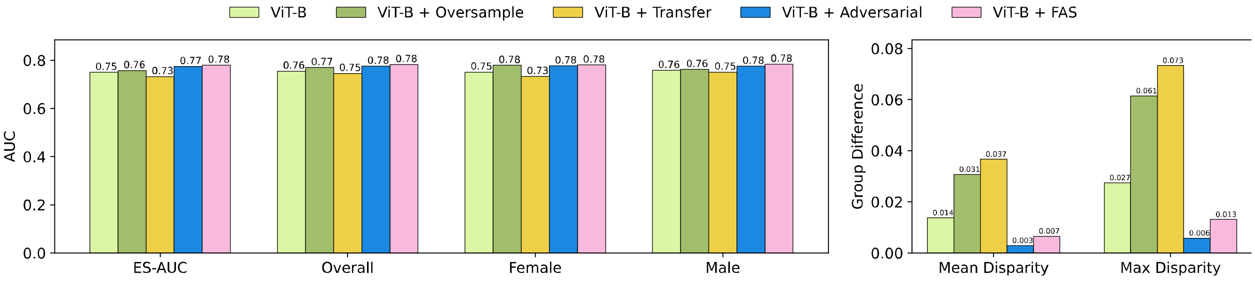


(e)

**Figure 8. Results on color fundus images on gender attributes of the ODIR dataset.** (a) The accuracy of various baseline models. (b) Differences in accuracy for various baseline models after applying oversampling techniques. (c) Differences in accuracy for various baseline models after applying adversarial techniques. (d) The accuracy of EfficientNet and its integration with oversampling, adversarial, transfer learning and our FAS techniques. (e) The accuracy of ViT-B and its integration with oversampling, adversarial, transfer learning and our FAS techniques.

##### Inhouse SLO Fundus: Race

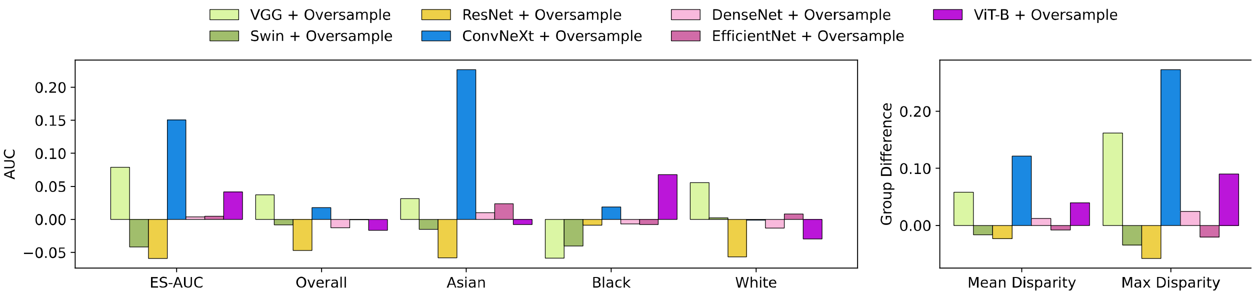


(a)

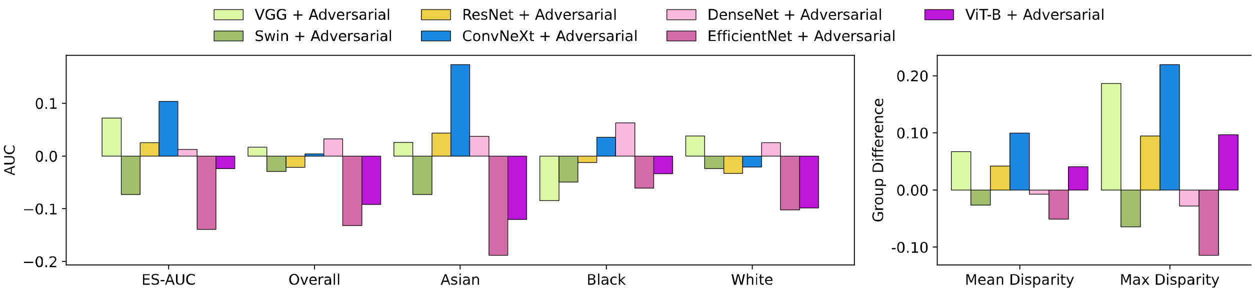


(b)

**Figure 9. Results on SLO fundus images on race attributes of the in-house dataset.** (a) Differences in accuracy for various baseline models after applying oversampling techniques. (b) Differences in accuracy for various baseline models after applying adversarial techniques.

##### Inhouse SLO Fundus: Gender

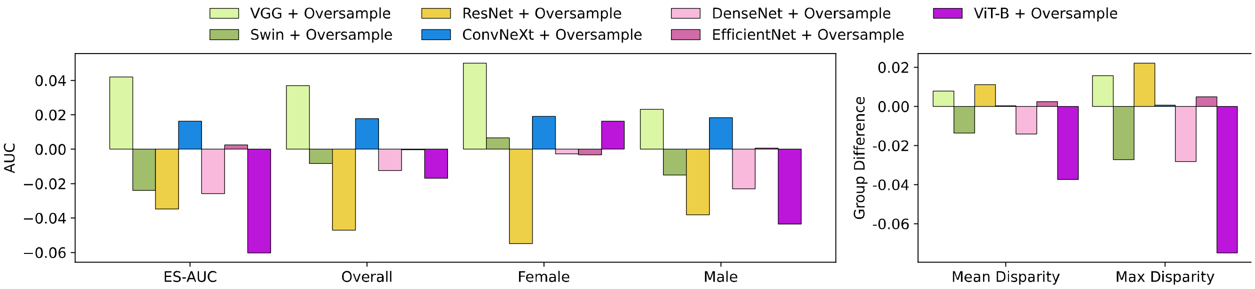


(a)

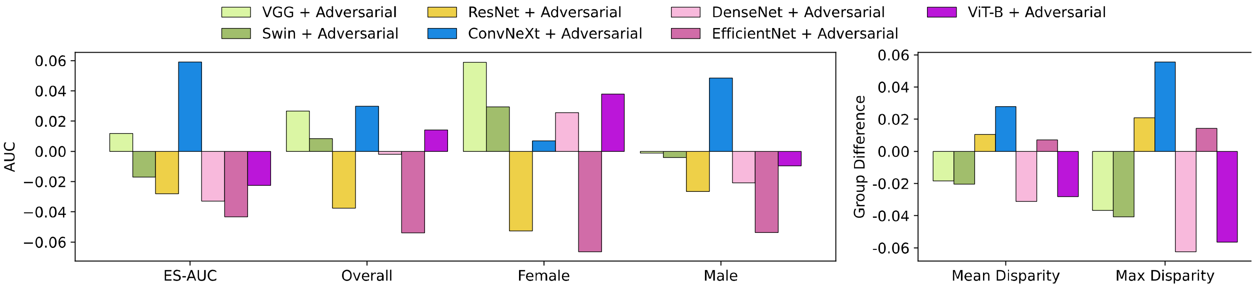


(b)

**Figure 10. Results on SLO fundus images on gender attributes of the in-house dataset.** (a) Differences in accuracy for various baseline models after applying oversampling techniques. (b) Differences in accuracy for various baseline models after applying adversarial techniques.

##### Inhouse SLO Fundus: Ethnicity

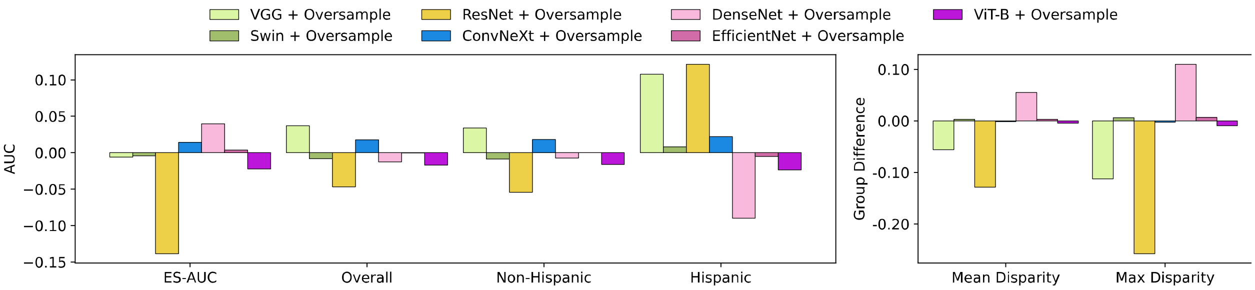


(a)

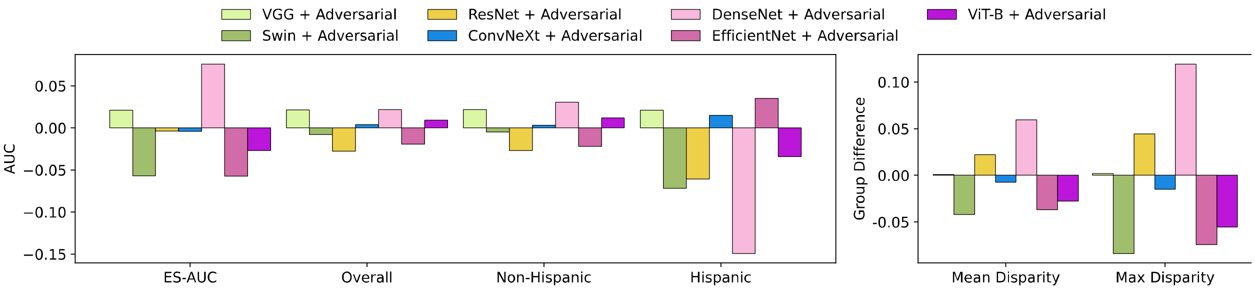


(b)

**Figure 11. Results on SLO fundus images on ethnicity attributes of the in-house dataset.** (a) Differences in accuracy for various baseline models after applying oversampling techniques. (b) Differences in accuracy for various baseline models after applying adversarial techniques.

#### Inhouse SLO Fundus: Language

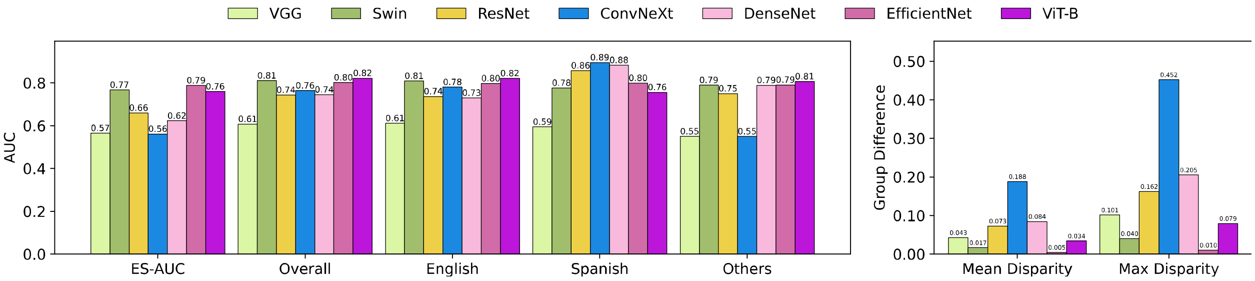


(a)

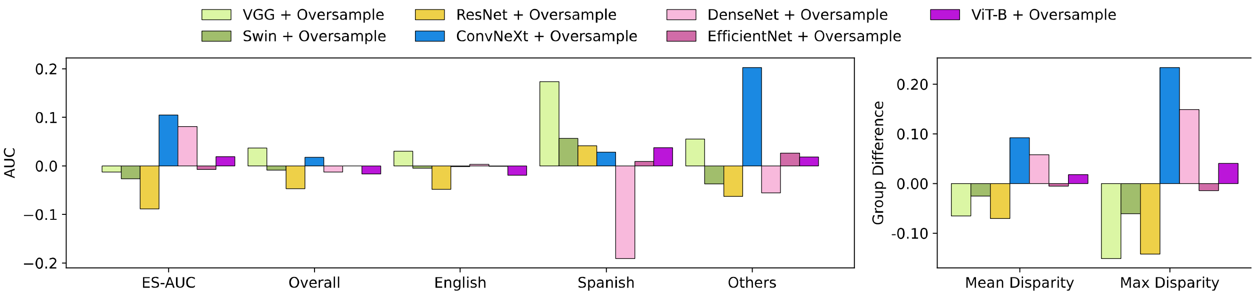


(b)

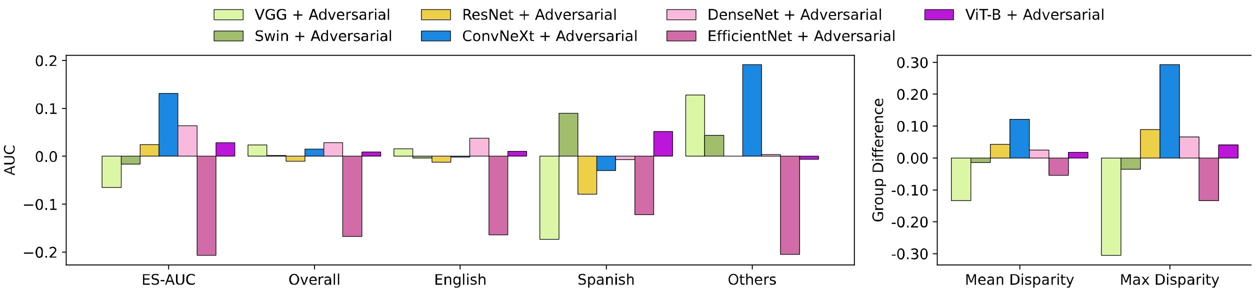


(c)

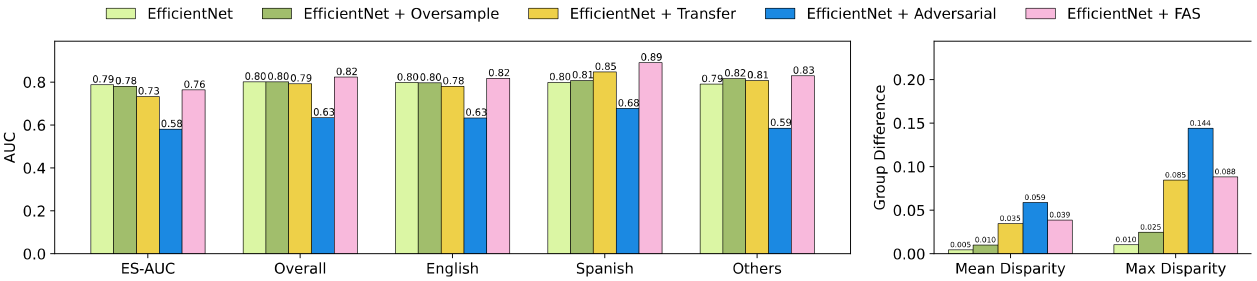


(d)

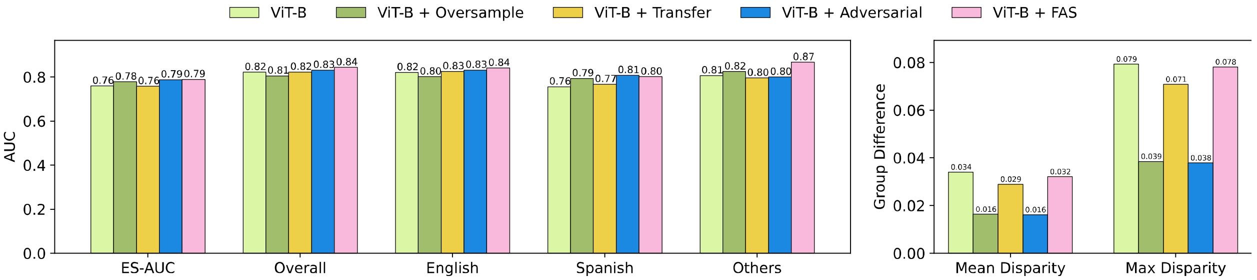


(e)

**Figure 12. Results on SLO fundus images on language attributes of the in-house dataset.** (a) The accuracy of various baseline models. (b) Differences in accuracy for various baseline models after applying oversampling techniques. (c) Differences in accuracy for various baseline models after applying adversarial techniques. (d) The accuracy of EfficientNet and its integration with oversampling, adversarial, transfer learning and our FAS techniques. (e) The accuracy of ViT-B and its integration with oversampling, adversarial, transfer learning and our FAS techniques.

**Inhouse SLO Fundus: Marital Status**

| | ES-AUC | Overall AUC | Married & Partnered | Single | Divorced | Widowed | Legally Separated | Mean Disparity | Max Disparity |
|---|---|---|---|---|---|---|---|---|---|
| VGG | 0.465 | 0.607 | 0.614 | 0.630 | 0.660 | 0.456 | 0.679 | 0.130 | 0.366 |
| Swin | 0.686 | 0.810 | 0.812 | 0.829 | 0.781 | 0.690 | 0.822 | 0.063 | 0.171 |
| ResNet | 0.585 | 0.743 | 0.734 | 0.781 | 0.780 | 0.612 | 0.799 | 0.091 | 0.251 |
| ConvNeXt | 0.654 | 0.764 | 0.753 | 0.780 | 0.798 | 0.718 | 0.824 | 0.048 | 0.140 |
| DenseNet | 0.511 | 0.745 | 0.745 | 0.810 | 0.638 | 0.540 | 0.665 | 0.124 | 0.363 |
| EfficientNet | 0.552 | 0.801 | 0.829 | 0.775 | 0.749 | 0.620 | 0.966 | 0.140 | 0.432 |
| ViT-B | 0.633 | 0.822 | 0.836 | 0.839 | 0.771 | 0.659 | 0.875 | 0.093 | 0.263 |

**Table 16. The accuracy of various baseline models on marital status attributes for in-house SLO fundus images.**

| | ES-AUC | Overall AUC | Married & Partnered | Single | Divorced | Widowed | Legally Separated | Mean Disparity | Max Disparity |
|---|---|---|---|---|---|---|---|---|---|
| VGG* | -0.001 | 0.037 | 0.022 | 0.073 | -0.133 | 0.145 | -0.197 | 0.008 | 0.022 |
| Swin* | -0.070 | -0.008 | -0.006 | 0.000 | 0.024 | -0.023 | 0.112 | -0.043 | -0.163 |
| ResNet* | -0.125 | -0.047 | -0.032 | -0.046 | -0.003 | -0.153 | 0.047 | -0.097 | -0.305 |
| ConvNeXt* | -0.014 | 0.018 | 0.047 | 0.022 | -0.078 | -0.032 | -0.020 | -0.016 | -0.013 |
| DenseNet* | 0.021 | -0.012 | -0.016 | -0.029 | 0.036 | 0.067 | 0.209 | 0.000 | -0.001 |
| EfficientNet* | 0.103 | 0.000 | -0.021 | 0.036 | 0.030 | 0.088 | -0.256 | 0.083 | 0.304 |
| ViT-B* | -0.072 | -0.017 | -0.024 | -0.001 | -0.099 | -0.011 | 0.036 | -0.032 | -0.064 |

**Table 17. Differences in accuracy of various baseline models after applying oversampling techniques on marital status attributes using in-house SLO fundus images.** * denotes a model with oversampling techniques.

| | ES-AUC | Overall AUC | Married & Partnered | Single | Divorced | Widowed | Legally Separated | Mean Disparity | Max Disparity |
|---|---|---|---|---|---|---|---|---|---|
| VGG# | 0.041 | 0.046 | 0.043 | 0.067 | -0.031 | 0.059 | -0.107 | 0.032 | 0.088 |
| Swin# | -0.025 | 0.008 | 0.014 | -0.002 | 0.053 | 0.010 | 0.088 | -0.019 | -0.085 |
| ResNet# | 0.004 | -0.004 | -0.004 | -0.014 | -0.022 | 0.064 | 0.076 | 0.003 | -0.018 |
| ConvNeXt# | -0.007 | 0.021 | 0.054 | -0.017 | -0.084 | 0.095 | 0.027 | -0.012 | -0.035 |
| DenseNet# | -0.013 | -0.001 | 0.009 | -0.032 | 0.044 | -0.026 | 0.237 | -0.047 | -0.158 |
| EfficientNet# | -0.018 | -0.064 | -0.096 | 0.001 | 0.032 | -0.072 | -0.123 | 0.004 | 0.031 |
| ViT-B# | 0.140 | 0.031 | 0.015 | 0.032 | 0.042 | 0.180 | 0.008 | 0.064 | 0.181 |

**Table 18. Differences in accuracy of various baseline models after applying adversarial techniques on marital status attributes using in-house SLO fundus images.** # denotes a model with adversarial techniques.

| | ES-AUC | Overall AUC | Married & Partnered | Single | Divorced | Widowed | Legally Separated | Mean Disparity | Max Disparity |
|---|---|---|---|---|---|---|---|---|---|
| EfficientNet | 0.552 | 0.801 | 0.829 | 0.775 | 0.749 | 0.620 | 0.966 | 0.140 | 0.432 |
| EfficientNet* | 0.655 | 0.801 | 0.809 | 0.811 | 0.779 | 0.708 | 0.710 | 0.057 | 0.128 |
| EfficientNet¶ | 0.603 | 0.796 | 0.800 | 0.780 | 0.712 | 0.665 | 0.881 | 0.094 | 0.271 |
| EfficientNet# | 0.533 | 0.737 | 0.734 | 0.776 | 0.781 | 0.547 | 0.843 | 0.137 | 0.401 |
| EfficientNet† | 0.631 | 0.802 | 0.805 | 0.831 | 0.790 | 0.654 | 0.882 | 0.095 | 0.284 |

**Table 19. The performances of EfficientNet and its variants on marital status attributes using in-house SLO fundus images.** * denotes EfficientNet with oversampling, ¶ denotes EfficientNet with transfer learning, # denotes EfficientNet with adversarial, and † denotes EfficientNet with our FAS techniques.

| | ES-AUC | Overall AUC | Married & Partnered | Single | Divorced | Widowed | Legally Separated | Mean Disparity | Max Disparity |
|---|---|---|---|---|---|---|---|---|---|

| | | | | | | | | | |
|---|---|---|---|---|---|---|---|---|---|
| ViT-B | 0.633 | 0.822 | 0.836 | 0.839 | 0.771 | 0.659 | 0.875 | 0.093 | 0.263 |
| ViT-B* | 0.561 | 0.805 | 0.812 | 0.838 | 0.672 | 0.648 | 0.911 | 0.125 | 0.327 |
| ViT-B¶ | 0.631 | 0.824 | 0.838 | 0.842 | 0.769 | 0.658 | 0.875 | 0.094 | 0.263 |
| ViT-B# | 0.773 | 0.852 | 0.852 | 0.871 | 0.814 | 0.839 | 0.883 | 0.029 | 0.082 |
| ViT-B† | 0.647 | 0.838 | 0.846 | 0.861 | 0.821 | 0.684 | 0.933 | 0.097 | 0.297 |

**Table 20. The performances of ViT-B and its variants on marital status attributes using in-house SLO fundus images.** * denotes ViT-B with oversampling, ¶ denotes EfficientNet with transfer learning, # denotes EfficientNet with adversarial, and † denotes ViT-B with our FAS techniques.

**Inhouse SLO Fundus: Gender + Race**

| | ES-AUC | Overall AUC | Female Asian | Female Black | Female White | Male Asian | Male Black | Male White | Mean Disparity | Max Disparity |
|---|---|---|---|---|---|---|---|---|---|---|
| VGG | 0.378 | 0.607 | 0.379 | 0.503 | 0.764 | 0.678 | 0.563 | 0.604 | 0.202 | 0.634 |
| Swin | 0.683 | 0.810 | 0.753 | 0.773 | 0.819 | 0.759 | 0.825 | 0.795 | 0.035 | 0.089 |
| ResNet | 0.620 | 0.743 | 0.713 | 0.622 | 0.744 | 0.710 | 0.752 | 0.738 | 0.058 | 0.175 |
| ConvNeXt | 0.514 | 0.764 | 0.612 | 0.532 | 0.776 | 0.726 | 0.809 | 0.756 | 0.128 | 0.362 |
| DenseNet | 0.548 | 0.745 | 0.528 | 0.775 | 0.781 | 0.694 | 0.756 | 0.729 | 0.116 | 0.339 |
| EfficientNet | 0.654 | 0.801 | 0.843 | 0.744 | 0.800 | 0.691 | 0.813 | 0.799 | 0.062 | 0.189 |
| ViT-B | 0.629 | 0.822 | 0.856 | 0.867 | 0.743 | 0.698 | 0.846 | 0.822 | 0.077 | 0.206 |

**Table 21. The accuracy of various baseline models on gender and race attributes for in-house SLO fundus images.**

| | ES-AUC | Overall AUC | Female Asian | Female Black | Female White | Male Asian | Male Black | Male White | Mean Disparity | Max Disparity |
|---|---|---|---|---|---|---|---|---|---|---|
| VGG* | 0.158 | 0.037 | 0.267 | -0.021 | -0.115 | -0.017 | 0.066 | 0.039 | 0.107 | 0.357 |
| Swin* | -0.094 | -0.008 | -0.129 | 0.019 | -0.058 | -0.020 | 0.028 | -0.012 | -0.052 | -0.196 |
| ResNet* | -0.041 | -0.047 | -0.017 | -0.059 | -0.018 | 0.011 | -0.071 | -0.043 | -0.020 | -0.057 |
| ConvNeXt* | 0.109 | 0.018 | 0.094 | 0.308 | 0.015 | 0.029 | 0.016 | -0.015 | 0.067 | 0.190 |
| DenseNet* | 0.010 | -0.012 | 0.062 | -0.003 | -0.016 | 0.005 | 0.007 | -0.031 | 0.030 | 0.090 |
| EfficientNet* | -0.047 | 0.000 | -0.125 | 0.055 | 0.043 | -0.057 | -0.006 | 0.021 | -0.027 | -0.070 |
| ViT-B* | -0.008 | -0.017 | 0.066 | -0.035 | 0.082 | 0.060 | 0.005 | -0.057 | 0.008 | 0.000 |

**Table 22. Differences in accuracy of various baseline models after applying oversampling techniques on gender and race attributes for in-house SLO fundus images.** * denotes a model with oversampling techniques.

| | ES-AUC | Overall AUC | Female Asian | Female Black | Female White | Male Asian | Male Black | Male White | Mean Disparity | Max Disparity |
|---|---|---|---|---|---|---|---|---|---|---|
| VGG# | 0.128 | 0.032 | 0.329 | -0.010 | -0.153 | -0.034 | 0.059 | 0.036 | 0.101 | 0.297 |
| Swin# | -0.035 | 0.010 | 0.035 | -0.099 | 0.008 | 0.014 | 0.019 | 0.017 | -0.033 | -0.118 |
| ResNet # | -0.128 | -0.054 | -0.268 | 0.012 | -0.023 | -0.053 | -0.048 | -0.072 | -0.073 | -0.224 |
| ConvNeXt# | 0.087 | 0.029 | 0.136 | 0.090 | 0.003 | 0.028 | 0.024 | 0.024 | 0.047 | 0.097 |
| DenseNet # | 0.088 | 0.024 | 0.338 | -0.027 | 0.031 | 0.096 | 0.009 | 0.015 | 0.061 | 0.180 |
| EfficientNet # | -0.143 | -0.076 | 0.045 | -0.188 | -0.032 | 0.029 | -0.089 | -0.110 | -0.074 | -0.269 |

| ViT-B # | 0.035 | 0.005 | -0.038 | -0.028 | 0.065 | -0.013 | 0.025 | -0.015 | 0.007 | -0.019 |
|---|---|---|---|---|---|---|---|---|---|---|

**Table 23. Differences in accuracy of various baseline models after applying adversarial techniques on gender and race attributes for in-house SLO fundus images.** # denotes a model with adversarial techniques.

| | ES-AUC | Overall AUC | Female Asian | Female Black | Female White | Male Asian | Male Black | Male White | Mean Disparity | Max Disparity |
|---|---|---|---|---|---|---|---|---|---|---|
| EfficientNet | 0.654 | 0.801 | 0.843 | 0.744 | 0.800 | 0.691 | 0.813 | 0.799 | 0.062 | 0.189 |
| EfficientNet* | 0.607 | 0.801 | 0.718 | 0.799 | 0.842 | 0.634 | 0.807 | 0.820 | 0.090 | 0.259 |
| EfficientNet# | 0.512 | 0.725 | 0.888 | 0.555 | 0.768 | 0.721 | 0.724 | 0.689 | 0.136 | 0.459 |
| EfficientNet† | 0.684 | 0.805 | 0.833 | 0.754 | 0.842 | 0.764 | 0.799 | 0.791 | 0.040 | 0.110 |

**Table 24. The performances of EfficientNet and its variants on gender and race attributes for in-house SLO fundus images.** * denotes EfficientNet with oversampling, # denotes EfficientNet with adversarial, and † denotes EfficientNet with our FAS techniques.

| | ES-AUC | Overall AUC | Female Asian | Female Black | Female White | Male Asian | Male Black | Male White | Mean Disparity | Max Disparity |
|---|---|---|---|---|---|---|---|---|---|---|
| ViT-B | 0.629 | 0.822 | 0.856 | 0.867 | 0.743 | 0.698 | 0.846 | 0.822 | 0.077 | 0.206 |
| ViT-B* | 0.621 | 0.805 | 0.923 | 0.832 | 0.825 | 0.757 | 0.851 | 0.765 | 0.069 | 0.205 |
| ViT-B# | 0.664 | 0.827 | 0.819 | 0.839 | 0.807 | 0.685 | 0.871 | 0.808 | 0.070 | 0.225 |
| ViT-B† | 0.597 | 0.819 | 0.706 | 0.898 | 0.802 | 0.726 | 0.861 | 0.792 | 0.083 | 0.235 |

**Table 25. The performances of ViT-B and its variants on gender and race attributes for in-house SLO fundus images.** * denotes ViT-B with oversampling, # denotes EfficientNet with adversarial, and † denotes ViT-B with our FAS techniques.

**Inhouse SLO Fundus: Gender + Ethnicity**

| | ES-AUC | Overall AUC | Female Non-Hispanic | Female Hispanic | Female Non-Hispanic | Male Hispanic | Mean Disparity | Max Disparity |
|---|---|---|---|---|---|---|---|---|
| VGG | 0.516 | 0.607 | 0.598 | 0.609 | 0.569 | 0.734 | 0.104 | 0.272 |
| Swin | 0.714 | 0.810 | 0.825 | 0.796 | 0.831 | 0.725 | 0.052 | 0.130 |
| ResNet | 0.661 | 0.743 | 0.748 | 0.729 | 0.820 | 0.773 | 0.046 | 0.122 |
| ConvNeXt | 0.555 | 0.764 | 0.779 | 0.741 | 0.954 | 0.616 | 0.158 | 0.442 |
| DenseNet | 0.593 | 0.745 | 0.749 | 0.726 | 0.918 | 0.805 | 0.100 | 0.257 |
| EfficientNet | 0.697 | 0.801 | 0.811 | 0.786 | 0.896 | 0.770 | 0.060 | 0.156 |
| ViT-B | 0.722 | 0.822 | 0.831 | 0.816 | 0.860 | 0.737 | 0.055 | 0.149 |

**Table 26. The accuracy of various baseline models on gender and ethnicity attributes for in-house SLO fundus images.**

| | ES-AUC | Overall AUC | Female Non-Hispanic | Female Hispanic | Female Non-Hispanic | Male Hispanic | Mean Disparity | Max Disparity |
|---|---|---|---|---|---|---|---|---|
| VGG* | 0.027 | 0.037 | 0.044 | 0.024 | 0.167 | -0.009 | 0.031 | 0.111 |

| | | | | | | | | |
|---|---|---|---|---|---|---|---|---|
| Swin* | -0.060 | -0.008 | 0.004 | -0.014 | 0.079 | 0.005 | -0.030 | -0.093 |
| ResNet* | -0.186 | -0.047 | -0.063 | -0.045 | 0.113 | 0.129 | -0.122 | -0.235 |
| ConvNeXt* | 0.079 | 0.018 | 0.026 | 0.013 | -0.005 | 0.152 | 0.059 | 0.192 |
| DenseNet* | -0.019 | -0.012 | 0.001 | -0.016 | -0.066 | -0.190 | -0.016 | -0.066 |
| EfficientNet* | -0.109 | 0.000 | -0.006 | 0.004 | 0.048 | -0.175 | -0.095 | -0.278 |
| ViT-B* | -0.136 | -0.017 | 0.018 | -0.042 | 0.021 | -0.154 | -0.088 | -0.221 |

**Table 27. Differences in accuracy of various baseline models after applying oversampling techniques on gender and ethnicity attributes for in-house SLO fundus images.** * denotes a model with oversampling techniques.

| | ES-AUC | Overall AUC | Female Non-Hispanic | Female Hispanic | Female Non-Hispanic | Male Hispanic | Mean Disparity | Max Disparity |
|---|---|---|---|---|---|---|---|---|
| VGG# | 0.049 | 0.040 | 0.043 | 0.037 | -0.001 | -0.027 | 0.028 | 0.058 |
| Swin# | 0.004 | 0.014 | 0.024 | 0.006 | 0.046 | 0.051 | 0.004 | 0.008 |
| ResNet# | -0.098 | -0.034 | -0.018 | -0.044 | 0.015 | -0.153 | -0.065 | -0.182 |
| ConvNeXt# | 0.083 | 0.012 | 0.007 | 0.014 | -0.150 | 0.318 | 0.070 | 0.212 |
| DenseNet# | 0.050 | 0.020 | 0.027 | 0.018 | -0.02 | -0.015 | 0.024 | 0.056 |
| EfficientNet# | -0.217 | -0.106 | -0.139 | -0.067 | -0.425 | 0.103 | -0.147 | -0.424 |
| ViT-B# | -0.117 | 0.025 | 0.047 | 0.007 | 0.095 | -0.125 | -0.095 | -0.256 |

**Table 28. Differences in accuracy of various baseline models after applying adversarial techniques on gender and ethnicity attributes for in-house SLO fundus images.** # denotes a model with adversarial techniques.

| | ES-AUC | Overall AUC | Female Non-Hispanic | Female Hispanic | Female Non-Hispanic | Male Hispanic | Mean Disparity | Max Disparity |
|---|---|---|---|---|---|---|---|---|
| EfficientNet | 0.697 | 0.801 | 0.811 | 0.786 | 0.896 | 0.770 | 0.060 | 0.156 |
| EfficientNet* | 0.588 | 0.801 | 0.805 | 0.791 | 0.943 | 0.595 | 0.155 | 0.435 |
| EfficientNet# | 0.479 | 0.695 | 0.672 | 0.719 | 0.470 | 0.873 | 0.207 | 0.580 |
| EfficientNet† | 0.698 | 0.815 | 0.837 | 0.792 | 0.896 | 0.772 | 0.058 | 0.152 |

**Table 29. The performances of EfficientNet and its variants on gender and ethnicity attributes for in-house SLO fundus images.** * denotes EfficientNet with oversampling, # denotes EfficientNet with adversarial,and † denotes EfficientNet with our FAS techniques.

| | ES-AUC | Overall AUC | Female Non-Hispanic | Female Hispanic | Female Non-Hispanic | Male Hispanic | Mean Disparity | Max Disparity |
|---|---|---|---|---|---|---|---|---|
| ViT-B | 0.722 | 0.822 | 0.831 | 0.816 | 0.860 | 0.737 | 0.055 | 0.149 |
| ViT-B* | 0.587 | 0.805 | 0.848 | 0.774 | 0.881 | 0.583 | 0.144 | 0.370 |
| ViT-B# | 0.606 | 0.847 | 0.878 | 0.823 | 0.955 | 0.612 | 0.150 | 0.405 |
| ViT-B† | 0.632 | 0.827 | 0.840 | 0.820 | 0.883 | 0.593 | 0.136 | 0.350 |

**Table 30. The performances of ViT-B and its variants on gender and ethnicity attributes for in-house SLO fundus images.** * denotes ViT-B with oversampling, # denotes EfficientNet with adversarial, and † denotes ViT-B with our FAS techniques.

#### Harvard-FairVision30k SLO Fundus: Race

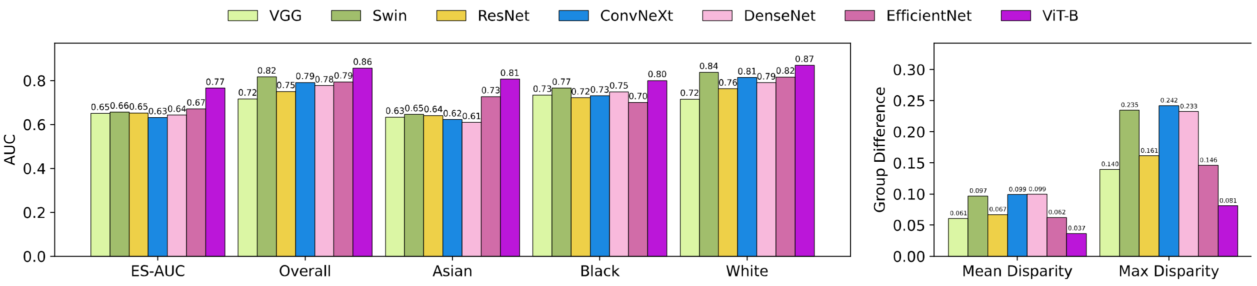


(a)

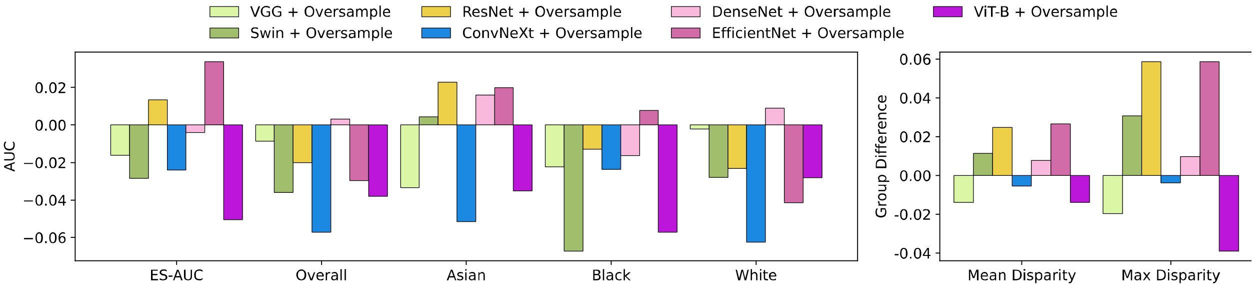


(b)

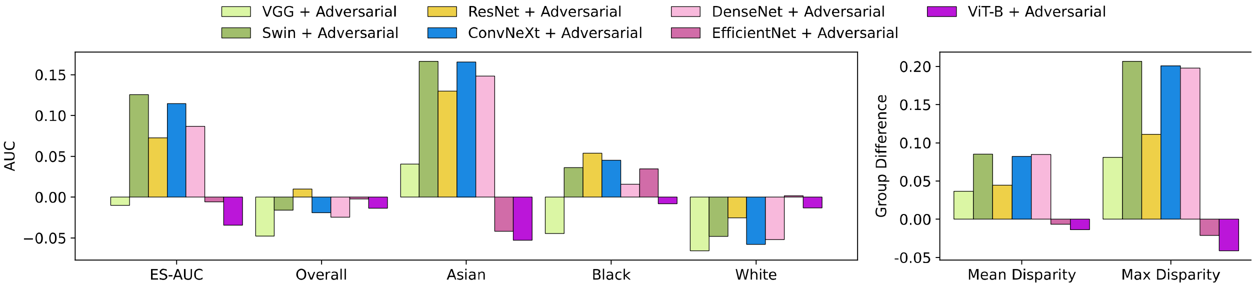


(c)

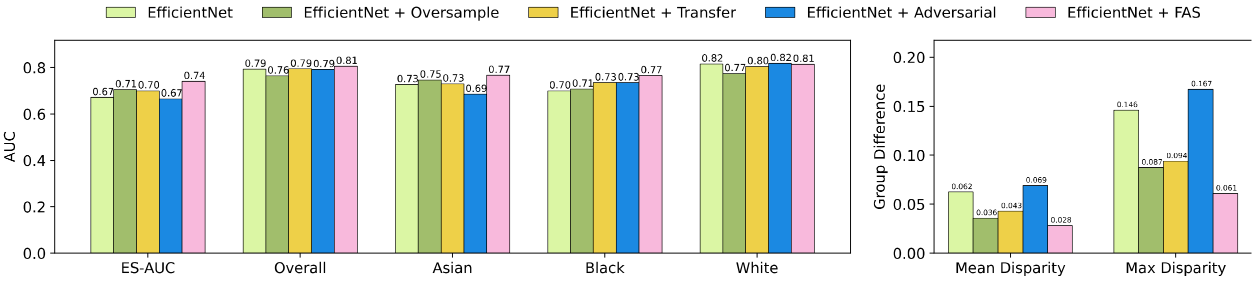


(d)

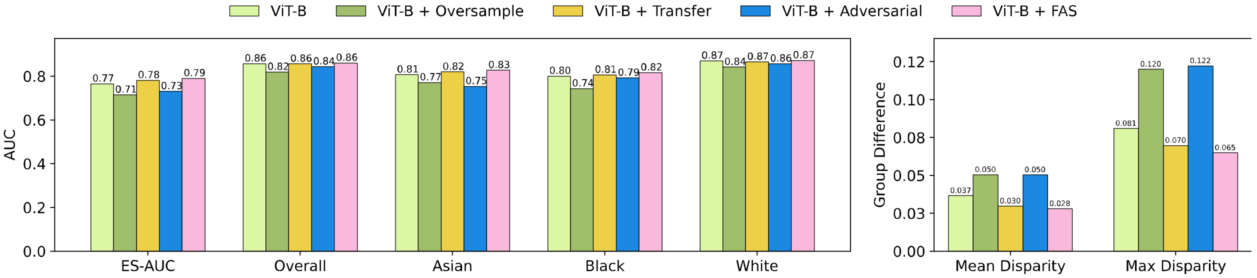


(e)

**Figure 13. Results on SLO fundus images on race attributes of the Harvard-FairVision30k dataset.** (a) The accuracy of various baseline models. (b) Differences in accuracy for various baseline models after applying oversampling techniques. (c) Differences in accuracy for various baseline models after applying adversarial techniques. (d) The accuracy of EfficientNet and its integration with oversampling, adversarial, transfer learning and our FAS techniques. (e) The accuracy of ViT-B and its integration with oversampling, adversarial, transfer learning and our FAS techniques.

#### Harvard-FairVision30k SLO Fundus: Gender

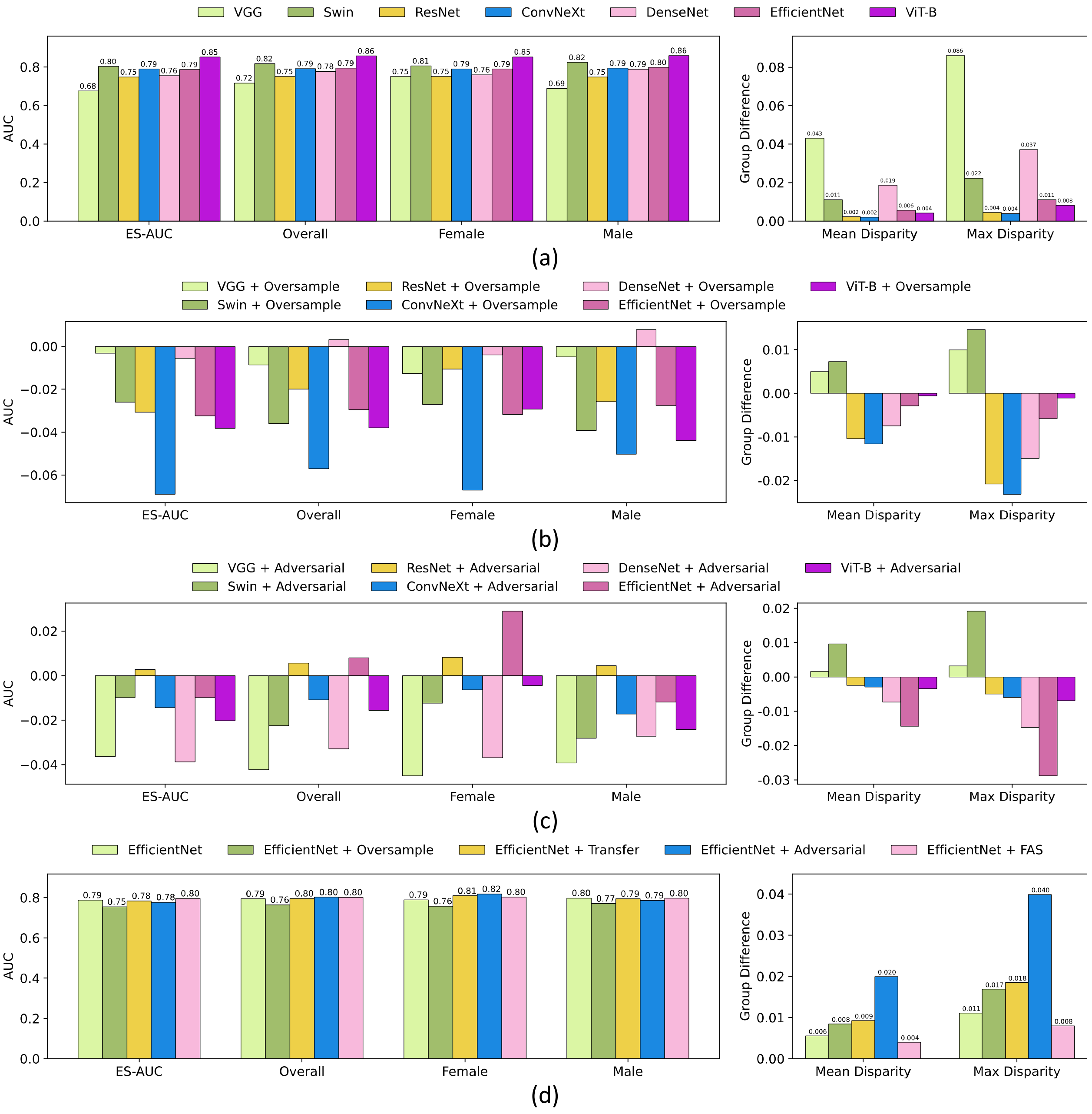

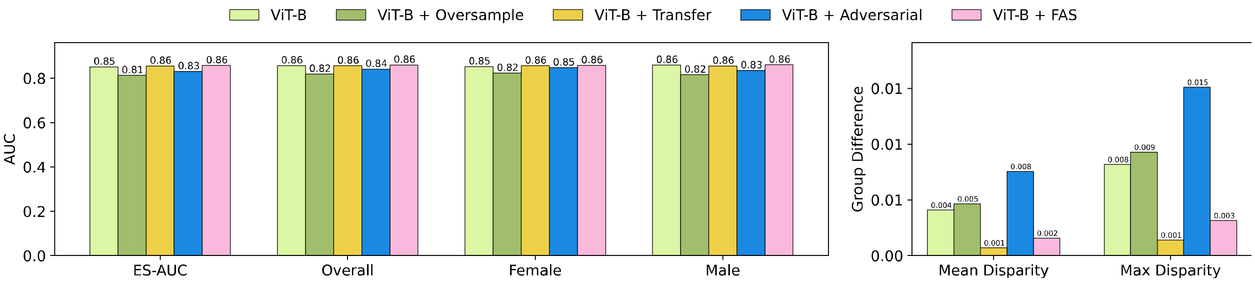


(e)

**Figure 14. Results on SLO fundus images on gender attributes of the Harvard-FairVision30k dataset.** (a) The accuracy of various baseline models. (b) Differences in accuracy for various baseline models after applying oversampling techniques. (c) Differences in accuracy for various baseline models after applying adversarial techniques. (d) The accuracy of EfficientNet and its integration with oversampling, adversarial, transfer learning and our FAS techniques. (e) The accuracy of ViT-B and its integration with oversampling, adversarial, transfer learning and our FAS techniques.

#### Harvard-FairVision30k SLO Fundus: Ethnicity

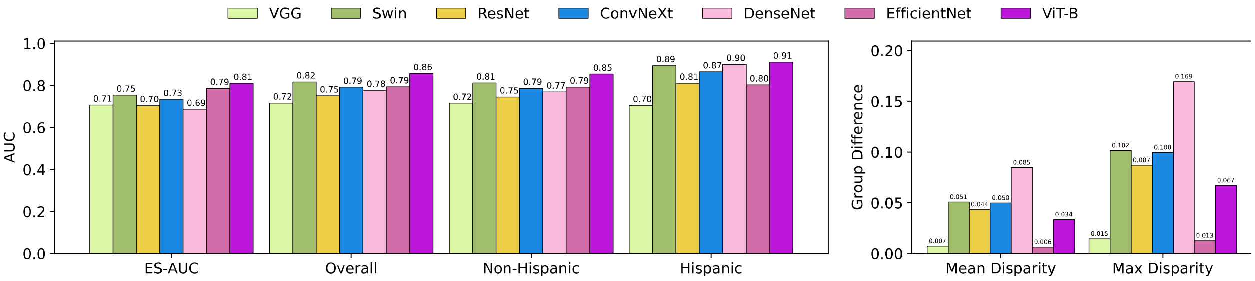


(a)

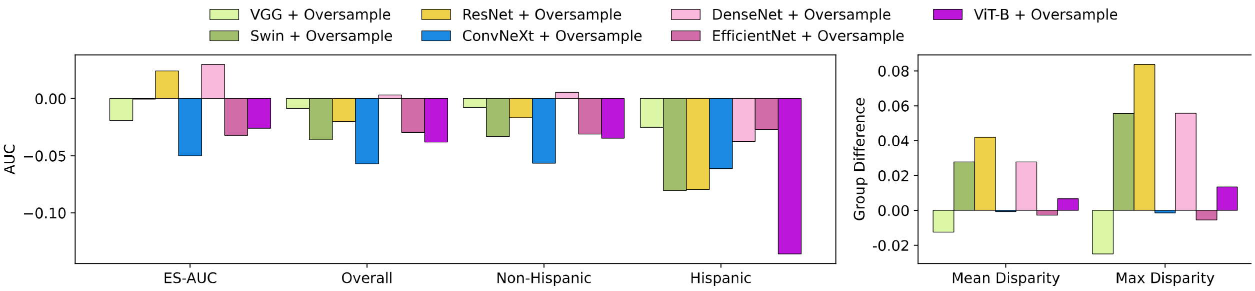


(b)

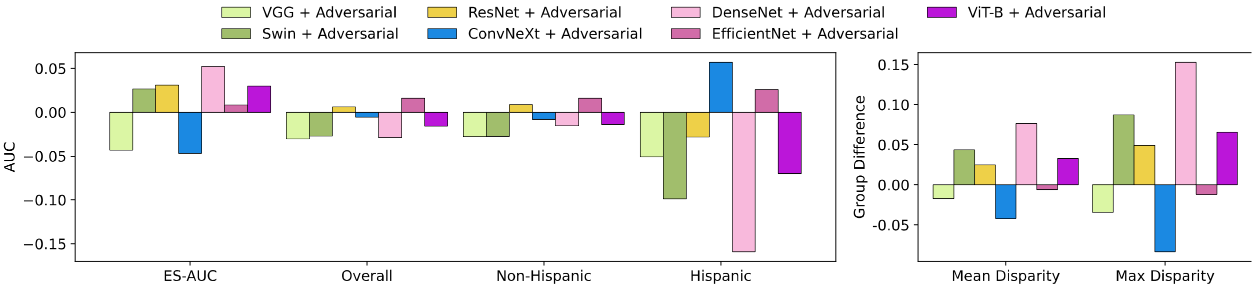

(c)

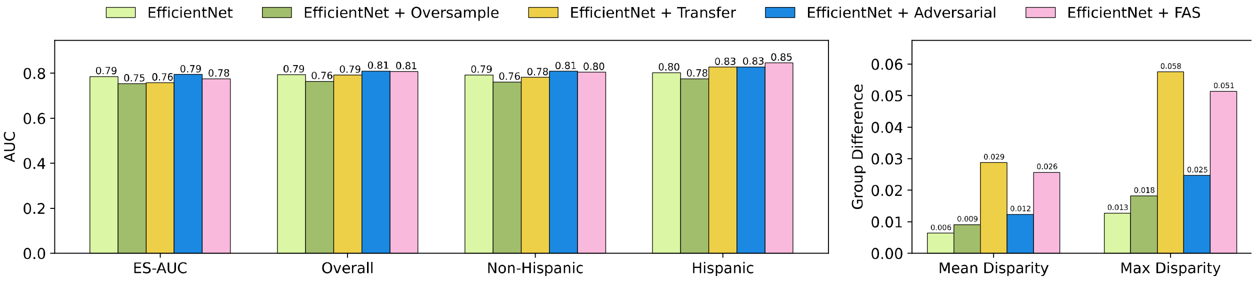


(d)

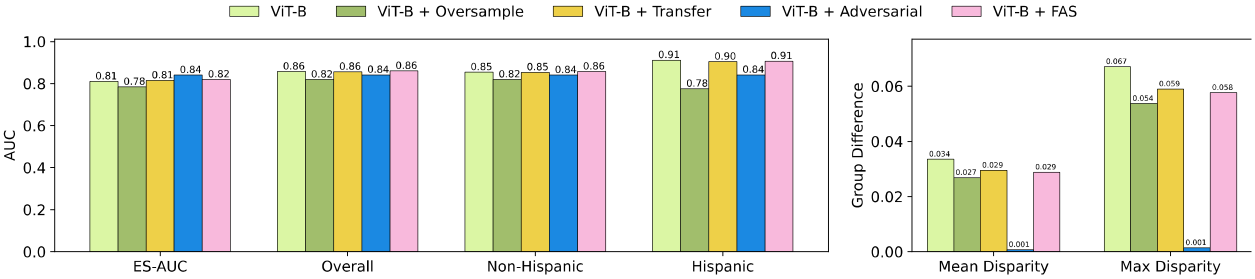


(e)

**Figure 15. Results on SLO fundus images on ethnicity attributes of the Harvard-FairVision30k dataset.** (a) The accuracy of various baseline models. (b) Differences in accuracy for various baseline models after applying oversampling techniques. (c) Differences in accuracy for various baseline models after applying adversarial techniques. (d) The accuracy of EfficientNet and its integration with oversampling, adversarial, transfer learning and our FAS techniques. (e) The accuracy of ViT-B and its integration with oversampling, adversarial, transfer learning and our FAS techniques.

#### Harvard-FairVision30k SLO Fundus: Language

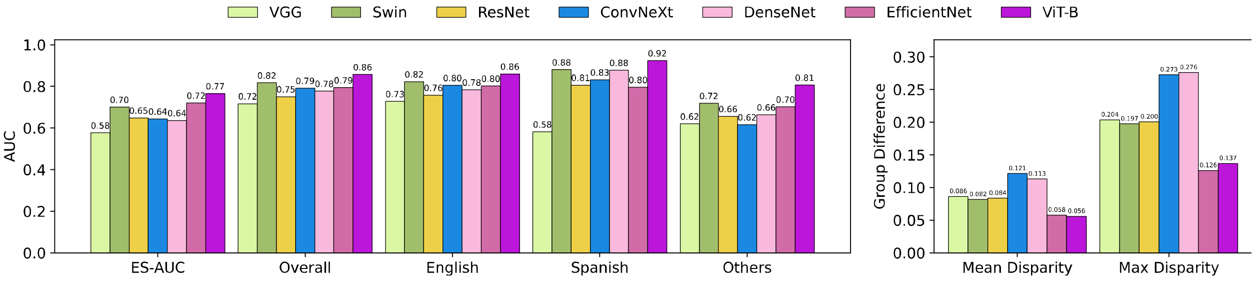


(a)

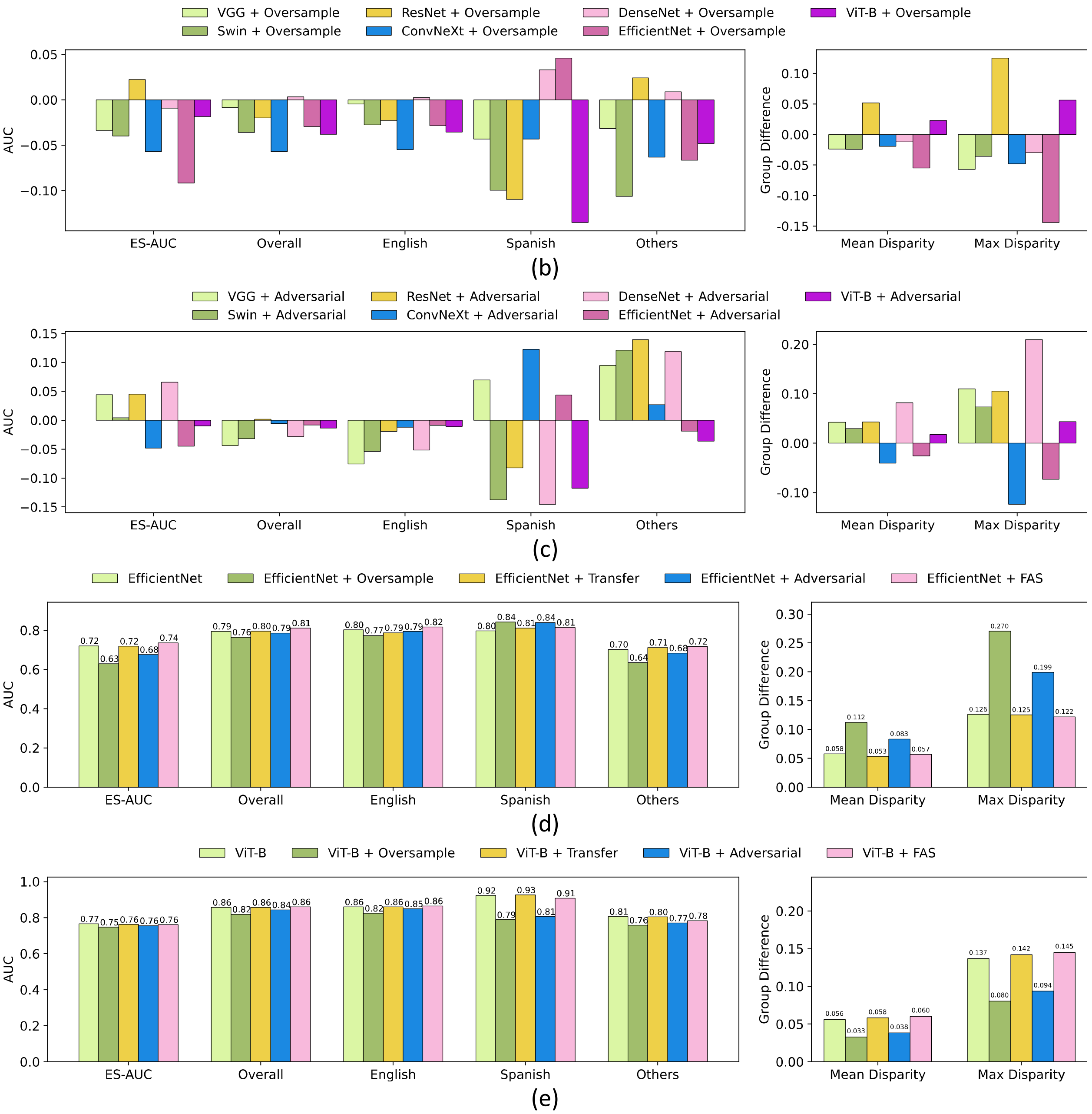


**Figure 16. Results on SLO fundus images on language attributes of the Harvard-FairVision30k dataset.** (a) The accuracy of various baseline models. (b) Differences in accuracy for various baseline models after applying oversampling techniques. (c) Differences in accuracy for various baseline models after applying adversarial techniques. (d) The accuracy of EfficientNet and its integration with oversampling, adversarial, transfer learning and our FAS techniques. (e) The accuracy of ViT-B and its integration with oversampling, adversarial, transfer learning and our FAS techniques.

##### Harvard-FairVision30k SLO Fundus: Marital Status

| | ES-AUC | Overall AUC | Married & Partnered | Single | Divorced | Widowed | Legally Separated | Mean Disparity | Max Disparity |
|---|---|---|---|---|---|---|---|---|---|
| VGG | 0.602 | 0.717 | 0.697 | 0.710 | 0.777 | 0.734 | 0.804 | 0.056 | 0.149 |
| Swin | 0.688 | 0.817 | 0.814 | 0.822 | 0.792 | 0.756 | 0.911 | 0.063 | 0.189 |
| ResNet | 0.617 | 0.750 | 0.747 | 0.753 | 0.746 | 0.670 | 0.875 | 0.088 | 0.274 |
| ConvNeXt | 0.605 | 0.792 | 0.772 | 0.814 | 0.830 | 0.702 | 0.929 | 0.094 | 0.287 |
| DenseNet | 0.633 | 0.777 | 0.786 | 0.777 | 0.745 | 0.670 | 0.857 | 0.078 | 0.241 |
| EfficientNet | 0.683 | 0.794 | 0.796 | 0.806 | 0.800 | 0.662 | 0.804 | 0.070 | 0.182 |
| ViT-B | 0.690 | 0.857 | 0.854 | 0.868 | 0.845 | 0.767 | 0.982 | 0.081 | 0.251 |

**Table 31. The accuracy of various baseline models on marital status attributes for Harvard-FairVision30k SLO fundus images.**

| | ES-AUC | Overall AUC | Married & Partnered | Single | Divorced | Widowed | Legally Separated | Mean Disparity | Max Disparity |
|---|---|---|---|---|---|---|---|---|---|
| VGG* | -0.012 | -0.009 | -0.010 | 0.009 | -0.040 | -0.034 | 0.036 | -0.020 | -0.066 |
| Swin* | -0.139 | -0.036 | -0.046 | -0.002 | -0.056 | -0.082 | 0.089 | -0.079 | -0.229 |
| ResNet* | -0.006 | -0.020 | -0.040 | 0.018 | 0.033 | -0.005 | -0.161 | 0.029 | 0.117 |
| ConvNeXt* | -0.020 | -0.057 | -0.063 | -0.039 | -0.067 | -0.076 | -0.143 | 0.013 | 0.069 |
| DenseNet* | -0.051 | 0.003 | 0.021 | -0.013 | -0.017 | -0.036 | -0.178 | 0.000 | 0.019 |
| EfficientNet* | -0.115 | -0.03 | -0.025 | -0.033 | -0.025 | -0.052 | 0.125 | -0.062 | -0.236 |
| ViT-B* | -0.038 | -0.038 | -0.051 | -0.012 | -0.037 | -0.034 | -0.268 | 0.017 | 0.077 |

**Table 32. Differences in accuracy of various baseline models after applying oversampling techniques on marital status attributes using Harvard-FairVision30k SLO fundus images.** * denotes a model with oversampling techniques.

| | ES-AUC | Overall AUC | Married & Partnered | Single | Divorced | Widowed | Legally Separated | Mean Disparity | Max Disparity |
|---|---|---|---|---|---|---|---|---|---|
| VGG# | -0.055 | -0.044 | -0.023 | -0.029 | -0.045 | -0.130 | -0.036 | -0.027 | -0.094 |
| Swin# | 0.036 | -0.024 | -0.042 | -0.023 | -0.002 | 0.074 | -0.146 | 0.034 | 0.106 |
| ResNet# | 0.065 | 0.006 | 0.029 | 0.036 | 0.009 | 0.061 | -0.148 | 0.055 | 0.191 |
| ConvNeXt# | 0.123 | -0.006 | 0.011 | -0.011 | -0.059 | 0.104 | -0.168 | 0.072 | 0.229 |
| DenseNet# | 0.082 | -0.021 | -0.048 | -0.020 | 0.010 | 0.111 | -0.112 | 0.059 | 0.183 |
| EfficientNet# | -0.059 | -0.011 | -0.012 | -0.008 | 0.000 | 0.063 | 0.143 | -0.023 | -0.101 |
| ViT-B# | 0.016 | -0.020 | -0.024 | 0.015 | 0.008 | -0.024 | -0.125 | 0.023 | 0.083 |

**Table 33. Differences in accuracy of various baseline models after applying adversarial techniques on marital status attributes using Harvard-FairVision30k SLO fundus images.** # denotes a model with adversarial techniques.

| | ES-AUC | Overall AUC | Married & Partnered | Single | Divorced | Widowed | Legally Separated | Mean Disparity | Max Disparity |
|---|---|---|---|---|---|---|---|---|---|
| EfficientNet | 0.683 | 0.794 | 0.796 | 0.806 | 0.800 | 0.662 | 0.804 | 0.070 | 0.182 |
| EfficientNet* | 0.568 | 0.764 | 0.771 | 0.773 | 0.775 | 0.610 | 0.929 | 0.132 | 0.417 |
| EfficientNet¶ | 0.601 | 0.784 | 0.770 | 0.807 | 0.806 | 0.681 | 0.929 | 0.101 | 0.316 |

| | | | | | | | | | |
|---|---|---|---|---|---|---|---|---|---|
| EfficientNet# | 0.624 | 0.783 | 0.784 | 0.798 | 0.800 | 0.725 | 0.946 | 0.093 | 0.283 |
| EfficientNet† | 0.645 | 0.798 | 0.800 | 0.822 | 0.802 | 0.721 | 0.929 | 0.084 | 0.261 |

**Table 34. The performances of EfficientNet and its variants on marital status attributes using Harvard-FairVision30k SLO fundus images.** * denotes EfficientNet with oversampling, ¶ denotes EfficientNet with transfer learning, # denotes EfficientNet with adversarial, and † denotes EfficientNet with our FAS techniques.

| | ES-AUC | Overall AUC | Married & Partnered | Single | Divorced | Widowed | Legally Separated | Mean Disparity | Max Disparity |
|---|---|---|---|---|---|---|---|---|---|
| ViT-B | 0.690 | 0.857 | 0.854 | 0.868 | 0.845 | 0.767 | 0.982 | 0.081 | 0.251 |
| ViT-B* | 0.653 | 0.819 | 0.803 | 0.856 | 0.808 | 0.734 | 0.714 | 0.064 | 0.174 |
| ViT-B¶ | 0.677 | 0.850 | 0.852 | 0.870 | 0.850 | 0.767 | 1.000 | 0.088 | 0.274 |
| ViT-B# | 0.707 | 0.837 | 0.830 | 0.883 | 0.853 | 0.743 | 0.857 | 0.058 | 0.168 |
| ViT-B† | 0.663 | 0.842 | 0.843 | 0.866 | 0.858 | 0.752 | 0.982 | 0.087 | 0.273 |

**Table 35. The performances of ViT-B and its variants on marital status attributes using Harvard-FairVision30k SLO fundus images.** * denotes ViT-B with oversampling, ¶ denotes EfficientNet with transfer learning, # denotes EfficientNet with adversarial, and † denotes ViT-B with our FAS techniques.

**Harvard-FairVision30k SLO Fundus: Gender + Race**

| | ES-AUC | Overall AUC | Female Asian | Female Black | Female White | Male Asian | Male Black | Male White | Mean Disparity | Max Disparity |
|---|---|---|---|---|---|---|---|---|---|---|
| VGG | 0.546 | 0.717 | 0.680 | 0.610 | 0.802 | 0.671 | 0.735 | 0.697 | 0.083 | 0.269 |
| Swin | 0.526 | 0.817 | 0.526 | 0.696 | 0.789 | 0.738 | 0.817 | 0.851 | 0.131 | 0.398 |
| ResNet | 0.590 | 0.750 | 0.656 | 0.646 | 0.714 | 0.729 | 0.756 | 0.762 | 0.060 | 0.155 |
| ConvNeXt | 0.547 | 0.792 | 0.644 | 0.654 | 0.768 | 0.695 | 0.801 | 0.824 | 0.089 | 0.228 |
| DenseNet | 0.496 | 0.777 | 0.398 | 0.689 | 0.714 | 0.790 | 0.781 | 0.793 | 0.178 | 0.509 |
| EfficientNet | 0.597 | 0.794 | 0.835 | 0.713 | 0.715 | 0.701 | 0.796 | 0.829 | 0.071 | 0.169 |
| ViT-B | 0.673 | 0.857 | 0.726 | 0.846 | 0.830 | 0.772 | 0.858 | 0.876 | 0.061 | 0.174 |

**Table 36. The accuracy of various baseline models on gender and race attributes for Harvard-FairVision30k SLO fundus images.**

| | ES-AUC | Overall AUC | Female Asian | Female Black | Female White | Male Asian | Male Black | Male White | Mean Disparity | Max Disparity |
|---|---|---|---|---|---|---|---|---|---|---|
| VGG* | -0.015 | -0.009 | 0.085 | -0.050 | -0.034 | -0.012 | -0.013 | 0.008 | -0.018 | -0.026 |
| Swin* | -0.021 | -0.036 | 0.020 | -0.013 | -0.043 | -0.081 | -0.026 | -0.026 | 0.012 | 0.040 |
| ResNet* | -0.012 | -0.020 | -0.102 | 0.099 | 0.006 | -0.034 | -0.002 | -0.038 | -0.032 | -0.118 |
| ConvNeXt* | -0.041 | -0.057 | -0.100 | -0.047 | -0.025 | -0.027 | -0.083 | -0.048 | -0.020 | -0.089 |
| DenseNet* | 0.049 | 0.003 | 0.248 | -0.051 | 0.015 | -0.048 | -0.020 | 0.031 | 0.095 | 0.271 |
| EfficientNet* | 0.027 | -0.030 | -0.002 | 0.025 | -0.001 | 0.014 | -0.039 | -0.042 | 0.016 | 0.013 |
| ViT-B* | -0.065 | -0.038 | -0.054 | -0.035 | -0.028 | -0.084 | -0.026 | -0.027 | -0.024 | -0.042 |

**Table 37. Differences in accuracy of various baseline models after applying oversampling techniques on gender and race attributes using Harvard-FairVision30k SLO fundus images.** * denotes a model with oversampling techniques.

| | ES-AUC | Overall AUC | Female Asian | Female Black | Female White | Male Asian | Male Black | Male White | Mean Disparity | Max Disparity |
|---|---|---|---|---|---|---|---|---|---|---|
| VGG# | 0.013 | -0.040 | 0.060 | 0.053 | -0.087 | 0.013 | -0.095 | -0.070 | 0.023 | 0.101 |
| Swin# | 0.186 | -0.026 | 0.270 | 0.127 | 0.004 | 0.067 | -0.046 | -0.097 | 0.102 | 0.311 |
| ResNet# | 0.093 | 0.004 | 0.128 | 0.123 | 0.047 | 0.006 | -0.023 | -0.022 | 0.035 | 0.087 |
| ConvNeXt# | 0.144 | -0.007 | 0.175 | 0.127 | 0.047 | 0.087 | -0.056 | -0.066 | 0.054 | 0.134 |
| DenseNet# | 0.180 | -0.022 | 0.336 | 0.062 | 0.057 | -0.004 | -0.063 | -0.030 | 0.148 | 0.419 |
| EfficientNet# | 0.012 | 0.006 | -0.059 | -0.038 | 0.076 | 0.029 | 0.056 | -0.063 | 0.004 | -0.052 |
| ViT-B# | -0.035 | -0.015 | -0.030 | -0.054 | 0.006 | -0.025 | -0.007 | -0.020 | -0.009 | -0.016 |

**Table 38. Differences in accuracy of various baseline models after applying adversarial techniques on gender and race attributes using Harvard-FairVision30k SLO fundus images.** # denotes a model with adversarial techniques.

| | ES-AUC | Overall AUC | Female Asian | Female Black | Female White | Male Asian | Male Black | Male White | Mean Disparity | Max Disparity |
|---|---|---|---|---|---|---|---|---|---|---|
| EfficientNet | 0.597 | 0.794 | 0.835 | 0.713 | 0.715 | 0.701 | 0.796 | 0.829 | 0.071 | 0.169 |
| EfficientNet* | 0.624 | 0.764 | 0.833 | 0.739 | 0.714 | 0.714 | 0.756 | 0.787 | 0.055 | 0.156 |
| EfficientNet# | 0.609 | 0.800 | 0.776 | 0.675 | 0.791 | 0.729 | 0.851 | 0.766 | 0.068 | 0.220 |
| EfficientNet† | 0.615 | 0.805 | 0.709 | 0.746 | 0.748 | 0.737 | 0.826 | 0.813 | 0.052 | 0.146 |

**Table 39. The performances of EfficientNet and its variants on gender and race attributes using Harvard-FairVision30k SLO fundus imges.** * denotes EfficientNet with oversampling, # denotes EfficientNet with adversarial, and † denotes EfficientNet with our FAS techniques.

| | ES-AUC | Overall AUC | Female Asian | Female Black | Female White | Male Asian | Male Black | Male White | Mean Disparity | Max Disparity |
|---|---|---|---|---|---|---|---|---|---|---|
| ViT-B | 0.673 | 0.857 | 0.726 | 0.846 | 0.830 | 0.772 | 0.858 | 0.876 | 0.061 | 0.174 |
| ViT-B* | 0.609 | 0.819 | 0.672 | 0.811 | 0.802 | 0.689 | 0.832 | 0.849 | 0.085 | 0.216 |
| ViT-B# | 0.639 | 0.842 | 0.696 | 0.792 | 0.836 | 0.747 | 0.851 | 0.856 | 0.070 | 0.190 |
| ViT-B† | 0.709 | 0.860 | 0.767 | 0.875 | 0.856 | 0.778 | 0.863 | 0.876 | 0.053 | 0.126 |

**Table 40. The performances of ViT-B and its variants on gender and race attributes using Harvard-FairVision30k SLO fundus imges.** * denotes ViT-B with oversampling, # denotes EfficientNet with adversarial,and † denotes ViT-B with our FAS techniques.

**Harvard-FairVision30k SLO Fundus: Gender + Ethnicity**

| | ES-AUC | Overall AUC | Female Non-Hispanic | Female Hispanic | Female Non-Hispanic | Male Hispanic | Mean Disparity | Max Disparity |
|---|---|---|---|---|---|---|---|---|
| VGG | 0.663 | 0.717 | 0.749 | 0.686 | 0.710 | 0.727 | 0.032 | 0.088 |
| Swin | 0.684 | 0.817 | 0.797 | 0.820 | 0.938 | 0.866 | 0.066 | 0.173 |

| | | | | | | | | |
|---|---|---|---|---|---|---|---|---|
| ResNet | 0.662 | 0.750 | 0.746 | 0.743 | 0.804 | 0.818 | 0.045 | 0.100 |
| ConvNeXt | 0.666 | 0.792 | 0.783 | 0.790 | 0.900 | 0.862 | 0.062 | 0.148 |
| DenseNet | 0.607 | 0.777 | 0.749 | 0.783 | 0.929 | 0.870 | 0.091 | 0.232 |
| EfficientNet | 0.656 | 0.794 | 0.782 | 0.801 | 0.896 | 0.706 | 0.086 | 0.240 |
| ViT-B | 0.775 | 0.857 | 0.849 | 0.857 | 0.915 | 0.896 | 0.032 | 0.078 |

**Table 41. The accuracy of various baseline models on gender and ethnicity attributes for Harvard-FairVision30k SLO fundus images.**

| | ES-AUC | Overall AUC | Female Non-Hispanic | Female Hispanic | Female Non-Hispanic | Male Hispanic | Mean Disparity | Max Disparity |
|---|---|---|---|---|---|---|---|---|
| VGG* | -0.050 | -0.009 | -0.014 | -0.001 | 0.027 | -0.095 | -0.028 | -0.060 |
| Swin* | 0.018 | -0.036 | -0.025 | -0.036 | -0.087 | -0.056 | 0.027 | 0.071 |
| ResNet* | 0.028 | -0.020 | -0.006 | -0.024 | -0.092 | -0.069 | 0.024 | 0.049 |
| ConvNeXt* | -0.039 | -0.057 | -0.066 | -0.050 | -0.089 | -0.056 | 0.007 | 0.019 |
| DenseNet* | 0.034 | 0.003 | 0.000 | 0.007 | -0.094 | 0.030 | 0.019 | 0.038 |
| EfficientNet* | 0.036 | -0.030 | -0.032 | -0.030 | -0.073 | 0.035 | 0.044 | 0.132 |
| ViT-B* | -0.048 | -0.038 | -0.028 | -0.039 | -0.098 | -0.199 | -0.032 | -0.073 |

**Table 42. Differences in accuracy of various baseline models after applying oversampling techniques on gender and ethnicity attributes using Harvard-FairVision30k SLO fundus images.** * denotes a model with oversampling techniques.

| | ES-AUC | Overall AUC | Female Non-Hispanic | Female Hispanic | Female Non-Hispanic | Male Hispanic | Mean Disparity | Max Disparity |
|---|---|---|---|---|---|---|---|---|
| VGG# | -0.052 | -0.044 | -0.042 | -0.021 | -0.020 | -0.097 | -0.010 | -0.025 |
| Swin# | 0.074 | -0.031 | 0.001 | -0.028 | -0.152 | -0.098 | 0.052 | 0.136 |
| ResNet# | 0.054 | 0.018 | 0.029 | 0.015 | -0.013 | -0.082 | 0.019 | 0.029 |
| ConvNeXt# | 0.006 | -0.007 | 0.006 | -0.019 | -0.042 | 0.000 | 0.011 | 0.032 |
| DenseNet# | 0.124 | -0.022 | 0.022 | -0.033 | -0.174 | -0.127 | 0.078 | 0.196 |
| EfficientNet# | 0.047 | 0.015 | 0.016 | 0.010 | 0.029 | 0.082 | 0.017 | 0.071 |
| ViT-B# | -0.016 | -0.014 | -0.002 | -0.018 | -0.025 | -0.108 | -0.011 | -0.044 |

**Table 43. Differences in accuracy of various baseline models after applying adversarial techniques on gender and ethnicity attributes using Harvard-FairVision30k SLO fundus images.** # denotes a model with adversarial techniques.

| | ES-AUC | Overall AUC | Female Non-Hispanic | Female Hispanic | Female Non-Hispanic | Male Hispanic | Mean Disparity | Max Disparity |
|---|---|---|---|---|---|---|---|---|
| EfficientNet | 0.656 | 0.794 | 0.782 | 0.801 | 0.896 | 0.706 | 0.086 | 0.240 |
| EfficientNet* | 0.692 | 0.764 | 0.750 | 0.771 | 0.823 | 0.740 | 0.042 | 0.108 |
| EfficientNet# | 0.704 | 0.809 | 0.798 | 0.810 | 0.925 | 0.788 | 0.068 | 0.169 |
| EfficientNet† | 0.653 | 0.808 | 0.810 | 0.801 | 0.933 | 0.706 | 0.100 | 0.281 |

**Table 44. The performances of EfficientNet and its variants on gender and ethnicity attributes using Harvard-FairVision30k SLO fundus images.** * denotes EfficientNet with

oversampling, # denotes EfficientNet with adversarial, and † denotes EfficientNet with our FAS techniques.

| | ES-AUC | Overall AUC | Female Non-Hispanic | Female Hispanic | Female Non-Hispanic | Male Hispanic | Mean Disparity | Max Disparity |
|---|---|---|---|---|---|---|---|---|
| ViT-B | 0.775 | 0.857 | 0.849 | 0.857 | 0.915 | 0.896 | 0.032 | 0.078 |
| ViT-B* | 0.727 | 0.819 | 0.821 | 0.818 | 0.817 | 0.697 | 0.064 | 0.151 |
| ViT-B# | 0.759 | 0.843 | 0.847 | 0.838 | 0.890 | 0.788 | 0.043 | 0.122 |
| ViT-B† | 0.789 | 0.860 | 0.854 | 0.858 | 0.923 | 0.879 | 0.032 | 0.080 |

**Table 45. The performances of ViT-B and its variants on gender and ethnicity attributes using Harvard-FairVision30k SLO fundus images.** * denotes ViT-B with oversampling, # denotes EfficientNet with adversarial, and † denotes ViT-B with our FAS techniques.

##### Inhouse OCT B-scans: Race

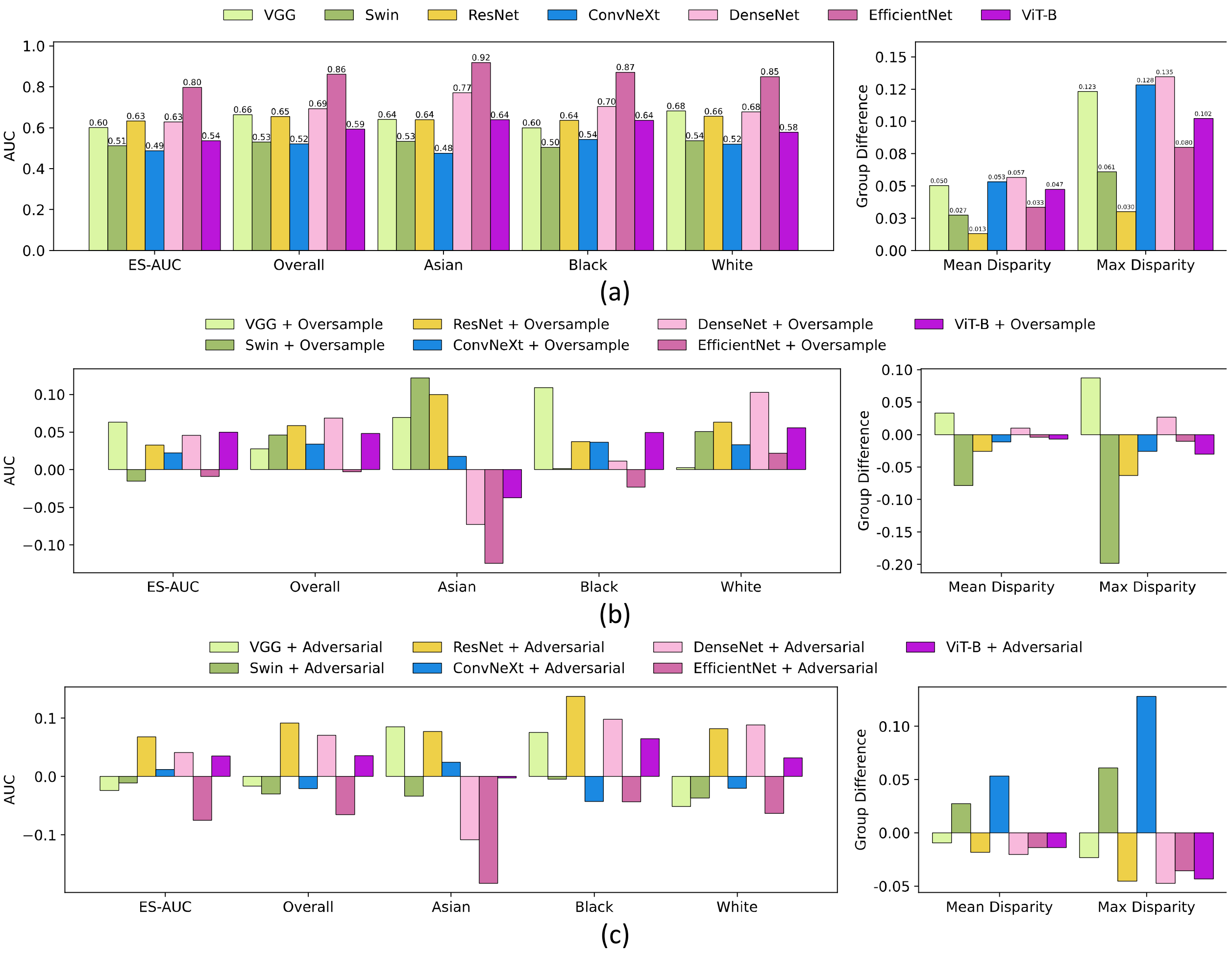

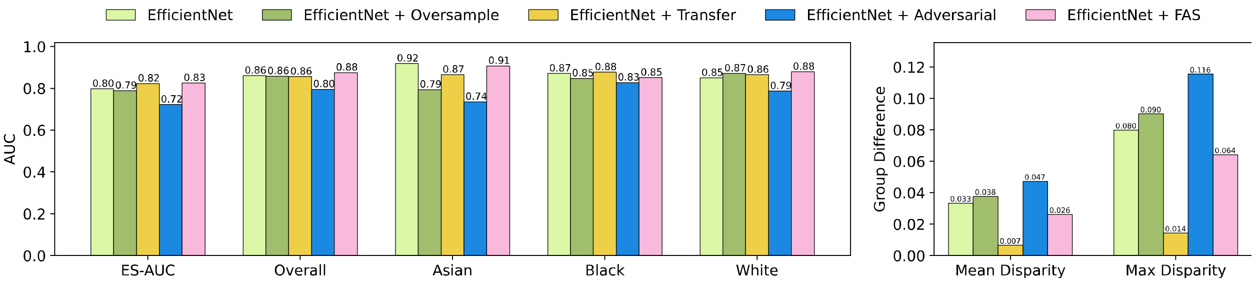


(d)

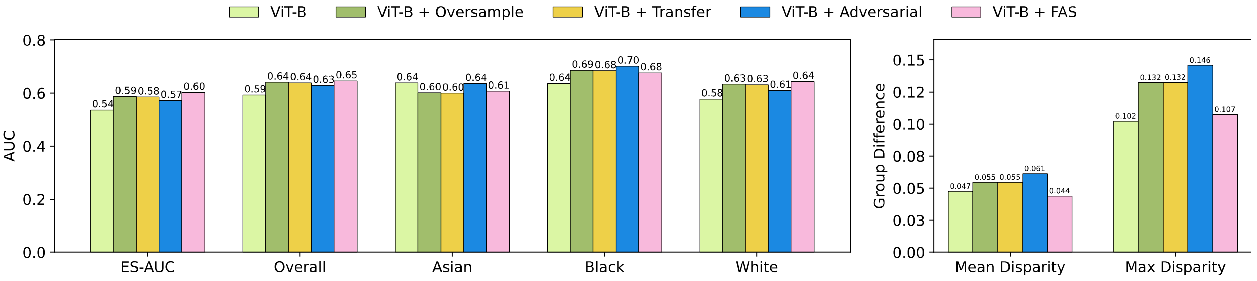


(e)

**Figure 17. Results on OCT B-scans on race attributes of the in-house dataset.** (a) The accuracy of various baseline models. (b) Differences in accuracy for various baseline models after applying oversampling techniques. (c) Differences in accuracy for various baseline models after applying adversarial techniques. (d) The accuracy of EfficientNet and its integration with oversampling, adversarial, transfer learning and our FAS techniques. (e) The accuracy of ViT-B and its integration with oversampling, adversarial, transfer learning and our FAS techniques.

#### Inhouse OCT B-scans: Gender

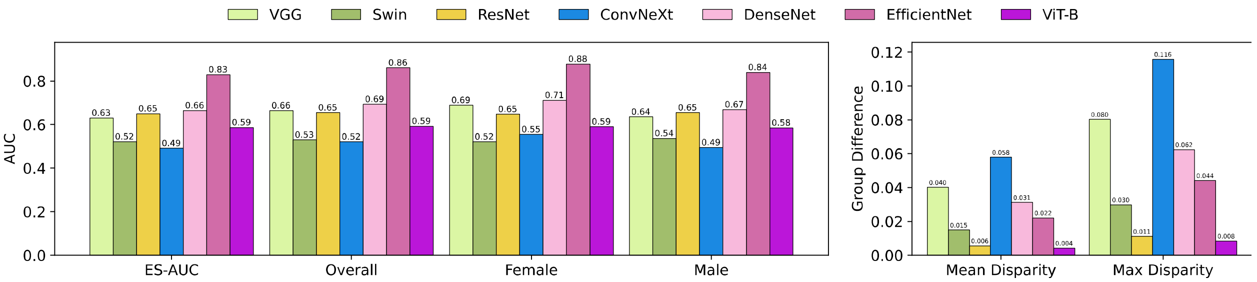


(a)

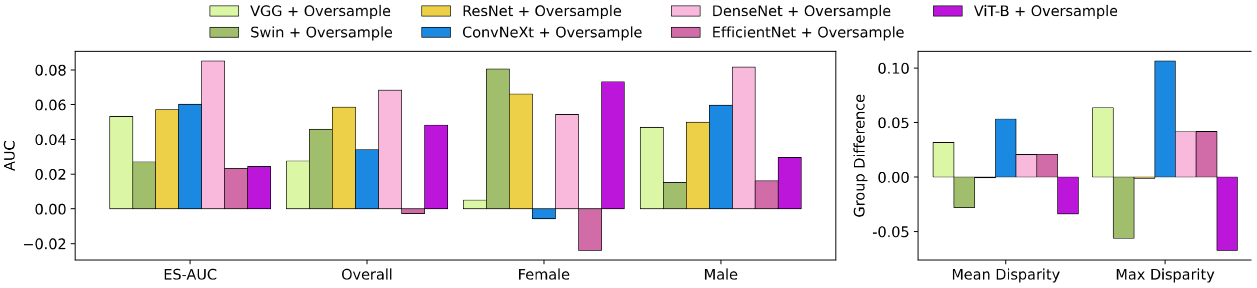


(b)

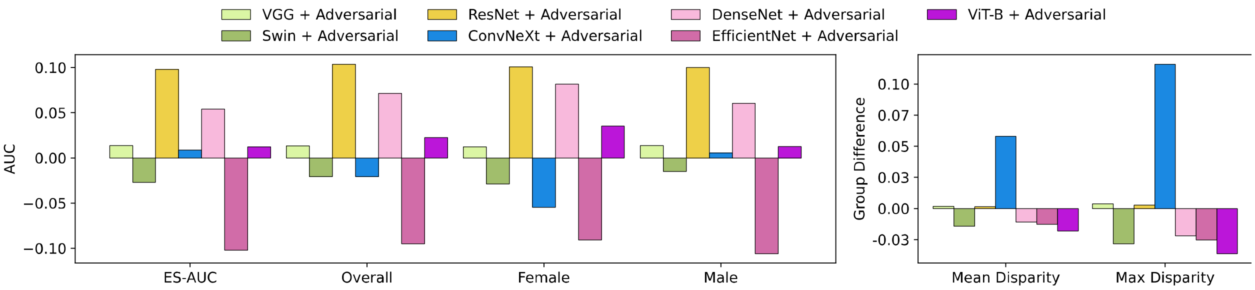


(c)

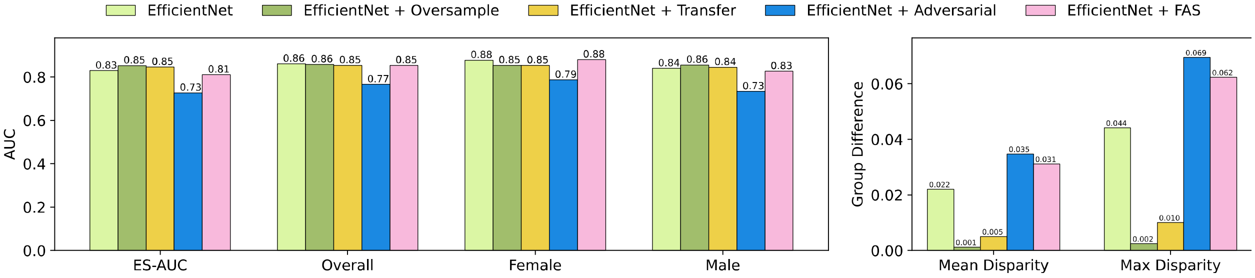


(d)

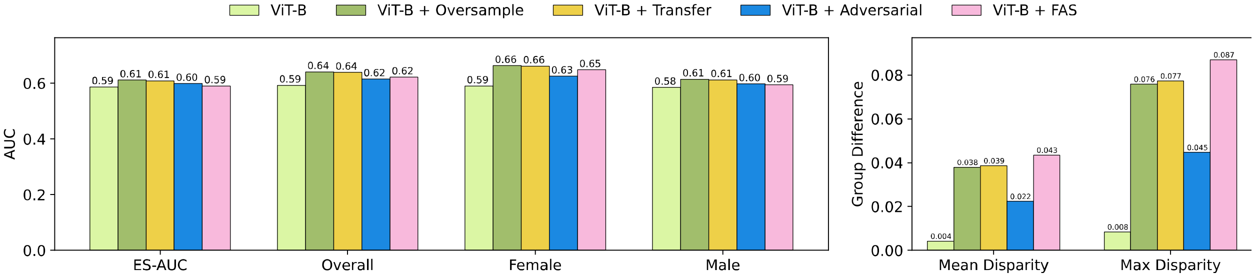


(e)

**Figure 18. Results on OCT B-scans on gender attributes of the in-house dataset.** (a) The accuracy of various baseline models. (b) Differences in accuracy for various baseline models after applying oversampling techniques. (c) Differences in accuracy for various baseline models after applying adversarial techniques. (d) The accuracy of EfficientNet and its integration with oversampling, adversarial, transfer learning and our FAS techniques. (e) The accuracy of ViT-B and its integration with oversampling, adversarial, transfer learning and our FAS techniques.

#### Inhouse OCT B-scans: Ethnicity

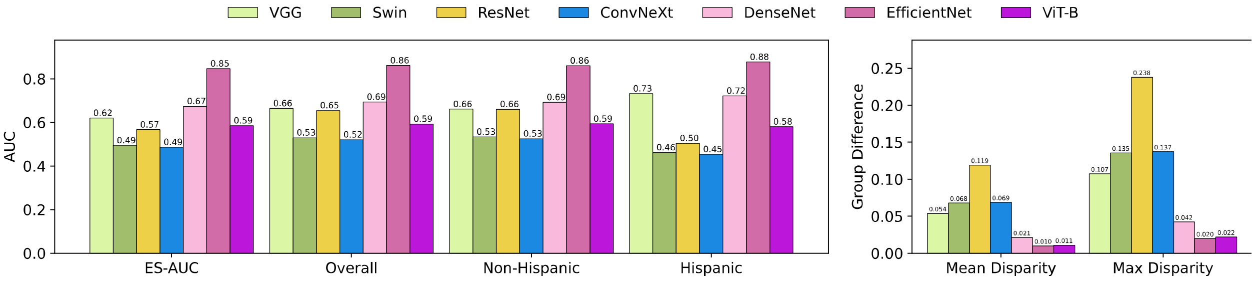


(a)

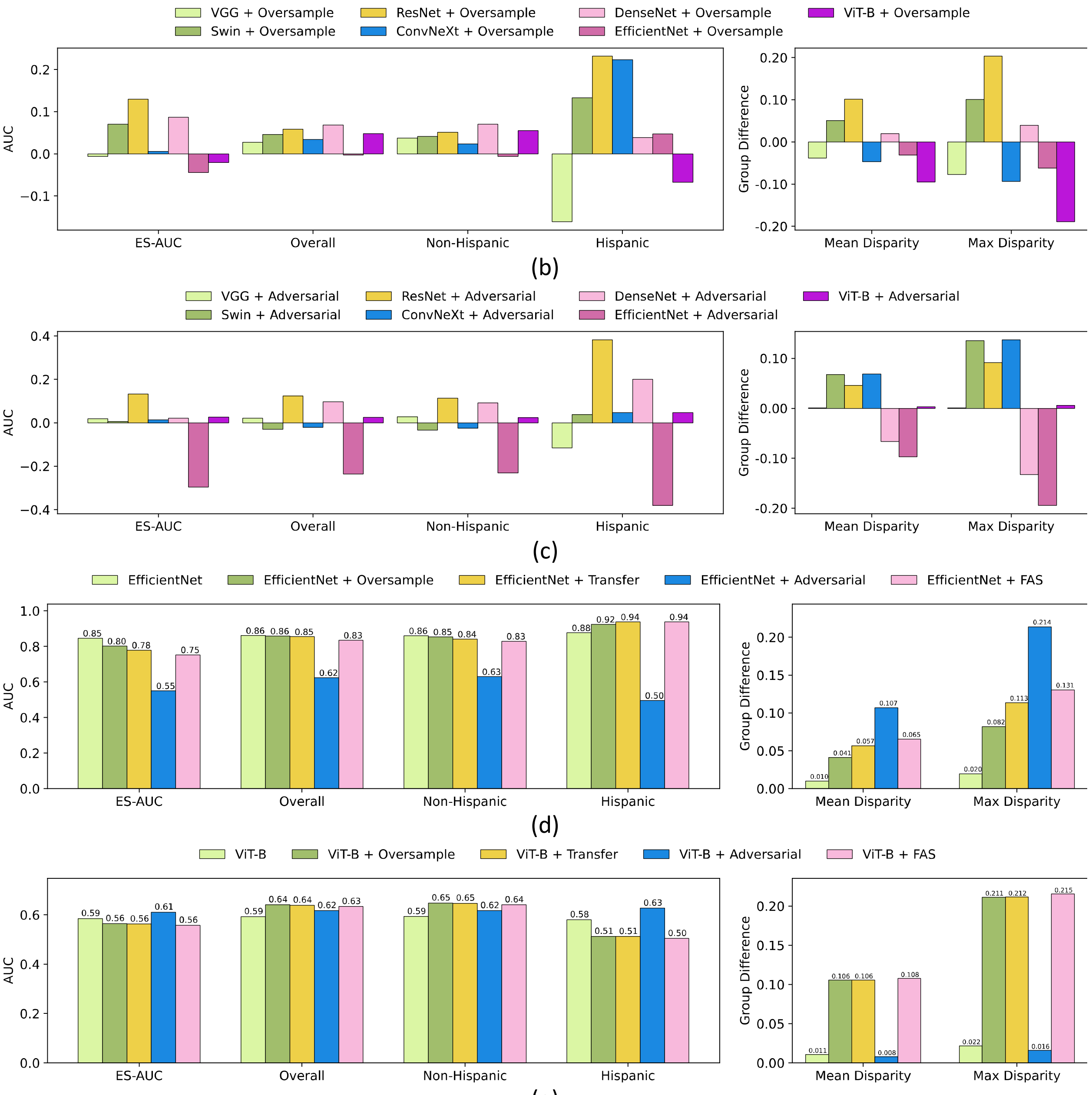


(e)

**Figure 19. Results on OCT B-scans on ethnicity attributes of the in-house dataset.** (a) The accuracy of various baseline models. (b) Differences in accuracy for various baseline models after applying oversampling techniques. (c) Differences in accuracy for various baseline models after applying adversarial techniques. (d) The accuracy of EfficientNet and its integration with oversampling, adversarial, transfer learning and our FAS techniques. (e) The accuracy of ViT-B and its integration with oversampling, adversarial, transfer learning and our FAS techniques.

#### Inhouse OCT B-scans: Language

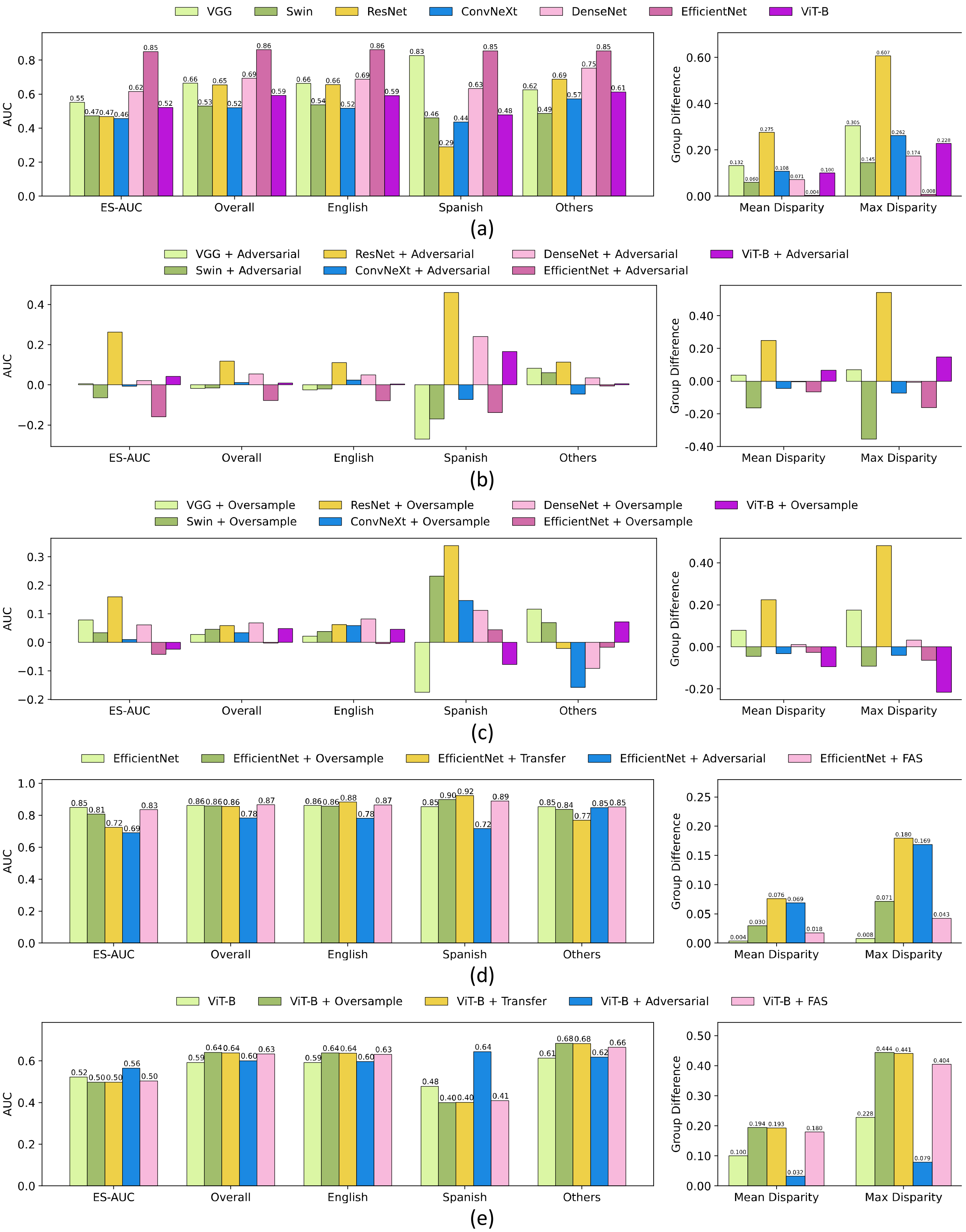


**Figure 20. Results on OCT B-scans on language attributes of the in-house dataset.** (a) The accuracy of various baseline models. (b) Differences in accuracy for various baseline

models after applying oversampling techniques. (c) Differences in accuracy for various baseline models after applying adversarial techniques. (d) The accuracy of EfficientNet and its integration with oversampling, adversarial, transfer learning and our FAS techniques. (e) The accuracy of ViT-B and its integration with oversampling, adversarial, transfer learning and our FAS techniques.

**Inhouse OCT B-scans: Marital Status**

| | ES-AUC | Overall AUC | Married & Partnered | Single | Divorced | Widowed | Legally Separated | Mean Disparity | Max Disparity |
|---|---|---|---|---|---|---|---|---|---|
| VGG | 0.519 | 0.664 | 0.688 | 0.659 | 0.650 | 0.591 | 0.827 | 0.118 | 0.355 |
| Swin | 0.437 | 0.530 | 0.546 | 0.505 | 0.542 | 0.498 | 0.657 | 0.107 | 0.299 |
| ResNet | 0.580 | 0.655 | 0.635 | 0.681 | 0.666 | 0.645 | 0.716 | 0.044 | 0.124 |
| ConvNeXt | 0.454 | 0.521 | 0.535 | 0.492 | 0.526 | 0.449 | 0.494 | 0.058 | 0.165 |
| DenseNet | 0.652 | 0.693 | 0.695 | 0.668 | 0.709 | 0.695 | 0.714 | 0.023 | 0.065 |
| EfficientNet | 0.752 | 0.861 | 0.855 | 0.851 | 0.937 | 0.864 | 0.910 | 0.040 | 0.100 |
| ViT-B | 0.419 | 0.593 | 0.592 | 0.622 | 0.668 | 0.493 | 0.801 | 0.170 | 0.520 |

**Table 46. The accuracy of various baseline models on marital status attributes for in-house OCT B-scans.**

| | ES-AUC | Overall AUC | Married & Partnered | Single | Divorced | Widowed | Legally Separated | Mean Disparity | Max Disparity |
|---|---|---|---|---|---|---|---|---|---|
| VGG* | 0.008 | 0.028 | 0.019 | 0.078 | 0.016 | -0.025 | -0.236 | 0.024 | 0.108 |
| Swin* | -0.005 | 0.046 | 0.078 | -0.003 | 0.015 | -0.113 | -0.083 | -0.035 | -0.117 |
| ResNet* | 0.026 | 0.059 | 0.088 | 0.034 | 0.049 | -0.036 | 0.056 | -0.031 | -0.104 |
| ConvNeXt* | 0.038 | 0.034 | 0.026 | 0.047 | 0.068 | 0.098 | 0.120 | 0.007 | 0.032 |
| DenseNet* | 0.017 | 0.068 | 0.072 | 0.070 | 0.115 | 0.056 | 0.086 | -0.019 | -0.047 |
| EfficientNet* | -0.069 | -0.003 | 0.003 | 0.003 | -0.006 | -0.103 | 0.028 | -0.035 | -0.108 |
| ViT-B* | 0.069 | 0.048 | 0.062 | 0.046 | 0.010 | 0.040 | -0.034 | 0.053 | 0.154 |

**Table 47. Differences in accuracy of various baseline models after applying oversampling techniques on marital status attributes using in-house OCT B-scans.** * denotes a model with oversampling techniques.

| | ES-AUC | Overall AUC | Married & Partnered | Single | Divorced | Widowed | Legally Separated | Mean Disparity | Max Disparity |
|---|---|---|---|---|---|---|---|---|---|
| VGG# | -0.014 | -0.024 | -0.025 | -0.018 | -0.103 | -0.047 | -0.242 | 0.043 | 0.170 |
| Swin# | -0.014 | 0.017 | 0.025 | 0.043 | -0.091 | -0.037 | -0.192 | 0.015 | 0.078 |
| ResNet# | -0.056 | 0.080 | 0.069 | 0.103 | 0.089 | 0.169 | 0.239 | -0.072 | -0.218 |
| ConvNeXt# | -0.116 | 0.010 | 0.007 | 0.023 | 0.179 | 0.038 | -0.289 | -0.246 | -0.776 |
| DenseNet# | -0.042 | 0.079 | 0.080 | 0.107 | 0.043 | 0.036 | 0.258 | -0.090 | -0.247 |
| EfficientNet# | -0.015 | -0.068 | -0.056 | -0.077 | -0.113 | -0.060 | -0.110 | 0.020 | 0.036 |
| ViT-B# | 0.035 | 0.015 | 0.021 | -0.011 | -0.086 | 0.054 | 0.051 | -0.008 | 0.018 |

**Table 48. Differences in accuracy of various baseline models after applying adversarial techniques on marital status attributes using in-house OCT B-scans.** # denotes a model with adversarial techniques.

| | ES-AUC | Overall AUC | Married & Partnered | Single | Divorced | Widowed | Legally Separated | Mean Disparity | Max Disparity |
|---|---|---|---|---|---|---|---|---|---|
| EfficientNet | 0.752 | 0.861 | 0.855 | 0.851 | 0.937 | 0.864 | 0.910 | 0.040 | 0.100 |
| EfficientNet* | 0.683 | 0.858 | 0.858 | 0.854 | 0.931 | 0.760 | 0.939 | 0.075 | 0.208 |
| EfficientNet¶ | 0.661 | 0.856 | 0.845 | 0.868 | 0.960 | 0.884 | 0.994 | 0.067 | 0.175 |
| EfficientNet# | 0.737 | 0.792 | 0.798 | 0.774 | 0.824 | 0.803 | 0.800 | 0.020 | 0.063 |
| EfficientNet† | 0.668 | 0.841 | 0.825 | 0.888 | 0.832 | 0.771 | 0.959 | 0.076 | 0.223 |

**Table 49. The performances of EfficientNet and its variants on marital status attributes using in-house OCT B-scans.** * denotes EfficientNet with oversampling, ¶ denotes EfficientNet with transfer learning, # denotes EfficientNet with adversarial, and † denotes EfficientNet with our FAS techniques.

| | ES-AUC | Overall AUC | Married & Partnered | Single | Divorced | Widowed | Legally Separated | Mean Disparity | Max Disparity |
|---|---|---|---|---|---|---|---|---|---|
| ViT-B | 0.419 | 0.593 | 0.592 | 0.622 | 0.668 | 0.493 | 0.801 | 0.170 | 0.520 |
| ViT-B* | 0.489 | 0.641 | 0.654 | 0.668 | 0.678 | 0.533 | 0.767 | 0.117 | 0.366 |
| ViT-B¶ | 0.512 | 0.646 | 0.652 | 0.667 | 0.672 | 0.534 | 0.744 | 0.105 | 0.326 |
| ViT-B# | 0.454 | 0.608 | 0.613 | 0.611 | 0.582 | 0.547 | 0.852 | 0.178 | 0.502 |
| ViT-B† | 0.480 | 0.653 | 0.664 | 0.654 | 0.678 | 0.535 | 0.858 | 0.158 | 0.494 |

**Table 50. The performances of ViT-B and its variants on marital status attributes using in-house OCT B-scans.** * denotes ViT-B with oversampling, ¶ denotes EfficientNet with transfer learning, # denotes EfficientNet with adversarial, and † denotes ViT-B with our FAS techniques.

**Inhouse OCT B-scans: Gender + Race**

| | ES-AUC | Overall AUC | Female Asian | Female Black | Female White | Male Asian | Male Black | Male White | Mean Disparity | Max Disparity |
|---|---|---|---|---|---|---|---|---|---|---|
| VGG | 0.482 | 0.664 | 0.752 | 0.559 | 0.646 | 0.530 | 0.696 | 0.663 | 0.115 | 0.334 |
| Swin | 0.402 | 0.530 | 0.684 | 0.464 | 0.514 | 0.486 | 0.514 | 0.553 | 0.135 | 0.415 |
| ResNet | 0.541 | 0.655 | 0.595 | 0.654 | 0.683 | 0.574 | 0.631 | 0.672 | 0.060 | 0.166 |
| ConvNeXt | 0.430 | 0.521 | 0.482 | 0.484 | 0.596 | 0.502 | 0.542 | 0.501 | 0.078 | 0.220 |
| DenseNet | 0.491 | 0.693 | 0.945 | 0.722 | 0.750 | 0.662 | 0.680 | 0.662 | 0.142 | 0.408 |
| EfficientNet | 0.727 | 0.861 | 0.938 | 0.906 | 0.864 | 0.865 | 0.876 | 0.822 | 0.042 | 0.134 |
| ViT-B | 0.467 | 0.593 | 0.710 | 0.597 | 0.692 | 0.598 | 0.557 | 0.585 | 0.096 | 0.259 |

**Table 51. The accuracy of various baseline models on gender and race attributes for in-house OCT B-scans.**

| | ES-AUC | Overall | Female | Female | Female | Male | Male | Male | Mean | Max |
|---|---|---|---|---|---|---|---|---|---|---|

| | | AUC | Asian | Black | White | Asian | Black | White | Disparity | Disparity |
|---|---|---|---|---|---|---|---|---|---|---|
| VGG* | 0.132 | 0.028 | 0.003 | 0.117 | 0.073 | 0.163 | -0.018 | 0.020 | 0.075 | 0.222 |
| Swin* | 0.020 | 0.046 | -0.046 | 0.189 | -0.035 | 0.044 | 0.116 | -0.006 | 0.023 | 0.113 |
| ResNet* | 0.074 | 0.059 | 0.175 | 0.056 | -0.013 | 0.096 | 0.090 | 0.037 | 0.013 | 0.026 |
| ConvNeXt* | -0.041 | 0.034 | -0.224 | 0.097 | 0.026 | 0.024 | 0.004 | 0.055 | -0.137 | -0.437 |
| DenseNet* | 0.084 | 0.068 | -0.226 | -0.063 | 0.021 | -0.048 | 0.081 | 0.121 | 0.060 | 0.185 |
| EfficientNet* | -0.034 | -0.003 | -0.029 | -0.184 | -0.007 | -0.031 | -0.023 | 0.057 | -0.026 | -0.083 |
| ViT-B* | 0.020 | 0.048 | -0.025 | -0.041 | 0.078 | 0.006 | 0.073 | 0.045 | -0.009 | -0.076 |

**Table 52. Differences in accuracy of various baseline models after applying oversampling techniques on gender and race attributes using in-house OCT B-scans.** * denotes a model with oversampling techniques.

| | ES-AUC | Overall AUC | Female Asian | Female Black | Female White | Male Asian | Male Black | Male White | Mean Disparity | Max Disparity |
|---|---|---|---|---|---|---|---|---|---|---|
| VGG# | 0.050 | 0.023 | -0.030 | 0.193 | 0.155 | 0.156 | 0.008 | -0.034 | 0.037 | 0.083 |
| Swin# | -0.009 | -0.018 | -0.135 | 0.141 | 0.102 | 0.007 | 0.003 | -0.083 | 0.029 | 0.130 |
| ResNet# | 0.051 | 0.106 | 0.068 | 0.089 | 0.131 | 0.098 | 0.155 | 0.086 | -0.012 | -0.032 |
| ConvNeXt# | -0.031 | 0.027 | 0.223 | 0.019 | 0.019 | 0.030 | -0.050 | 0.079 | -0.057 | -0.169 |
| DenseNet# | 0.072 | 0.095 | -0.216 | -0.033 | 0.148 | 0.076 | 0.147 | 0.086 | 0.053 | 0.143 |
| EfficientNet# | -0.251 | -0.201 | -0.397 | -0.416 | -0.206 | -0.184 | -0.167 | -0.187 | -0.076 | -0.198 |
| ViT-B# | -0.002 | 0.029 | 0.129 | 0.028 | -0.01 | 0.005 | 0.036 | 0.027 | -0.041 | -0.137 |

**Table 53. Differences in accuracy of various baseline models after applying adversarial techniques on gender and race attributes using in-house OCT B-scans.** # denotes a model with adversarial techniques.

| | ES-AUC | Overall AUC | Female Asian | Female Black | Female White | Male Asian | Male Black | Male White | Mean Disparity | Max Disparity |
|---|---|---|---|---|---|---|---|---|---|---|
| EfficientNet | 0.727 | 0.861 | 0.938 | 0.906 | 0.864 | 0.865 | 0.876 | 0.822 | 0.042 | 0.134 |
| EfficientNet* | 0.693 | 0.858 | 0.909 | 0.722 | 0.857 | 0.834 | 0.852 | 0.879 | 0.068 | 0.217 |
| EfficientNet# | 0.476 | 0.660 | 0.540 | 0.490 | 0.658 | 0.680 | 0.709 | 0.635 | 0.118 | 0.332 |
| EfficientNet† | 0.719 | 0.868 | 0.974 | 0.884 | 0.843 | 0.862 | 0.893 | 0.839 | 0.052 | 0.155 |

**Table 54. The performances of EfficientNet and its variants on gender and race attributes using in-house OCT B-scans.** * denotes EfficientNet with oversampling, # denotes EfficientNet with transfer learning, and † denotes EfficientNet with our FAS techniques.

| | ES-AUC | Overall AUC | Female Asian | Female Black | Female White | Male Asian | Male Black | Male White | Mean Disparity | Max Disparity |
|---|---|---|---|---|---|---|---|---|---|---|
| ViT-B | 0.467 | 0.593 | 0.710 | 0.597 | 0.692 | 0.598 | 0.557 | 0.585 | 0.096 | 0.259 |
| ViT-B* | 0.486 | 0.641 | 0.685 | 0.556 | 0.770 | 0.604 | 0.630 | 0.630 | 0.105 | 0.334 |
| ViT-B# | 0.464 | 0.621 | 0.839 | 0.625 | 0.682 | 0.603 | 0.593 | 0.612 | 0.137 | 0.396 |
| ViT-B† | 0.498 | 0.636 | 0.661 | 0.568 | 0.756 | 0.590 | 0.630 | 0.624 | 0.095 | 0.294 |

**Table 55. The performances of ViT-B and its variants on gender and race attributes using in-house OCT B-scans.** * denotes ViT-B with oversampling, # denotes ViT-B with transfer learning, and † denotes ViT-B with our FAS techniques.

**Inhouse OCT B-scans: Gender + Ethnicity**

| | ES-AUC | Overall AUC | Female Non-Hispanic | Female Hispanic | Female Non-Hispanic | Male Hispanic | Mean Disparity | Max Disparity |
|---|---|---|---|---|---|---|---|---|
| VGG | 0.482 | 0.664 | 0.696 | 0.623 | 0.593 | 0.898 | 0.179 | 0.458 |
| Swin | 0.455 | 0.530 | 0.522 | 0.542 | 0.492 | 0.424 | 0.085 | 0.224 |
| ResNet | 0.521 | 0.655 | 0.657 | 0.656 | 0.426 | 0.631 | 0.147 | 0.352 |
| ConvNeXt | 0.426 | 0.521 | 0.567 | 0.493 | 0.381 | 0.529 | 0.133 | 0.357 |
| DenseNet | 0.602 | 0.693 | 0.714 | 0.664 | 0.671 | 0.773 | 0.062 | 0.157 |
| EfficientNet | 0.799 | 0.861 | 0.878 | 0.838 | 0.862 | 0.896 | 0.025 | 0.068 |
| ViT-B | 0.456 | 0.593 | 0.599 | 0.580 | 0.467 | 0.748 | 0.169 | 0.475 |

**Table 56. The accuracy of various baseline models on gender and ethnicity attributes for in-house OCT B-scans.**

| | ES-AUC | Overall AUC | Female Non-Hispanic | Female Hispanic | Female Non-Hispanic | Male Hispanic | Mean Disparity | Max Disparity |
|---|---|---|---|---|---|---|---|---|
| VGG* | 0.007 | 0.028 | 0.016 | 0.055 | -0.180 | -0.103 | -0.027 | -0.092 |
| Swin* | 0.023 | 0.046 | 0.084 | 0.005 | 0.044 | 0.256 | -0.014 | -0.026 |
| ResNet* | 0.160 | 0.059 | 0.054 | 0.047 | 0.309 | 0.094 | 0.130 | 0.306 |
| ConvNeXt* | 0.017 | 0.034 | -0.032 | 0.061 | 0.383 | 0.002 | -0.042 | -0.063 |
| DenseNet* | 0.103 | 0.068 | 0.056 | 0.084 | 0.053 | 0.010 | 0.033 | 0.080 |
| EfficientNet* | -0.046 | -0.003 | -0.031 | 0.016 | 0.080 | 0.003 | -0.019 | -0.042 |
| ViT-B* | 0.048 | 0.048 | 0.081 | 0.035 | -0.023 | -0.118 | 0.030 | 0.106 |

**Table 57. Differences in accuracy of various baseline models after applying oversampling techniques on gender and ethnicity attributes using in-house OCT B-scans.** * denotes a model with oversampling techniques.

| | ES-AUC | Overall AUC | Female Non-Hispanic | Female Hispanic | Female Non-Hispanic | Male Hispanic | Mean Disparity | Max Disparity |
|---|---|---|---|---|---|---|---|---|
| VGG# | 0.084 | 0.017 | 0.026 | 0.022 | -0.037 | -0.217 | 0.090 | 0.216 |
| Swin# | -0.018 | 0.022 | 0.068 | -0.026 | -0.010 | 0.248 | -0.047 | -0.119 |
| ResNet# | 0.069 | 0.086 | 0.116 | 0.048 | 0.438 | 0.174 | 0.069 | 0.135 |
| ConvNeXt# | 0.039 | 0.043 | 0.009 | 0.064 | 0.063 | 0.107 | 0.010 | 0.016 |
| DenseNet# | 0.064 | 0.105 | 0.114 | 0.099 | 0.254 | 0.019 | -0.014 | -0.047 |
| EfficientNet# | -0.156 | -0.149 | -0.128 | -0.133 | -0.122 | -0.218 | -0.015 | -0.034 |
| ViT-B# | 0.015 | 0.039 | 0.055 | 0.027 | -0.021 | -0.009 | 0.000 | 0.010 |

**Table 58. Differences in accuracy of various baseline models after applying adversarial techniques on gender and ethnicity attributes using in-house OCT B-scans.** # denotes a model with adversarial techniques.

| | ES-AUC | Overall AUC | Female Non-Hispanic | Female Hispanic | Female Non-Hispanic | Male Hispanic | Mean Disparity | Max Disparity |
|---|---|---|---|---|---|---|---|---|
| EfficientNet | 0.799 | 0.861 | 0.878 | 0.838 | 0.862 | 0.896 | 0.025 | 0.068 |

| | | | | | | | | |
|---|---|---|---|---|---|---|---|---|
| EfficientNet* | 0.752 | 0.858 | 0.847 | 0.853 | 0.941 | 0.900 | 0.045 | 0.110 |
| EfficientNet# | 0.643 | 0.712 | 0.751 | 0.705 | 0.740 | 0.678 | 0.040 | 0.102 |
| EfficientNet† | 0.708 | 0.865 | 0.881 | 0.840 | 0.940 | 0.972 | 0.059 | 0.153 |

**Table 59. The performances of EfficientNet and its variants on gender and ethnicity attributes using in-house OCT B-scans.** * denotes EfficientNet with oversampling, # denotes EfficientNet with transfer learning, and † denotes EfficientNet with our FAS techniques.

| | ES-AUC | Overall AUC | Female Non-Hispanic | Female Hispanic | Female Non-Hispanic | Male Hispanic | Mean Disparity | Max Disparity |
|---|---|---|---|---|---|---|---|---|
| ViT-B | 0.456 | 0.593 | 0.599 | 0.580 | 0.467 | 0.748 | 0.169 | 0.475 |
| ViT-B* | 0.504 | 0.641 | 0.680 | 0.615 | 0.444 | 0.630 | 0.139 | 0.368 |
| ViT-B# | 0.471 | 0.631 | 0.654 | 0.606 | 0.446 | 0.739 | 0.169 | 0.464 |
| ViT-B† | 0.495 | 0.650 | 0.684 | 0.627 | 0.444 | 0.599 | 0.137 | 0.370 |

**Table 60. The performances of ViT-B and its variants on gender and ethnicity attributes using in-house OCT B-scans.** * denotes ViT-B with oversampling, # denotes ViT-B with transfer learning, and † denotes ViT-B with our FAS techniques.

**Harvard-FairVision30k OCT B-scans: Race**

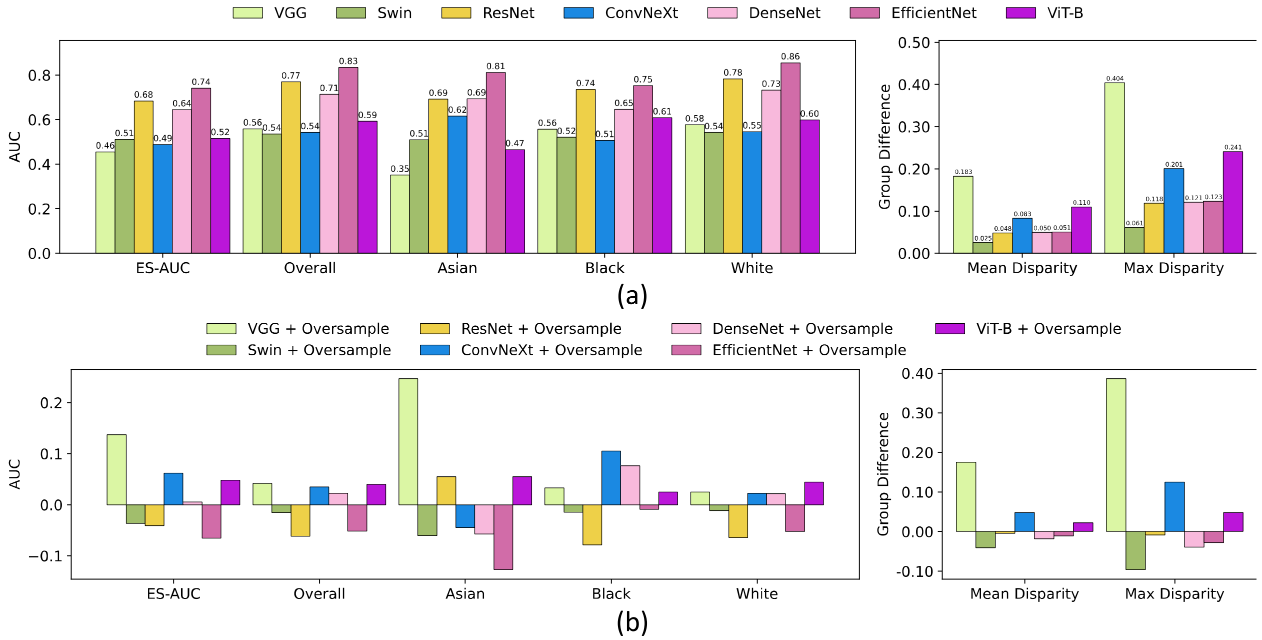

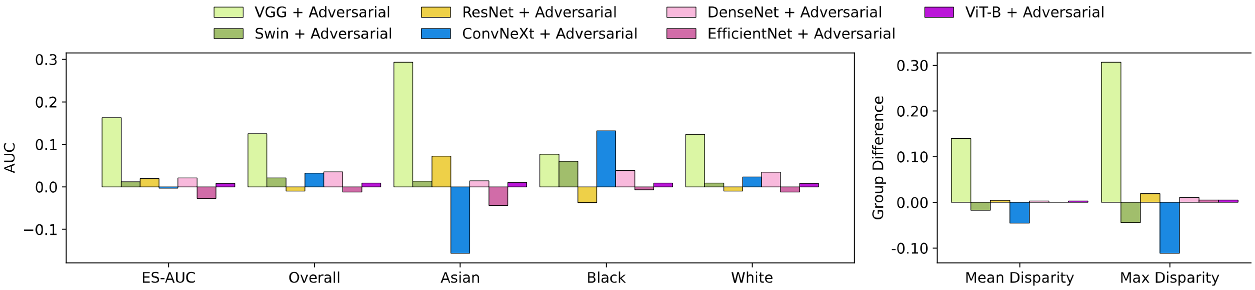


(c)

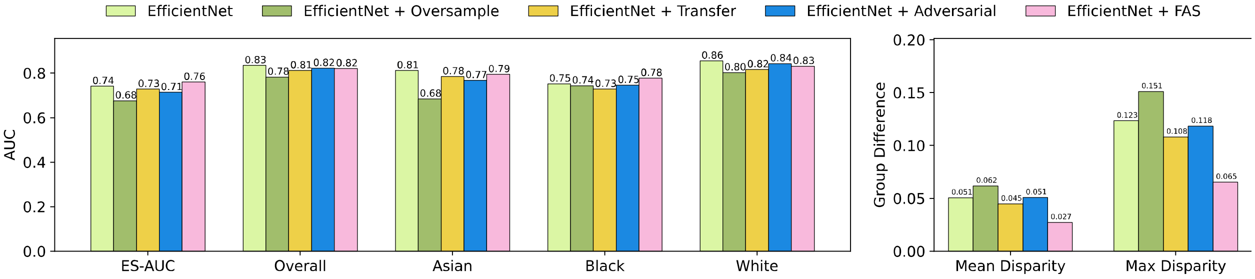


(d)

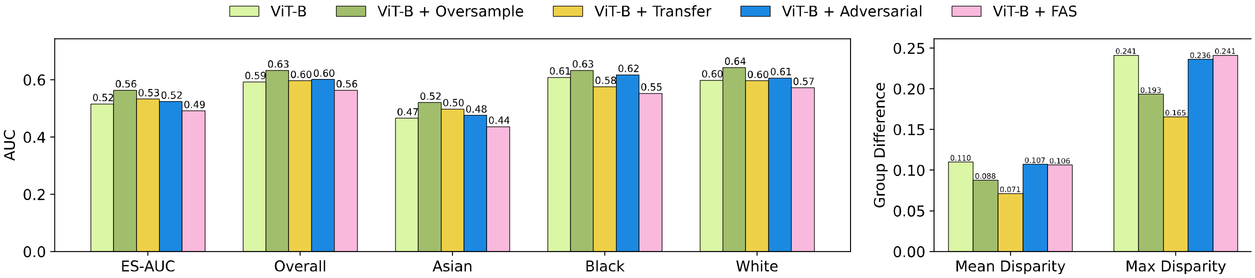


(e)

**Figure 21. Results on OCT B-scans on race attributes of the Harvard-FairVision30k dataset.** (a) The accuracy of various baseline models. (b) Differences in accuracy for various baseline models after applying oversampling techniques. (c) Differences in accuracy for various baseline models after applying adversarial techniques. (d) The accuracy of EfficientNet and its integration with oversampling, adversarial, transfer learning and our FAS techniques. (e) The accuracy of ViT-B and its integration with oversampling, adversarial, transfer learning and our FAS techniques.

##### Harvard-FairVision30k OCT B-scans: Gender

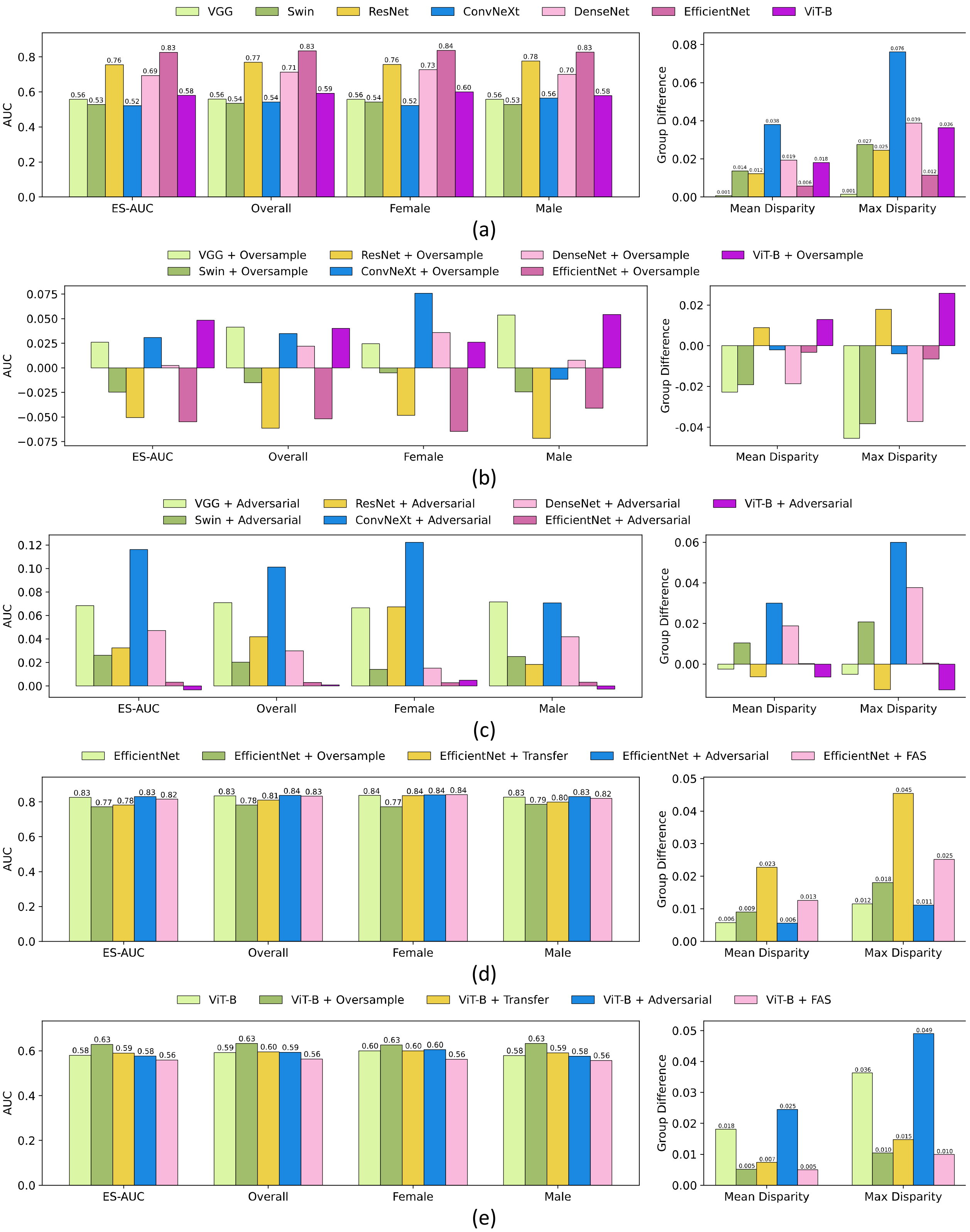


**Figure 22. Results on OCT B-scans on gender attributes of the Harvard-FairVision30k dataset.** (a) The accuracy of various baseline models. (b) Differences in accuracy for various

baseline models after applying oversampling techniques. (c) Differences in accuracy for various baseline models after applying adversarial techniques. (d) The accuracy of EfficientNet and its integration with oversampling, adversarial, transfer learning and our FAS techniques. (e) The accuracy of ViT-B and its integration with oversampling, adversarial, transfer learning and our FAS techniques.

#### Harvard-FairVision30k OCT B-scans: Ethnicity

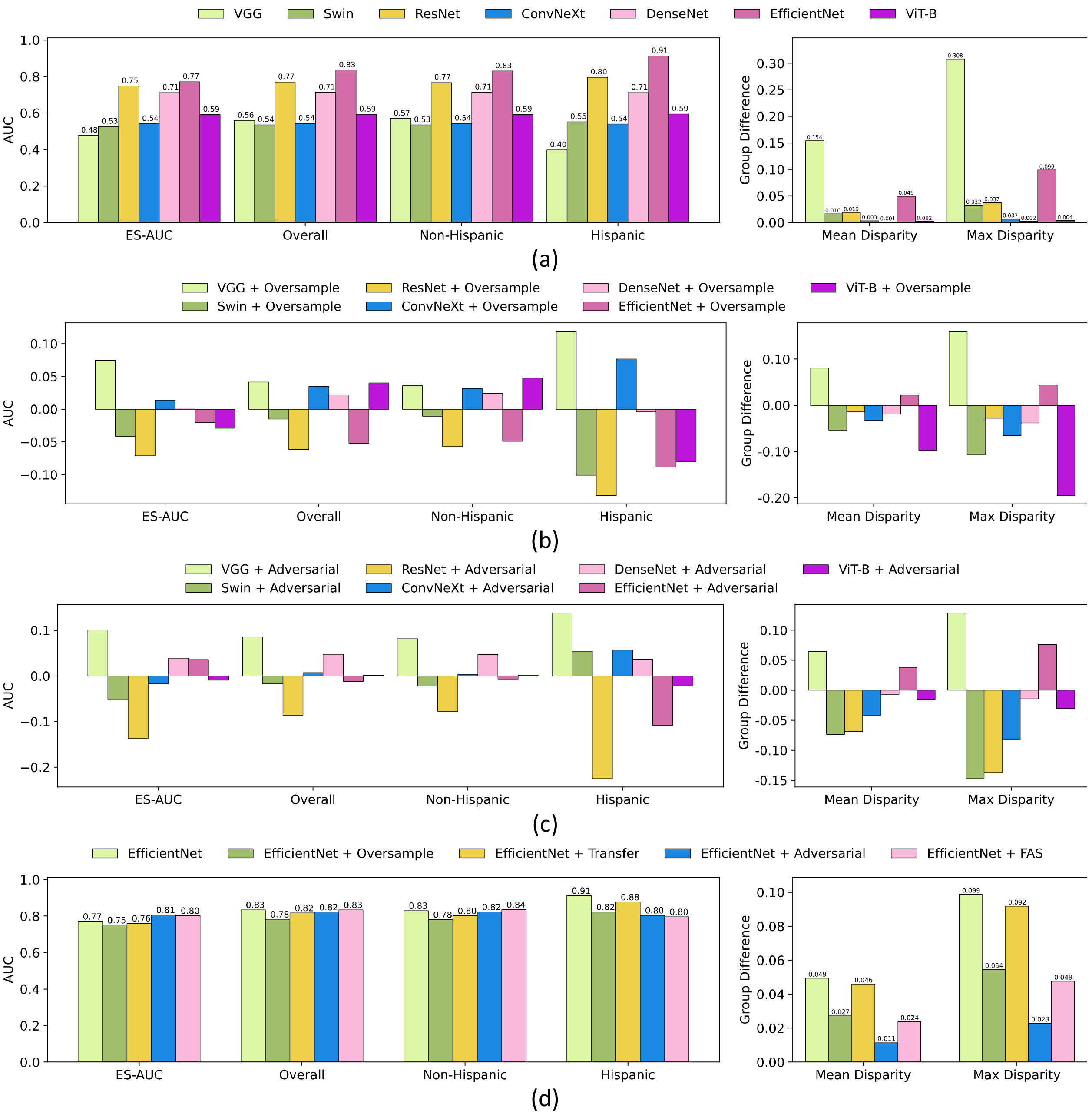

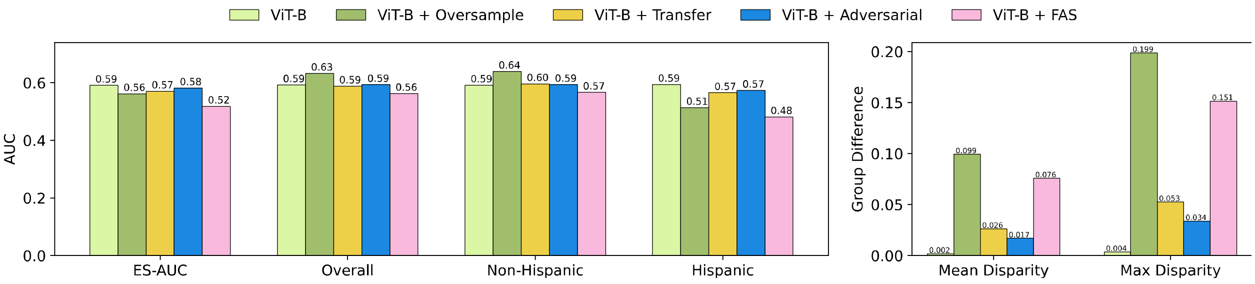


(e)

**Figure 23. Results on OCT B-scans on ethnicity attributes of the Harvard-FairVision30k dataset.** (a) The accuracy of various baseline models. (b) Differences in accuracy for various baseline models after applying oversampling techniques. (c) Differences in accuracy for various baseline models after applying adversarial techniques. (d) The accuracy of EfficientNet and its integration with oversampling, adversarial, transfer learning and our FAS techniques. (e) The accuracy of ViT-B and its integration with oversampling, adversarial, transfer learning and our FAS techniques.

#### Harvard-FairVision30k OCT B-scans: Language

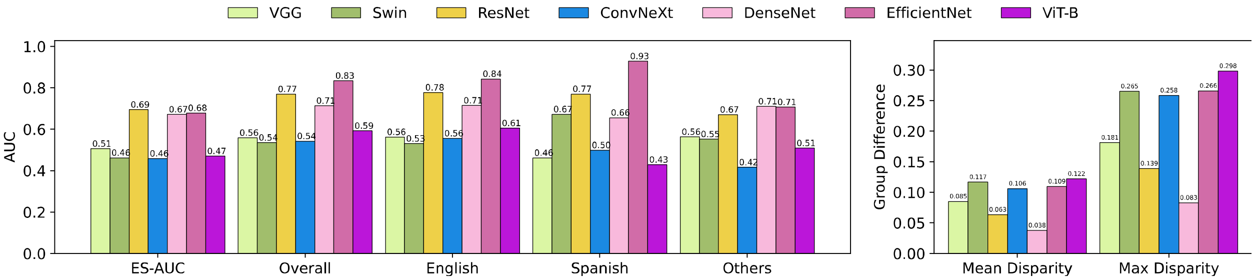


(a)

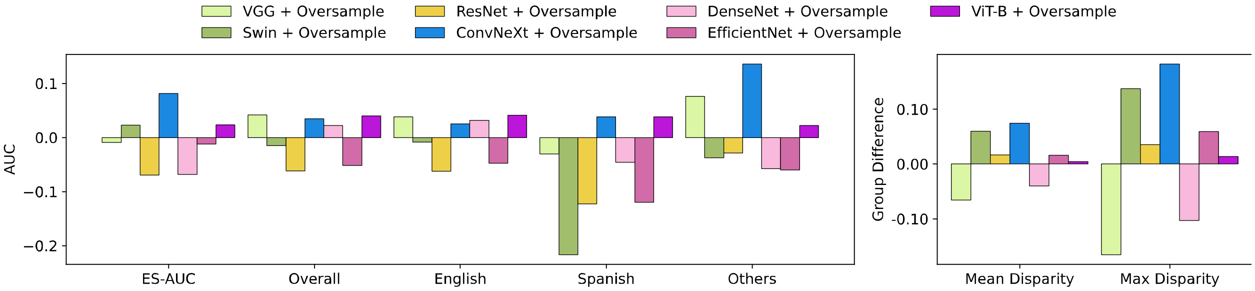


(b)

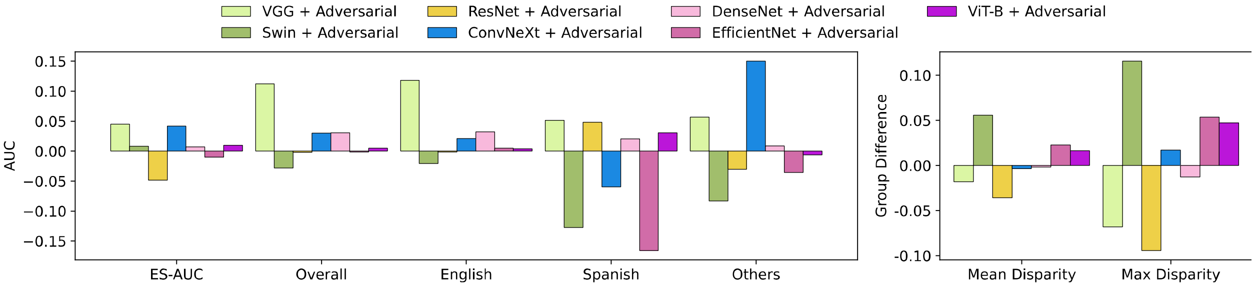


(c)

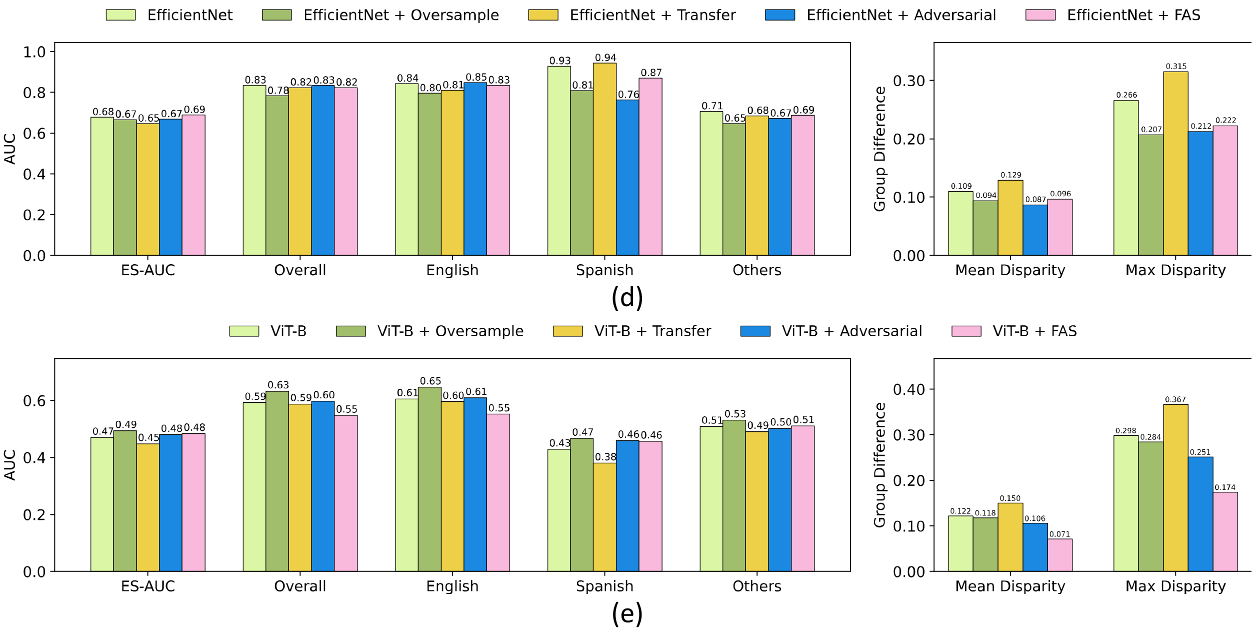


**Figure 23. Results on OCT B-scans on language attributes of the Harvard-FairVision30k dataset.** (a) The accuracy of various baseline models. (b) Differences in accuracy for various baseline models after applying oversampling techniques. (c) Differences in accuracy for various baseline models after applying adversarial techniques. (d) The accuracy of EfficientNet and its integration with oversampling, adversarial, transfer learning and our FAS techniques. (e) The accuracy of ViT-B and its integration with oversampling, adversarial, transfer learning and our FAS techniques.

##### Harvard-FairVision30k OCT B-scans: Marital Status

| | ES-AUC | Overall AUC | Married & Partnered | Single | Divorced | Widowed | Legally Separated | Mean Disparity | Max Disparity |
|---|---|---|---|---|---|---|---|---|---|
| VGG | 0.367 | 0.559 | 0.574 | 0.563 | 0.498 | 0.524 | 0.964 | 0.308 | 0.834 |
| Swin | 0.385 | 0.535 | 0.540 | 0.526 | 0.582 | 0.493 | 0.250 | 0.220 | 0.620 |
| ResNet | 0.601 | 0.769 | 0.765 | 0.790 | 0.755 | 0.758 | 1.000 | 0.122 | 0.318 |
| ConvNeXt | 0.408 | 0.542 | 0.561 | 0.494 | 0.500 | 0.545 | 0.759 | 0.179 | 0.488 |
| DenseNet | 0.593 | 0.713 | 0.697 | 0.730 | 0.749 | 0.670 | 0.804 | 0.064 | 0.188 |
| EfficientNet | 0.644 | 0.834 | 0.805 | 0.877 | 0.891 | 0.833 | 1.000 | 0.080 | 0.234 |
| ViT-B | 0.494 | 0.592 | 0.603 | 0.549 | 0.665 | 0.572 | 0.643 | 0.073 | 0.196 |

**Table 61. The accuracy of various baseline models on marital status attributes for Harvard-FairVision30k OCT B-scans.**

| | ES-AUC | Overall AUC | Married & Partnered | Single | Divorced | Widowed | Legally Separated | Mean Disparity | Max Disparity |
|---|---|---|---|---|---|---|---|---|---|
| VGG* | 0.064 | 0.041 | 0.018 | 0.049 | 0.164 | -0.009 | -0.143 | 0.137 | 0.323 |
| Swin* | 0.081 | -0.015 | -0.013 | 0.004 | -0.115 | 0.039 | 0.304 | 0.164 | 0.452 |

| | | | | | | | | | |
|---|---|---|---|---|---|---|---|---|---|
| ResNet* | -0.061 | -0.062 | -0.067 | -0.044 | -0.067 | -0.145 | -0.143 | 0.008 | -0.027 |
| ConvNeXt* | 0.085 | 0.035 | 0.021 | 0.039 | 0.150 | 0.007 | -0.205 | 0.108 | 0.285 |
| DenseNet* | 0.037 | 0.022 | 0.021 | 0.008 | 0.054 | 0.069 | -0.143 | 0.002 | -0.007 |
| EfficientNet* | -0.033 | -0.052 | -0.023 | -0.071 | -0.105 | -0.087 | 0.000 | -0.035 | -0.092 |
| ViT-B* | 0.013 | 0.040 | 0.050 | 0.070 | -0.026 | -0.049 | -0.107 | -0.013 | -0.010 |

**Table 62. Differences in accuracy of various baseline models after applying oversampling techniques on marital status attributes using Harvard-FairVision30k OCT B-scans.** * denotes a model with oversampling techniques.

| | ES-AUC | Overall AUC | Married & Partnered | Single | Divorced | Widowed | Legally Separated | Mean Disparity | Max Disparity |
|---|---|---|---|---|---|---|---|---|---|
| VGG# | 0.133 | -0.059 | -0.074 | -0.063 | 0.002 | -0.024 | -0.464 | 0.308 | 0.834 |
| Swin# | 0.052 | 0.035 | 0.058 | 0.022 | -0.091 | 0.063 | 0.161 | 0.106 | 0.290 |
| ResNet# | -0.026 | -0.018 | -0.036 | -0.004 | 0.040 | -0.019 | -0.054 | 0.018 | 0.029 |
| ConvNeXt# | 0.014 | 0.041 | 0.016 | 0.097 | 0.098 | 0.030 | 0.170 | -0.057 | -0.117 |
| DenseNet# | -0.014 | 0.026 | 0.035 | 0.040 | -0.026 | 0.054 | 0.143 | -0.051 | -0.114 |
| EfficientNet# | -0.003 | -0.026 | 0.005 | -0.069 | -0.029 | -0.073 | -0.036 | -0.006 | -0.020 |
| ViT-B# | -0.011 | -0.004 | 0.009 | -0.009 | -0.026 | -0.007 | -0.125 | -0.004 | -0.010 |

**Table 63. Differences in accuracy of various baseline models after applying adversarial techniques on marital status attributes using Harvard-FairVision30k OCT B-scans.** # denotes a model with adversarial techniques.

| | ES-AUC | Overall AUC | Married & Partnered | Single | Divorced | Widowed | Legally Separated | Mean Disparity | Max Disparity |
|---|---|---|---|---|---|---|---|---|---|
| EfficientNet | 0.644 | 0.834 | 0.805 | 0.877 | 0.891 | 0.833 | 1.000 | 0.080 | 0.234 |
| EfficientNet* | 0.611 | 0.782 | 0.782 | 0.806 | 0.786 | 0.746 | 1.000 | 0.115 | 0.325 |
| EfficientNet¶ | 0.587 | 0.828 | 0.796 | 0.878 | 0.894 | 0.737 | 1.000 | 0.108 | 0.318 |
| EfficientNet# | 0.641 | 0.808 | 0.810 | 0.808 | 0.862 | 0.760 | 0.964 | 0.086 | 0.253 |
| EfficientNet† | 0.620 | 0.816 | 0.804 | 0.856 | 0.807 | 0.744 | 1.000 | 0.106 | 0.313 |

**Table 64. The performances of EfficientNet and its variants on marital status attributes using Harvard-FairVision30k OCT B-scans.** * denotes EfficientNet with oversampling, ¶ denotes EfficientNet with transfer learning, # denotes EfficientNet with adversarial, and † denotes EfficientNet with our FAS techniques.

| | ES-AUC | Overall AUC | Married & Partnered | Single | Divorced | Widowed | Legally Separated | Mean Disparity | Max Disparity |
|---|---|---|---|---|---|---|---|---|---|
| ViT-B | 0.494 | 0.592 | 0.603 | 0.549 | 0.665 | 0.572 | 0.643 | 0.073 | 0.196 |
| ViT-B* | 0.507 | 0.632 | 0.653 | 0.619 | 0.638 | 0.523 | 0.536 | 0.085 | 0.206 |
| ViT-B¶ | 0.436 | 0.581 | 0.613 | 0.540 | 0.683 | 0.540 | 0.696 | 0.115 | 0.269 |
| ViT-B# | 0.483 | 0.589 | 0.612 | 0.539 | 0.639 | 0.565 | 0.518 | 0.076 | 0.206 |
| ViT-B† | 0.480 | 0.583 | 0.598 | 0.596 | 0.583 | 0.446 | 0.536 | 0.099 | 0.260 |

**Table 65. The performances of ViT-B and its variants on marital status attributes using Harvard-FairVision30k OCT B-scans.** * denotes ViT-B with oversampling, ¶ denotes EfficientNet

with transfer learning, # denotes EfficientNet with adversarial, and † denotes ViT-B with our FAS techniques.

**Harvard-FairVision30k OCT B-scans: Gender + Race**

| | ES-AUC | Overall AUC | Female Asian | Female Black | Female White | Male Asian | Male Black | Male White | Mean Disparity | Max Disparity |
|---|---|---|---|---|---|---|---|---|---|---|
| VGG | 0.380 | 0.559 | 0.341 | 0.353 | 0.565 | 0.55 | 0.564 | 0.584 | 0.185 | 0.434 |
| Swin | 0.478 | 0.535 | 0.552 | 0.484 | 0.507 | 0.536 | 0.553 | 0.532 | 0.046 | 0.128 |
| ResNet | 0.569 | 0.769 | 0.574 | 0.719 | 0.750 | 0.715 | 0.764 | 0.796 | 0.092 | 0.289 |
| ConvNeXt | 0.442 | 0.542 | 0.579 | 0.634 | 0.488 | 0.53 | 0.531 | 0.563 | 0.084 | 0.269 |
| DenseNet | 0.577 | 0.713 | 0.667 | 0.698 | 0.689 | 0.599 | 0.736 | 0.727 | 0.064 | 0.193 |
| EfficientNet | 0.606 | 0.834 | 0.683 | 0.855 | 0.795 | 0.704 | 0.850 | 0.854 | 0.086 | 0.206 |
| ViT-B | 0.408 | 0.592 | 0.417 | 0.458 | 0.664 | 0.540 | 0.585 | 0.603 | 0.143 | 0.417 |

**Table 66. The accuracy of various baseline models on gender and race attributes for Harvard-FairVision30k OCT B-scans.**

| | ES-AUC | Overall AUC | Female Asian | Female Black | Female White | Male Asian | Male Black | Male White | Mean Disparity | Max Disparity |
|---|---|---|---|---|---|---|---|---|---|---|
| VGG* | 0.162 | 0.041 | 0.217 | 0.249 | 0.000 | 0.055 | 0.023 | 0.026 | 0.153 | 0.349 |
| Swin* | -0.049 | -0.015 | -0.086 | -0.056 | -0.010 | -0.016 | 0.000 | -0.022 | -0.030 | -0.110 |
| ResNet* | 0.007 | -0.062 | 0.094 | 0.055 | -0.070 | -0.083 | -0.044 | -0.081 | 0.029 | 0.089 |
| ConvNeXt* | 0.023 | 0.035 | 0.047 | -0.104 | 0.179 | 0.014 | 0.044 | -0.006 | 0.000 | 0.031 |
| DenseNet* | -0.01 | 0.022 | 0.017 | -0.074 | 0.078 | 0.071 | 0.034 | 0.007 | -0.009 | -0.006 |
| EfficientNet* | -0.064 | -0.052 | -0.209 | -0.096 | -0.048 | 0.036 | -0.058 | -0.049 | -0.058 | -0.218 |
| ViT-B* | 0.051 | 0.040 | 0.080 | 0.043 | 0.004 | 0.056 | 0.034 | 0.055 | 0.035 | 0.147 |

**Table 67. Differences in accuracy of various baseline models after applying oversampling techniques on gender and race attributes using Harvard-FairVision30k OCT B-scans.** * denotes a model with oversampling techniques.

| | ES-AUC | Overall AUC | Female Asian | Female Black | Female White | Male Asian | Male Black | Male White | Mean Disparity | Max Disparity |
|---|---|---|---|---|---|---|---|---|---|---|
| VGG# | 0.153 | 0.102 | 0.300 | 0.147 | 0.108 | 0.087 | 0.104 | 0.091 | 0.093 | 0.170 |
| Swin# | -0.086 | 0.010 | -0.233 | 0.177 | 0.051 | -0.002 | -0.023 | 0.021 | -0.142 | -0.500 |
| ResNet# | 0.118 | 0.034 | 0.200 | 0.085 | 0.063 | -0.023 | 0.043 | 0.023 | 0.037 | 0.129 |
| ConvNeXt# | -0.010 | 0.028 | -0.133 | -0.040 | -0.022 | 0.058 | 0.023 | 0.041 | -0.026 | -0.008 |
| DenseNet# | -0.081 | 0.074 | -0.180 | -0.063 | 0.122 | 0.121 | 0.046 | 0.098 | -0.087 | -0.236 |
| EfficientNet# | 0.045 | -0.015 | 0.067 | -0.033 | 0.002 | -0.017 | -0.004 | -0.029 | 0.020 | 0.011 |
| ViT-B# | 0.025 | -0.025 | -0.011 | -0.013 | -0.096 | 0.008 | -0.019 | -0.029 | 0.026 | 0.122 |

**Table 68. Differences in accuracy of various baseline models after applying adversarial techniques on gender and race attributes using Harvard-FairVision30k OCT B-scans.** # denotes a model with adversarial techniques.

| | ES-AUC | Overall AUC | Female Asian | Female Black | Female White | Male Asian | Male Black | Male White | Mean Disparity | Max Disparity |
|---|---|---|---|---|---|---|---|---|---|---|
| EfficientNet | 0.606 | 0.834 | 0.683 | 0.855 | 0.795 | 0.704 | 0.850 | 0.854 | 0.086 | 0.206 |
| EfficientNet * | 0.542 | 0.782 | 0.474 | 0.758 | 0.747 | 0.740 | 0.793 | 0.806 | 0.144 | 0.424 |
| EfficientNet# | 0.651 | 0.820 | 0.750 | 0.821 | 0.797 | 0.687 | 0.847 | 0.825 | 0.066 | 0.195 |
| EfficientNet† | 0.605 | 0.831 | 0.622 | 0.776 | 0.814 | 0.765 | 0.834 | 0.855 | 0.092 | 0.281 |

**Table 69. The performances of EfficientNet and its variants on gender and race attributes using Harvard-FairVision30k OCT B-scans.** * denotes EfficientNet with oversampling, # denotes EfficientNet with adversarial, and † denotes EfficientNet with our FAS techniques.

| | ES-AUC | Overall AUC | Female Asian | Female Black | Female White | Male Asian | Male Black | Male White | Mean Disparity | Max Disparity |
|---|---|---|---|---|---|---|---|---|---|---|
| ViT-B | 0.408 | 0.592 | 0.417 | 0.458 | 0.664 | 0.540 | 0.585 | 0.603 | 0.143 | 0.417 |
| ViT-B* | 0.459 | 0.632 | 0.498 | 0.501 | 0.668 | 0.596 | 0.620 | 0.659 | 0.108 | 0.270 |
| ViT-B# | 0.432 | 0.567 | 0.406 | 0.444 | 0.568 | 0.548 | 0.566 | 0.574 | 0.118 | 0.295 |
| ViT-B† | 0.419 | 0.555 | 0.383 | 0.441 | 0.559 | 0.528 | 0.555 | 0.564 | 0.124 | 0.327 |

**Table 70. The performances of ViT-B and its variants on gender and race attributes using Harvard-FairVision30k OCT B-scans.** * denotes ViT-B with oversampling, # denotes EfficientNet with adversarial, and † denotes ViT-B with our FAS techniques.

**Harvard-FairVision30k OCT B-scans: Gender + Ethnicity**

| | ES-AUC | Overall AUC | Female Non-Hispanic | Female Hispanic | Female Non-Hispanic | Male Hispanic | Mean Disparity | Max Disparity |
|---|---|---|---|---|---|---|---|---|
| VGG | 0.411 | 0.559 | 0.562 | 0.573 | 0.511 | 0.264 | 0.225 | 0.553 |
| Swin | 0.512 | 0.535 | 0.543 | 0.526 | 0.542 | 0.556 | 0.020 | 0.056 |
| ResNet | 0.700 | 0.769 | 0.752 | 0.776 | 0.810 | 0.805 | 0.030 | 0.074 |
| ConvNeXt | 0.511 | 0.542 | 0.521 | 0.565 | 0.548 | 0.530 | 0.031 | 0.081 |
| DenseNet | 0.666 | 0.713 | 0.727 | 0.700 | 0.723 | 0.680 | 0.027 | 0.067 |
| EfficientNet | 0.712 | 0.834 | 0.831 | 0.824 | 0.923 | 0.905 | 0.052 | 0.118 |
| ViT-B | 0.501 | 0.592 | 0.594 | 0.583 | 0.664 | 0.494 | 0.102 | 0.287 |

**Table 71. The accuracy of various baseline models on gender and ethnicity attributes for Harvard-FairVision30k OCT B-scans.**

| | ES-AUC | Overall AUC | Female Non-Hispanic | Female Hispanic | Female Non-Hispanic | Male Hispanic | Mean Disparity | Max Disparity |
|---|---|---|---|---|---|---|---|---|
| VGG* | 0.091 | 0.041 | 0.025 | 0.044 | 0.016 | 0.242 | 0.151 | 0.369 |
| Swin* | -0.095 | -0.015 | -0.007 | -0.015 | 0.011 | -0.225 | -0.151 | -0.372 |
| ResNet* | -0.066 | -0.062 | -0.045 | -0.066 | -0.088 | -0.199 | -0.035 | -0.088 |
| ConvNeXt* | -0.016 | 0.035 | 0.071 | -0.014 | 0.119 | 0.013 | -0.054 | -0.134 |
| DenseNet* | -0.126 | 0.022 | 0.030 | 0.015 | 0.133 | -0.143 | -0.130 | -0.367 |
| EfficientNet* | -0.004 | -0.052 | -0.058 | -0.041 | -0.152 | -0.039 | 0.002 | -0.003 |
| ViT-B* | -0.014 | 0.040 | 0.031 | 0.063 | -0.029 | -0.134 | -0.087 | -0.166 |

**Table 72. Differences in accuracy of various baseline models after applying oversampling techniques on gender and ethnicity attributes using Harvard-FairVision30k OCT B-scans.** * denotes a model with oversampling techniques.

| | ES-AUC | Overall AUC | Female Non-Hispanic | Female Hispanic | Female Non-Hispanic | Male Hispanic | Mean Disparity | Max Disparity |
|---|---|---|---|---|---|---|---|---|
| VGG# | 0.089 | -0.058 | -0.062 | -0.073 | -0.011 | 0.236 | 0.224 | 0.553 |
| Swin# | -0.081 | 0.007 | -0.010 | 0.012 | 0.048 | 0.182 | -0.133 | -0.322 |
| ResNet# | -0.042 | -0.004 | -0.005 | -0.004 | 0.044 | -0.091 | -0.037 | -0.108 |
| ConvNeXt# | -0.067 | -0.004 | 0.053 | -0.049 | -0.013 | -0.145 | -0.101 | -0.269 |
| DenseNet# | -0.018 | 0.050 | 0.030 | 0.073 | 0.058 | -0.061 | -0.060 | -0.145 |
| EfficientNet# | 0.021 | -0.015 | -0.006 | -0.016 | -0.139 | -0.017 | 0.006 | -0.007 |
| ViT-B# | 0.000 | -0.032 | -0.031 | -0.024 | -0.171 | 0.022 | 0.049 | 0.162 |

**Table 73. Differences in accuracy of various baseline models after applying adversarial techniques on gender and ethnicity attributes using Harvard-FairVision30k OCT B-scans** # denotes a model with adversarial techniques.

| | ES-AUC | Overall AUC | Female Non-Hispanic | Female Hispanic | Female Non-Hispanic | Male Hispanic | Mean Disparity | Max Disparity |
|---|---|---|---|---|---|---|---|---|
| EfficientNet | 0.712 | 0.834 | 0.831 | 0.824 | 0.923 | 0.905 | 0.052 | 0.118 |
| EfficientNet* | 0.708 | 0.782 | 0.772 | 0.783 | 0.771 | 0.866 | 0.050 | 0.121 |
| EfficientNet# | 0.732 | 0.819 | 0.824 | 0.809 | 0.785 | 0.887 | 0.046 | 0.126 |
| EfficientNet† | 0.779 | 0.829 | 0.842 | 0.810 | 0.804 | 0.836 | 0.019 | 0.046 |

**Table 74. The performances of EfficientNet and its variants on gender and ethnicity attributes using Harvard-FairVision30k OCT B-scans** * denotes EfficientNet with oversampling, # denotes EfficientNet with adversarial, and † denotes EfficientNet with our FAS techniques.

| | ES-AUC | Overall AUC | Female Non-Hispanic | Female Hispanic | Female Non-Hispanic | Male Hispanic | Mean Disparity | Max Disparity |
|---|---|---|---|---|---|---|---|---|
| ViT-B | 0.501 | 0.592 | 0.594 | 0.583 | 0.664 | 0.494 | 0.102 | 0.287 |
| ViT-B* | 0.488 | 0.632 | 0.625 | 0.646 | 0.635 | 0.359 | 0.189 | 0.453 |
| ViT-B# | 0.501 | 0.561 | 0.563 | 0.558 | 0.492 | 0.515 | 0.053 | 0.125 |
| ViT-B† | 0.497 | 0.560 | 0.563 | 0.557 | 0.479 | 0.520 | 0.060 | 0.150 |

**Table 75. The performances of ViT-B and its variants on gender and ethnicity attributes using Harvard-FairVision30k OCT B-scans** * denotes ViT-B with oversampling, # denotes EfficientNet with adversarial, and † denotes ViT-B with our FAS techniques.

##### Impact of the Ratio of Training Data

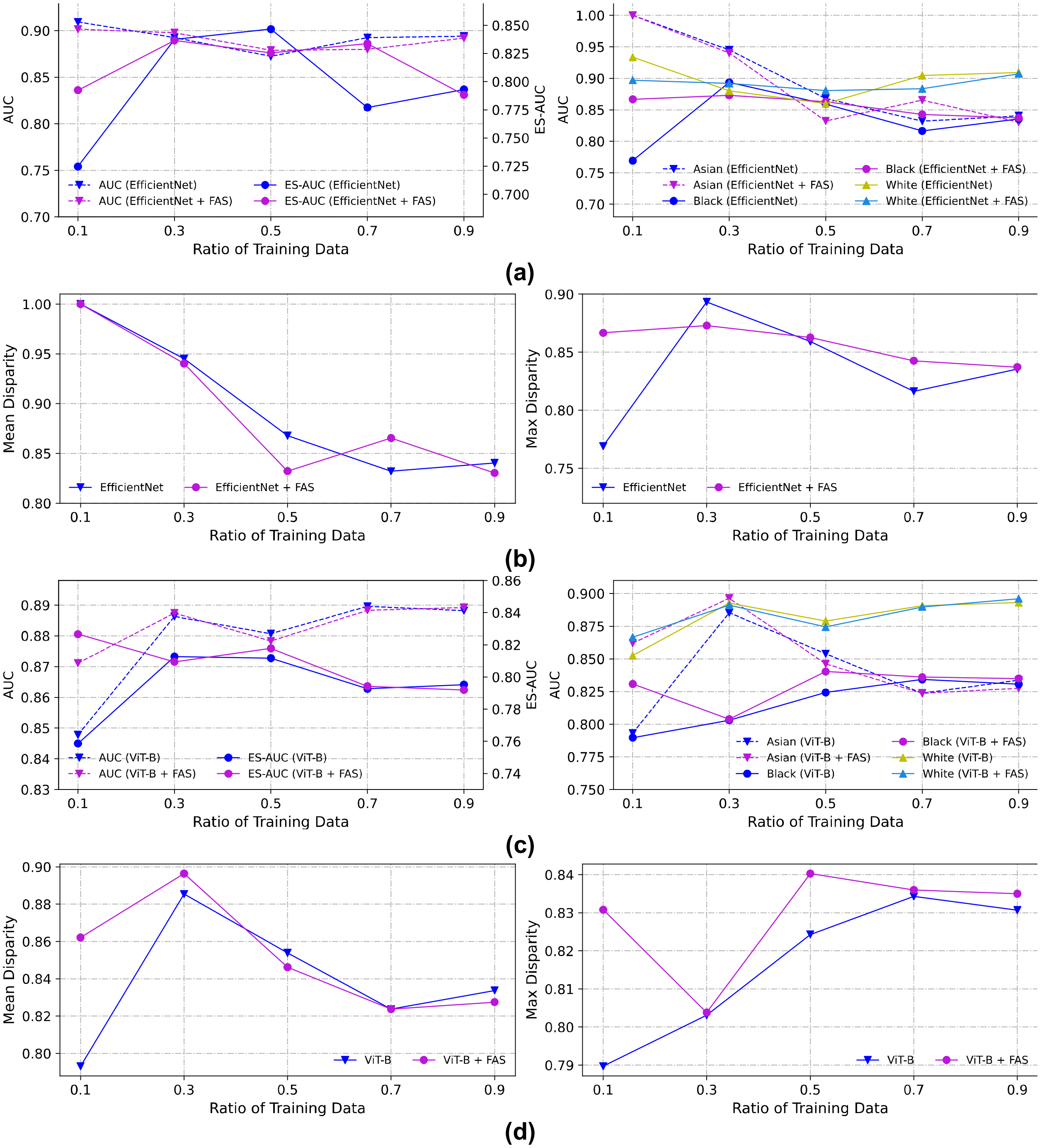


**Figure 24. The impact of different ratios of training data on race using in-house color fundus images.** (a) Overall AUC, ES- AUC and group AUCs for the EfficientNet for Asians, Blacks and Whites. (b) Mean and max disparities of the EfficientNet with and without adding FAS. (c) Overall AUC, ES- AUC and group AUCs for the ViT-B for Asians, Blacks and Whites. (d) Mean and max disparities of the ViT-B with and without adding FAS.

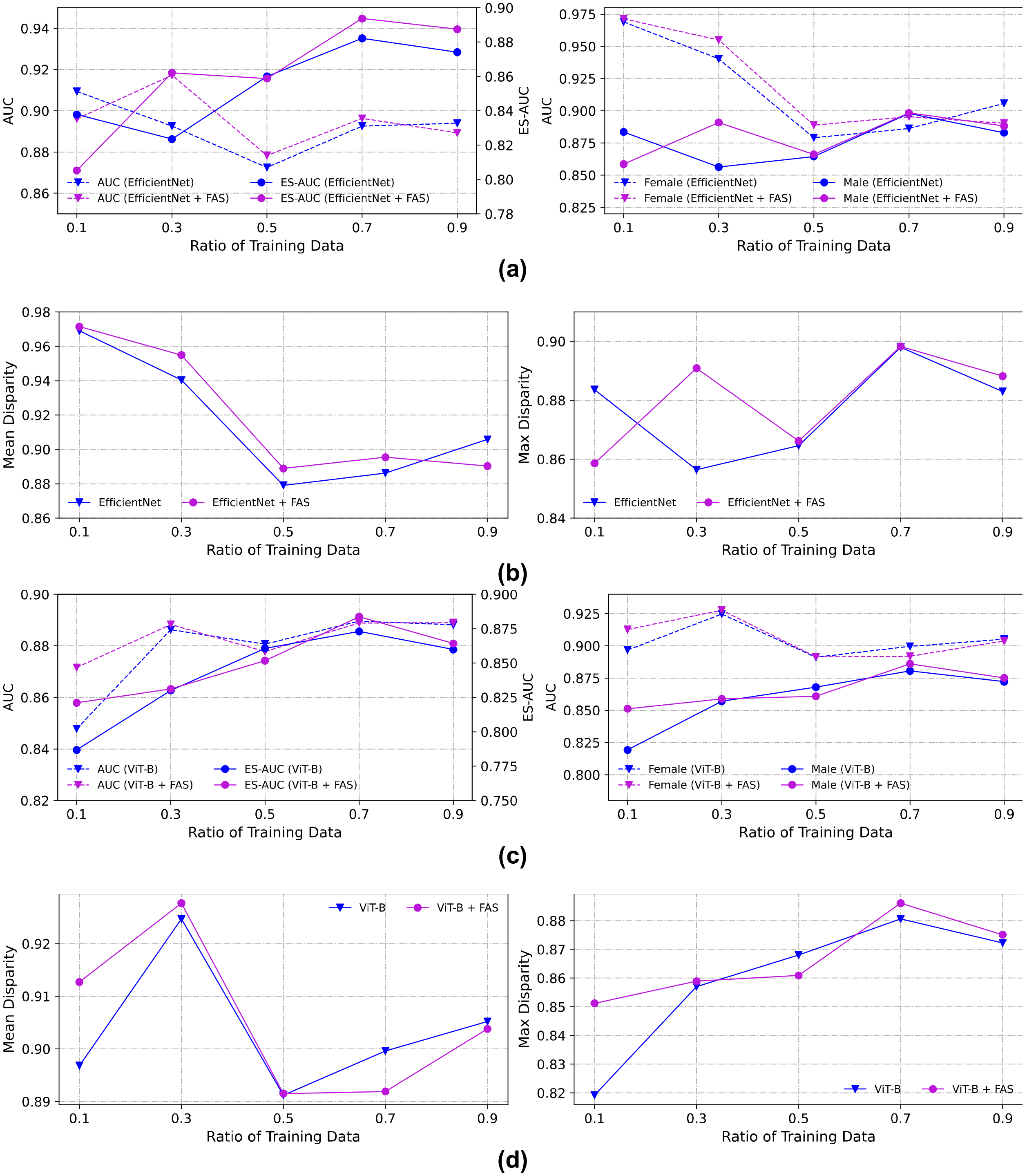


**Figure 25. The impact of different ratios of training data on gender using in-house color fundus images.** (a) Overall AUC, ES- AUC and group AUCs for the EfficientNet for Females and Males. (b) Mean and max disparities of the EfficientNet with and without adding FAS. (c) Overall AUC, ES- AUC and group AUCs for the ViT-B for Females and Males. (d) Mean and max disparities of the ViT-B with and without adding FAS.

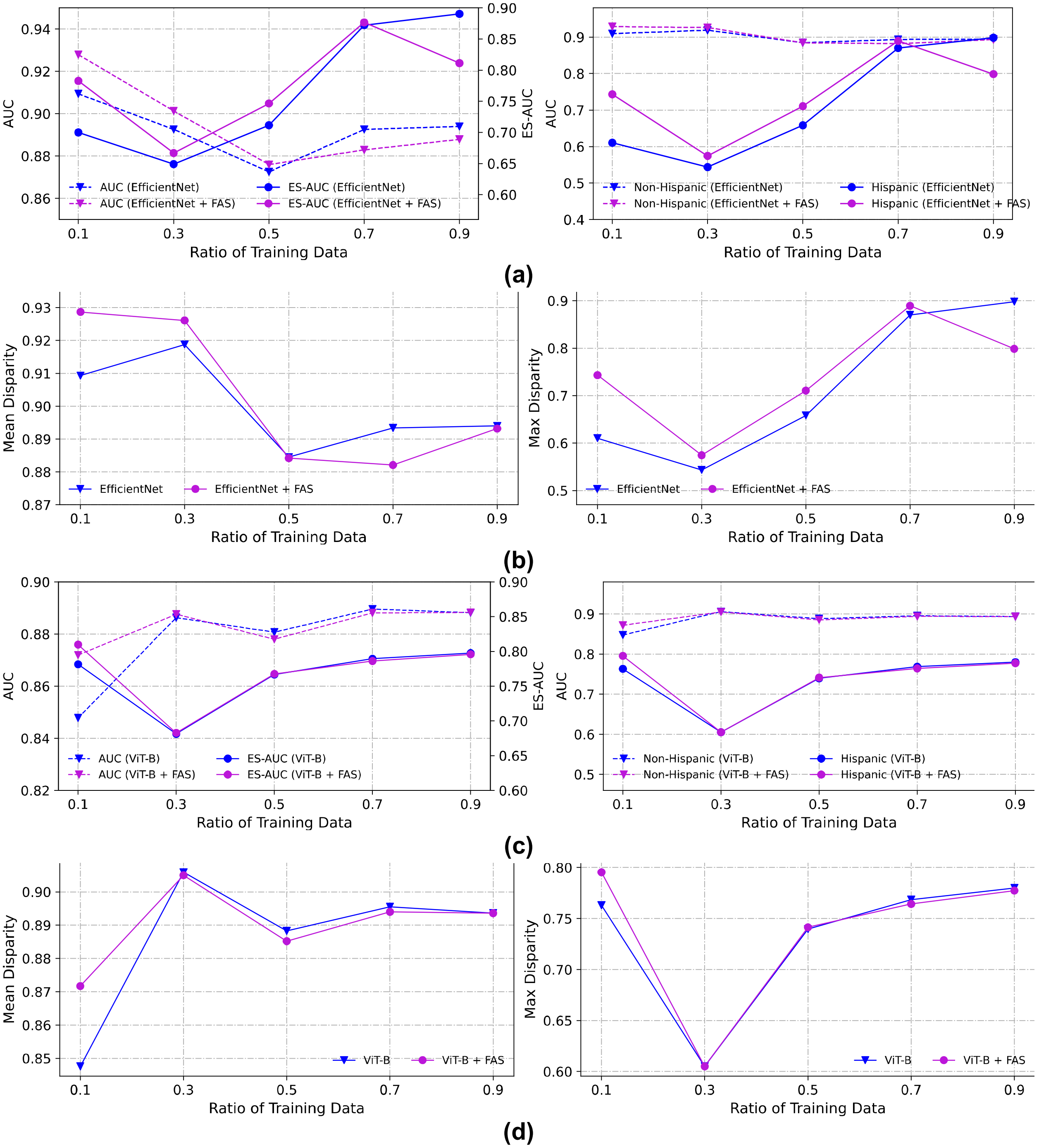


**Figure 26. The impact of different ratios of training data on ethnicity using in-house color fundus images.** (a) Overall AUC, ES- AUC and group AUCs for the EfficientNet for Non-Hispanics and Hispanics. (b) Mean and max disparities of the EfficientNet with and without adding FAS. (c) Overall AUC, ES- AUC and group AUCs for the ViT-B for Non-Hispanics and Hispanics. (d) Mean and max disparities of the ViT-B with and without adding FAS.

##### Inhouse Color Fundus Sensitivity Results

| | Overall Sensitivity | Asian | Black | White |
|---|---|---|---|---|
| EfficientNet | 0.723 | 0.563 | 0.629 | 0.737 |
| ViT-B | 0.752 | 0.667 | 0.731 | 0.764 |
| EfficientNet + FAS | 0.741 | 0.556 | 0.609 | 0.795 |
| ViT-B + FAS | 0.776 | 0.769 | 0.627 | 0.781 |

**Table 76. The sensitivity of EfficientNet and ViT-B at specificity = 0.9 on race using in-house color fundus images**.

| | Overall Sensitivity | Asian | Black | White |
|---|---|---|---|---|
| EfficientNet | 0.628 | 0.563 | 0.543 | 0.660 |
| ViT-B | 0.693 | 0.556 | 0.596 | 0.696 |
| EfficientNet + FAS | 0.655 | 0.556 | 0.543 | 0.705 |
| ViT-B + FAS | 0.706 | 0.769 | 0.490 | 0.723 |

**Table 77. The sensitivity of EfficientNet and ViT-B at specificity = 0.95 on race using in-house color fundus images**.

| | Overall Sensitivity | Female | Male |
|---|---|---|---|
| EfficientNet | 0.723 | 0.744 | 0.709 |
| ViT-B | 0.752 | 0.780 | 0.734 |
| EfficientNet + FAS | 0.741 | 0.752 | 0.721 |
| ViT-B + FAS | 0.804 | 0.837 | 0.774 |

**Table 78. The sensitivity of EfficientNet and ViT-B at specificity = 0.9 on gender using in-house color fundus images**.

| | Overall Specificity | Female | Male |
|---|---|---|---|
| EfficientNet | 0.628 | 0.656 | 0.598 |
| ViT-B | 0.693 | 0.688 | 0.697 |
| EfficientNet + FAS | 0.618 | 0.606 | 0.622 |
| ViT-B + FAS | 0.701 | 0.745 | 0.679 |

**Table 79. The sensitivity of EfficientNet and ViT-B at specificity = 0.95 on gender using in-house color fundus images**.

| | Overall Sensitivity | Non-Hispanic | Hispanic |
|---|---|---|---|
| EfficientNet | 0.723 | 0.726 | 0.625 |
| ViT-B | 0.752 | 0.767 | 0.563 |
| EfficientNet + FAS | 0.752 | 0.760 | 0.600 |
| ViT-B + FAS | 0.740 | 0.749 | 0.615 |

**Table 80. The sensitivity of EfficientNet and ViT-B at specificity = 0.9 on ethnicity using in-house color fundus images**.

| | Overall Sensitivity | Non-Hispanic | Hispanic |
|---|---|---|---|
| EfficientNet | 0.628 | 0.637 | 0.500 |
| ViT-B | 0.693 | 0.708 | 0.438 |
| EfficientNet + FAS | 0.614 | 0.615 | 0.600 |
| ViT-B + FAS | 0.663 | 0.672 | 0.538 |

**Table 81. The sensitivity of EfficientNet and ViT-B at specificity = 0.95 on ethnicity using in-house color fundus images**.
